# Tracking funding disparities in global health aid with machine learning

**DOI:** 10.1101/2025.06.20.25329993

**Authors:** Finn Stürenburg, Kerstin Forster, Nicolas Banholzer, Malte Toetzke, Kenneth Harttgen, Stefan Feuerriegel

**Author notes:** These authors contributed equally to this work (Finn Stürenburg, Kerstin Forster).

## Abstract

Reducing the global burden of disease is crucial for improving health outcomes worldwide. However, misalignment between health aid and country-level disease burden leaves vulnerable populations without necessary support for major health challenges, particularly in the least developed countries. In this paper, we develop a machine learning pipeline using large language models to track flows in official development assistance (ODA) earmarked for health and identify aid–burden misalignment. Specifically, we classified 3.7 million development aid projects from 2000 to 2022 (USD ∼332 billion) into 17 major categories of communicable, maternal, neonatal, and nutritional diseases (CMNNDs) and non-communicable diseases (NCDs). We then compared the rank of per capita ODA disbursement against the rank of disease burden (i.e., disability-adjusted life years [DALYs]) to identify relative aid–burden misalignment at the country level. Even though funding and disease burden are significantly correlated for many diseases, there are notable disparities. For example, NCDs account for 59.5% of global DALYs but received only 2.5% of health-related ODA. This disparity is particularly concerning because, in low and middle-income countries, there is an increasing double burden: not only from the traditional burden of CMNNDs, but also from the rising burden of NCDs. Our results also show that several regions face severe aid–burden misalignment across multiple diseases including Central Africa and parts of South Asia and West Africa. Our results identify health disparities to inform public policy decisions in development aid assistance. Overall, our machine learning approach supports the targeted allocation of health aid where it is most needed, thereby reducing the global burden of diseases.

## Introduction

Progress in global health has slowed since 2015 [1–5]. As of now, only one out of the five health-related Sustainable Development Goals is on track for 2030 [6, 7], with low- and middle-income countries facing large setbacks. These countries are confronted by a double burden of both severely under-resourced health systems [8, 9] and high disease burden from both communicable, maternal, neonatal, and nutritional diseases (CMNNDs) and non-communicable diseases (NCDs). For example, new HIV infections are three times above the 2025 goal [10], progress on reducing tuberculosis (TB) deaths is less than a third of the target [11], and cases for malaria have increased instead of reaching the reduction target for 2025 [12]. Without faster action, none of the six WHO regions will meet the 2030 mortality targets for NCDs [1]. Recent cuts to health aid – particularly by major donors such as the United States – are further widening the gap between health needs and available resources [13–16].

Official development assistance (ODA; hereafter referred to as aid) provides a key source of funding to address global health challenges, particularly in the least developed countries but also lower-middle-income and upper-middle-income countries that lack sufficient domestic resources [17–21]. However, substantial variation exists in how ODA is distributed across diseases and regions, leading to persistent gaps between funding needs and available resources [22–30]. Yet, systematic evidence on whether aid aligns with disease burden is limited. The Institute for Health Metrics and Evaluation’s Financing Global Health series provides comprehensive trends in development assistance for health from 1990–present across broad health areas, including non-communicable and neglected tropical diseases, and incorporates private foundation and NGO flows [31], but does not provide granular country-level assessments of ODA allocations relative to disease burden. Other existing databases are typically constrained to only a few common diseases (e.g., typically only HIV/AIDS, TB, malaria and maternal and neonatal health [32–39]) and often lack comprehensive temporal and geographic coverage (e.g., the focus is often limited exclusively to Africa or a few selected countries such as the Countdown countries) [32–35]. Most analyses precede 2020 and thus do not reflect the impact of the COVID-19 pandemic. Moreover, the aforementioned works aimed at tracking and analyzing ODA flows rely heavily on manual coding, which is not scalable and limits timely and comprehensive evidence for policy decision-making.

Here, we develop a machine learning pipeline using large language models to track ODA flows for public health (see Figure 1). This enables us to analyze the global allocation of ODA ear-marked to CMNNDs and NCDs and assess whether the distribution of funding is in line with the country-specific disease burden. Using our machine learning pipeline, we classified 3.7 million ODA projects from 2000 to 2022 (∼USD 332 billion across 157 recipient countries) into 17 major categories of CMNNDs and NCDs. We focus on donor contributions (ODA disbursements) and not domestic government or household health spending. We then assessed the alignment between disease-specific ODA flows and disease burden across major causes as defined by the Global Burden of Disease study [40, 41]. Specifically, we compared the rank of per capita ODA disbursement against the rank of disease burden (i.e., disability-adjusted life years [DALYs]) to identify relative aid–burden misalignment at the country level. We interpret aid–burden misalignment as a descriptive measure that highlights divergence, not as a prescriptive criterion for optimal funding allocation, given legitimate alternative objectives and acknowledging that DALY-based metrics reflect specific value assumptions [42, 43]. Our machine learning pipeline is scalable and allows for regular updating. It supports evidence-based policies in global health by identifying health needs and informs the targeted allocation of health aid to reduce the global burden of diseases.

**Fig. 1.**
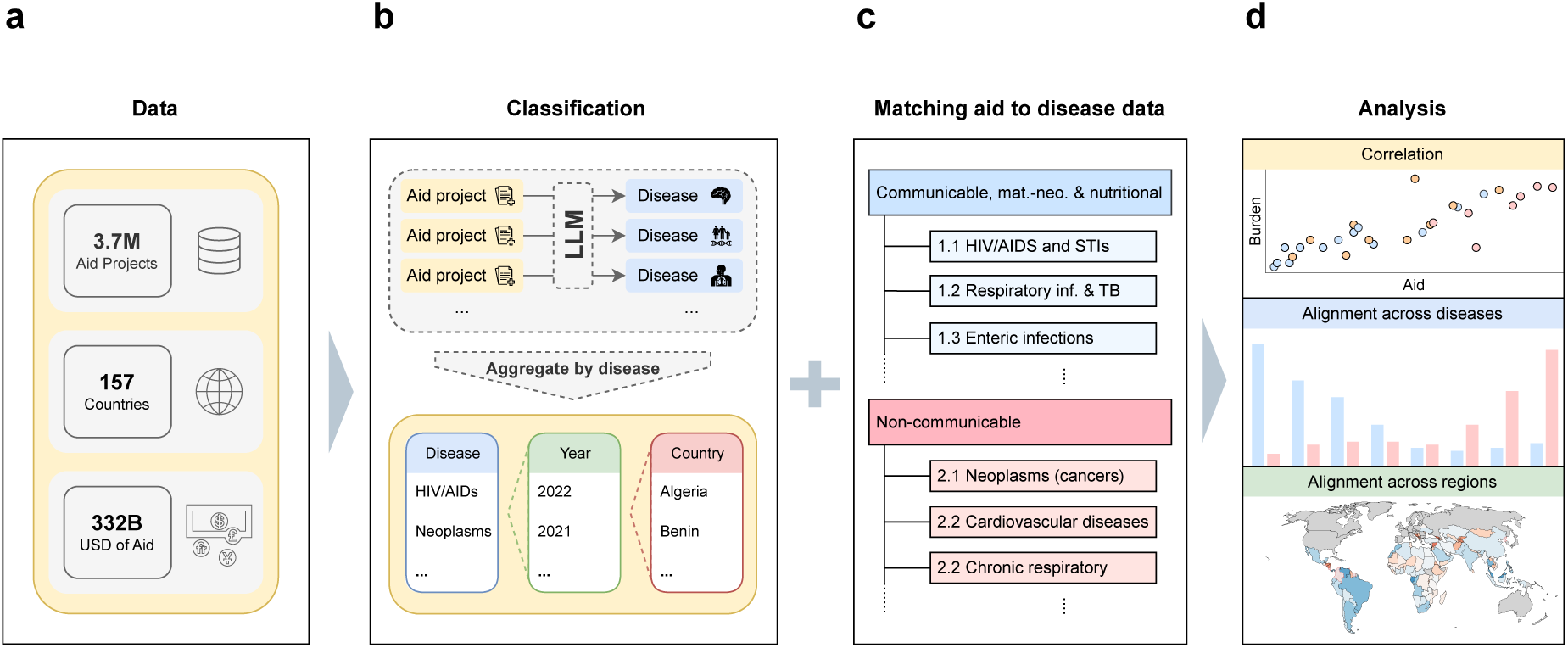
Machine learning pipeline to identify funding needs in global health. Our machine learning pipeline tracks country-level disparities between aid flows and disease burden across major diseases and regions to identify funding needs. **a**, The input to our machine learning pipeline are the project descriptions of 3.7 million development aid projects from around the globe. **b**, We then used a state-of-the-art, open-source large language model (Llama-3.1-70B) to perform multi-label classification of aid projects into major diseases. Here, we focus on 17 major causes of disease burden, grouped into CMNNDs and NCDs, as defined by the Global Burden of Disease study [40, 41]. Further, we aggregated the funding by disease, year, and country. **c**, We then matched the disease-specific ODA to health outcomes at the country level, namely, the disease burden as measured by disability-adjusted life years (DALYs) from the Global Burden of Disease study [40, 41]. **d**, Our machine learning pipeline then allows to analyze the alignment (e.g., via Spearman’s rank correlation coefficient) between ODA flows and disease burden. Together, our machine learning pipeline offers a scalable way to identify funding needs, which allows for regular updates to inform evidence-based policies in global health.

## Results

We analyzed 3.71 million development aid projects from 2000–2022 as provided by the Creditor Reporting System [44] of the Organization of Economic Cooperation and Development (OECD), which is the standardized reporting system for donors in the development sector across the world. Using an open-source large language model (Meta-Llama-3.1-70B-Instruct), we classified all projects into 17 causes of disease burden as defined by the Global Burden of Disease study [40,41]. Our categorization distinguishes (i) CMNNDs (e.g., HIV/AIDS and sexually transmitted infections [STIs], respiratory infections and TB, nutritional deficiencies) and (ii) NCDs (e.g., mental disorders, neoplasms, substance use disorders). The full list of disease categories is provided in Supplementary Table S1. We validated the classification from our machine learning approach by comparing it against (i) a keyword-based classification approach and (ii) manual annotations of 2,060 projects (see Supplementary Material S1), which confirms the accuracy of our machine learning pipeline. Of the 3.71 million projects, our machine learning pipeline classified 321,527 as being related to one of the major diseases (8.7%), while the remaining projects relate to other sustainability targets (e.g., climate action). We then aggregated the ODA flows across disease, year, and country. To contextualize these flows, we summarized the distribution of funding by donor (Supplementary Table S15) and found that global health ODA is dominated by a small number of countries and international funds as donors, and their funding decisions may have the largest impact on reducing the aid–burden disparities identified in this analysis.

### Distribution of global aid earmarked to health

The distribution of ODA shows large disparities between CMNNDs and NCDs (Figure 2). From 2000 to 2022, a total of USD 332 billion was allocated to disease-specific aid projects, of which 97.5% (USD 324 billion) targeted CMNNDs. In contrast, only 2.5% (USD 8 billion) was directed toward NCDs. Most funding was concentrated in a few areas, with HIV/AIDS and STIs alone receiving USD 112.4 billion and respiratory infections and TB receiving USD 79.4 billion. By comparison, NCDs received considerably less (e.g., cardiovascular diseases: USD 0.6 billion; diabetes including kidney disorders: USD 0.6 billion; and mental disorders: USD 2.5 billion), despite growing prevalence in low- and middle-income countries [8, 9, 40].

**Fig. 2.**
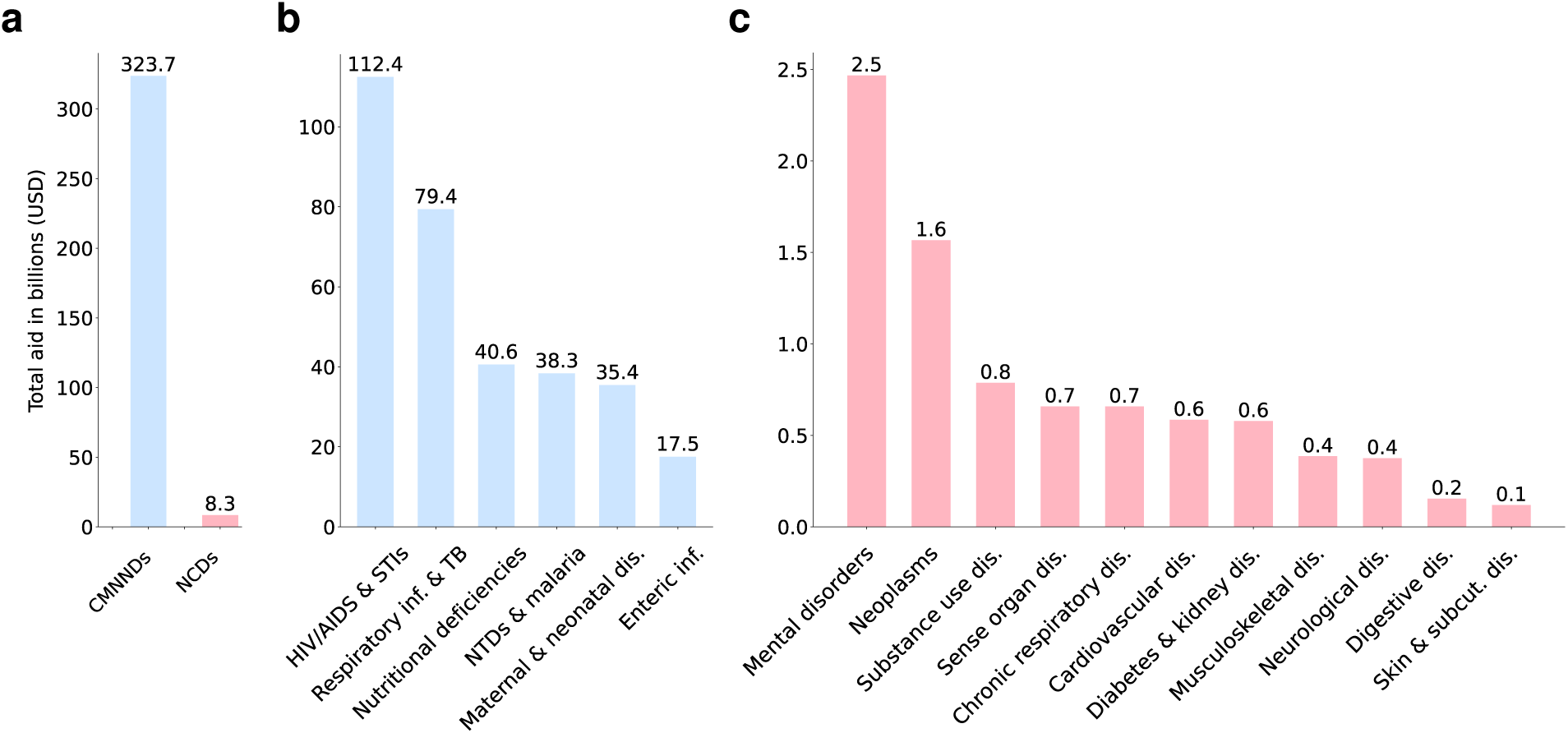
Distribution of total development aid (2000–2022) across diseases. Shown is **a**, the aggregate comparison of total aid between CMNNDs and NCDs, alongside total aid for **b**, CMNNDs and **c**, NCDs. The majority of funding was allocated to CMNNDs, particularly HIV/AIDS and STIs, and respiratory infections and TB. The categorization of diseases follows the Global Burden of Disease study [40, 41]. Abbreviations: STIs, sexually transmitted infections; TB, tuberculosis; NTDs, neglected tropical diseases.

Between 2000 and 2022, global health funding increased from USD 783 million per year to USD 35 billion per year (Figure 3; overall aid in this time period: USD 332 billion), which corresponds to a more than 40-fold increase. However, funding priorities shifted between 2000 and 2022. For example, between 2000 and 2022, funding for HIV/AIDS and STIs rose from USD 174 million to USD 6.9 billion, though their share of total aid declined over time; funding for nutritional deficiencies rose from USD 93 million to USD 4 billion; and funding for neglected tropical diseases and malaria rose from USD 77 million to USD 4 billion. Further, a large increase is seen in ODA earmarked for respiratory infections and TB during the COVID-19 pandemic, which reached an annual volume of USD 44 billion in 2020, before declining to USD 35 billion by 2022.

**Fig. 3.**
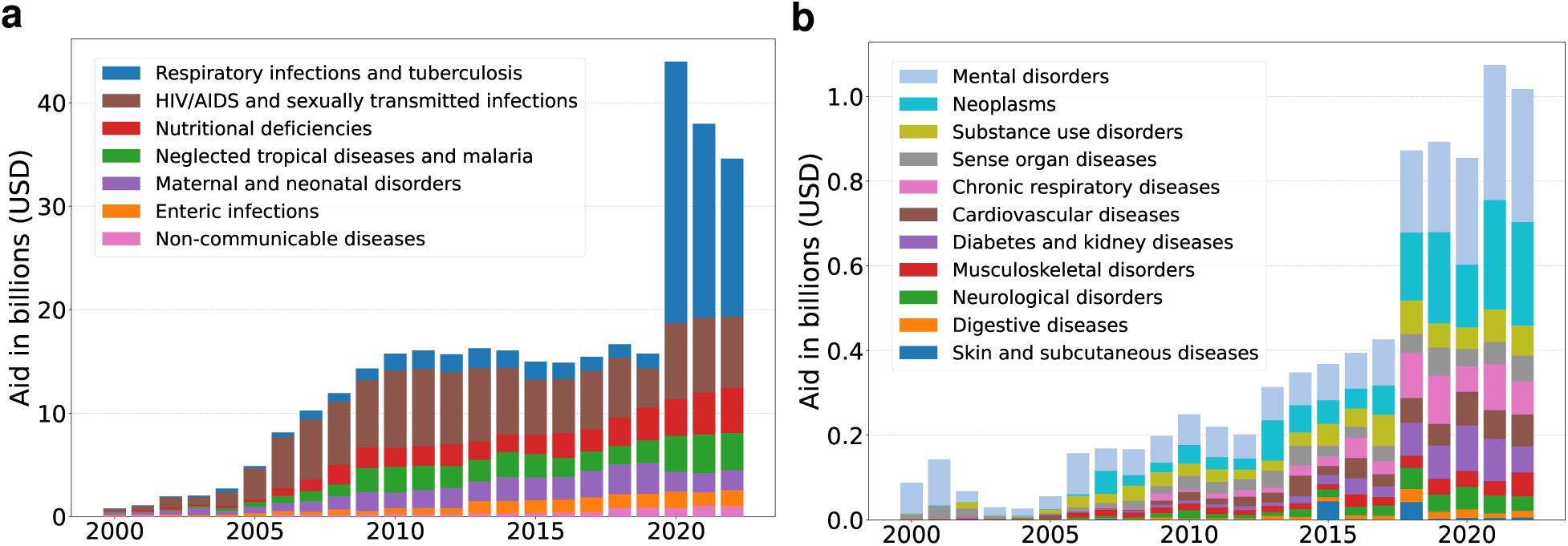
Temporal trends in annual global health aid across diseases (2000–2022). Shown are annual aid disbursements by disease category for **a**, CMNNDs and **b**, NCDs. Overall, annual aid increased from USD 0.78 billion in 2000 to USD 34.60 billion in 2022, with notable peaks in funding for respiratory infections and TB during the COVID-19 pandemic.

Over 2000–2022, accumulated disease-specific ODA per capita varies substantially across countries for both CMNNDs (Figure 4) and NCDs (Figure 5). For CMNNDs, ODA for HIV/AIDS and STIs is highest in Southern Africa (e.g., Eswatini: USD 623, Namibia: USD 557, Botswana: USD 495); ODA for respiratory infections and TB is highest in Central Asia and the Caucasus (e.g., Mongolia: USD 200, Kazakhstan: USD 105, Georgia: USD 80); ODA for nutritional deficiencies is highest in the Horn of Africa, the Middle East, and the Sahel (USD 50–110); ODA for neglected tropical diseases and malaria is highest in West Africa (USD 55–75). ODA for maternal and neonatal disorders is more broadly distributed, with notable per capita highs in South Sudan and Sierra Leone (USD 58–72). ODA for enteric infections is generally lower, typically below USD 30 per capita. For NCDs, ODA per capita is generally lower than for CMNNDs and more concentrated in a limited set of countries, with minimal disease-specific funding elsewhere.

**Fig. 4.**
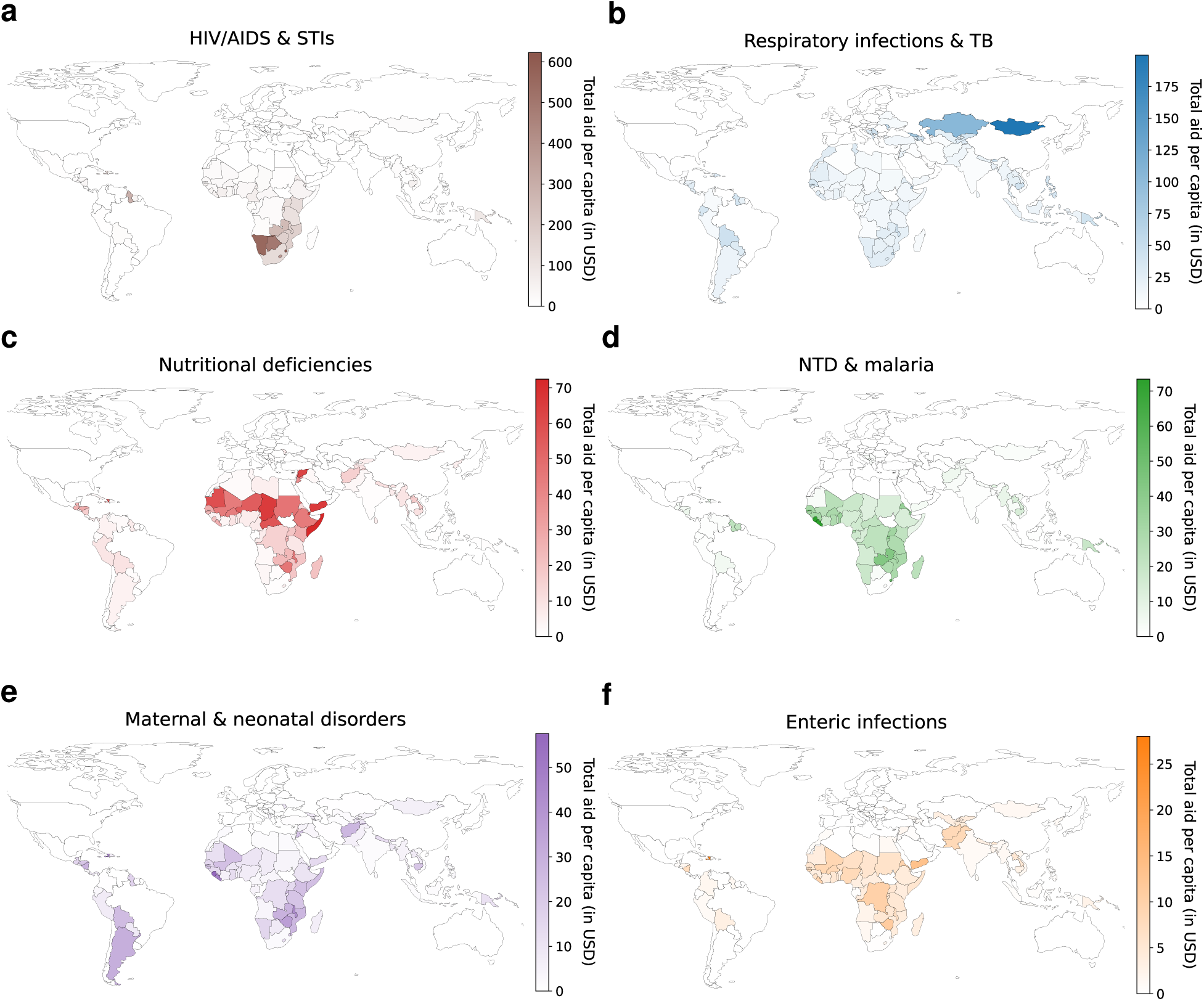
Geographic variation of global health aid (for CMNNDs). Maps show accumulated disease-specific ODA per capita (in USD) across countries from 2000 to 2022. The plots show **a**, HIV/AIDS and STIs; **b**, respiratory infections and TB; **c**, nutritional deficiencies; **d**, NTD and malaria; **e**, maternal and neonatal disorders; and **f**, enteric infections. Darker colors indicate higher per capita aid. Abbreviations: STIs, sexually transmitted infections; TB, tuberculosis; NTDs, neglected tropical diseases.

**Fig. 5.**
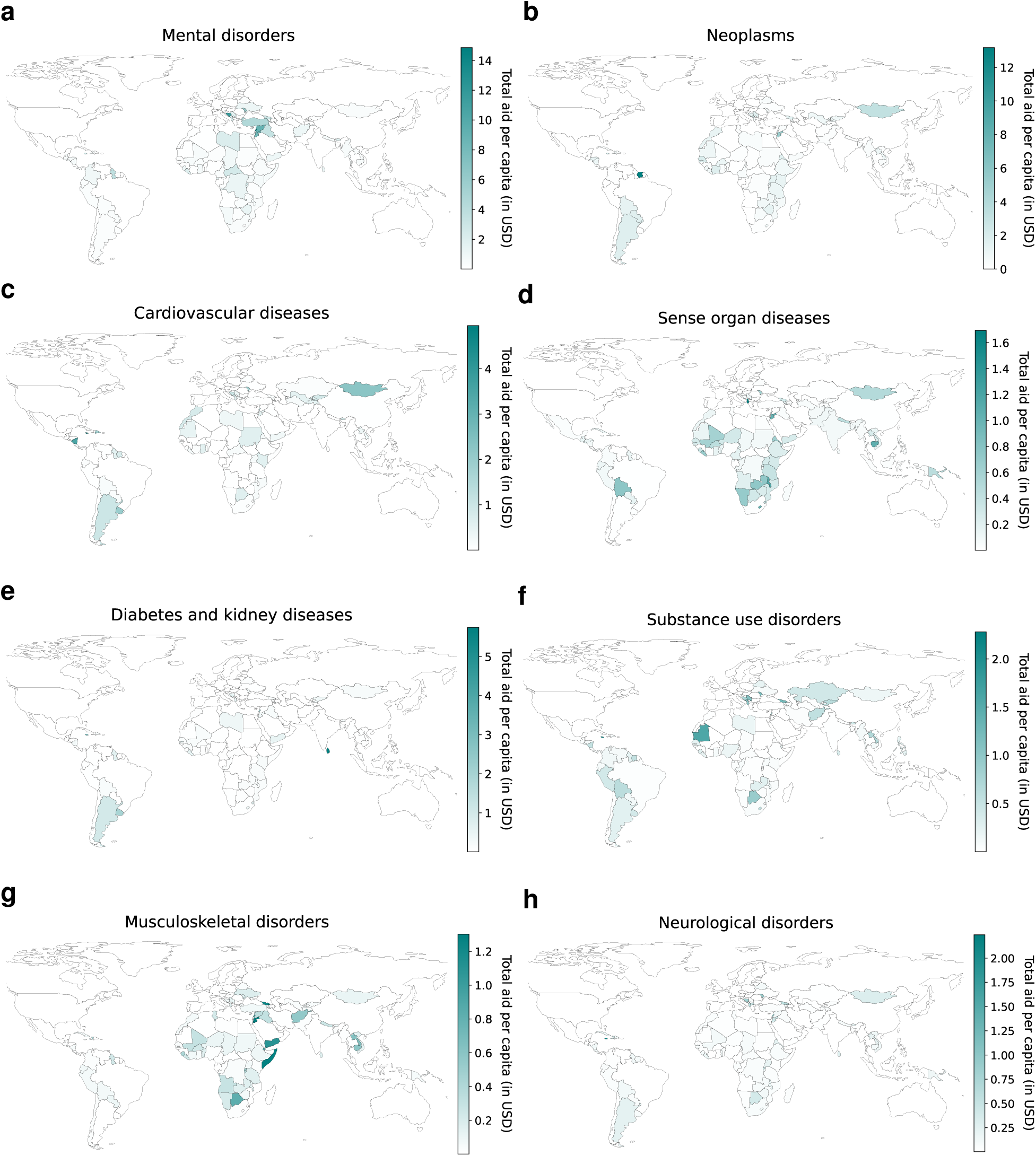
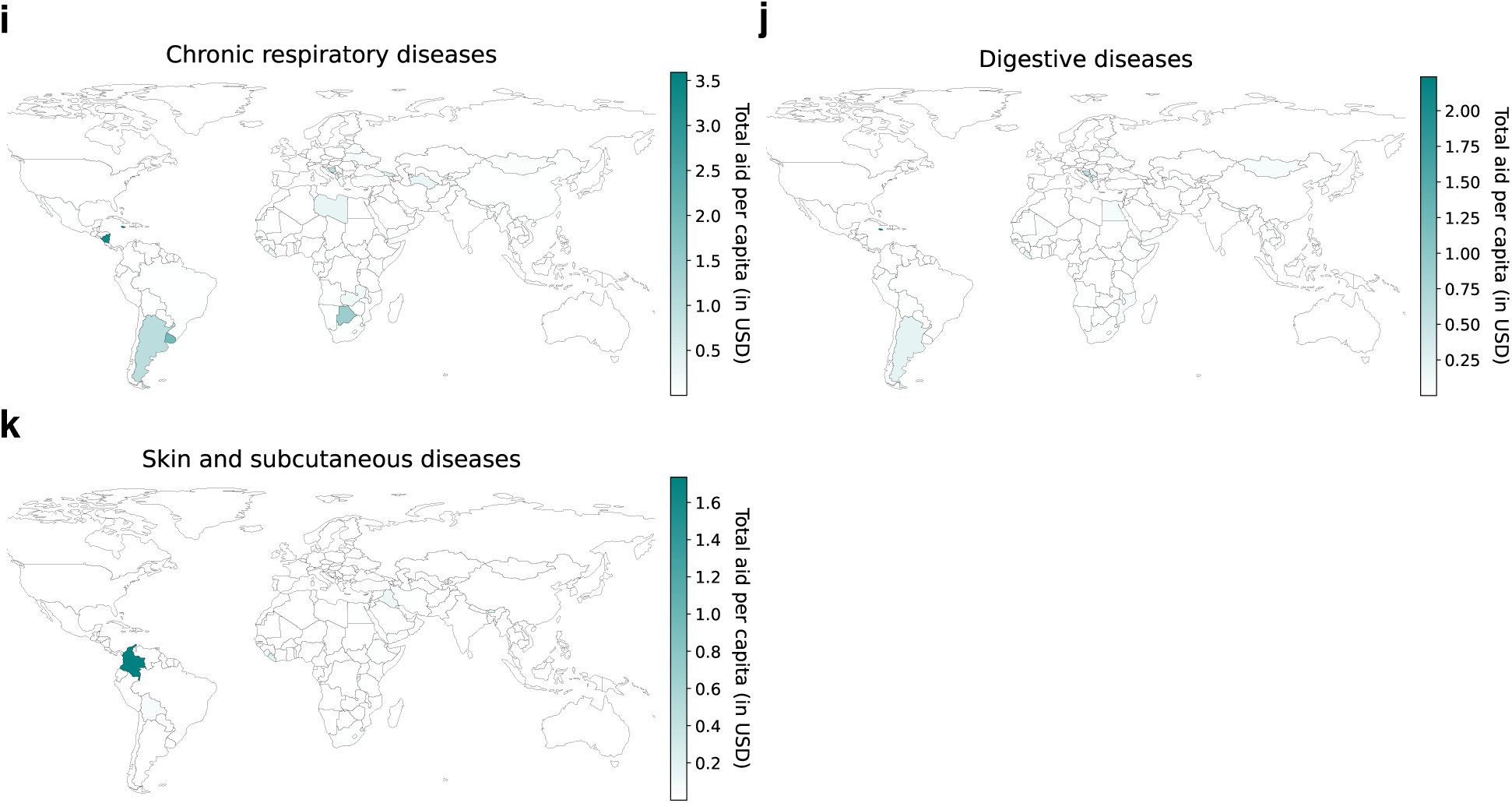
Geographic variation of global health aid (for NCDs). Maps show accumulated disease-specific ODA per capita (in USD) across countries from 2000 to 2022. Darker colors indicate higher per capita aid; white indicates countries with no recorded allocation of aid. The plots show **a**, mental disorders; **b**, neoplasms; **c**, cardiovascular diseases; **d**, sense organ diseases; **e**, diabetes and kidney diseases; **f**, substance use disorders; **g**, musculoskeletal disorders; **h**, neurological disorders; **i**, chronic respiratory diseases; **j**, digestive diseases; and **k**, skin and subcutaneous diseases. Darker colors indicate higher per capita aid.

### Alignment between aid and disease burden

Our analysis points to substantial disparities between the proportion of allocated ODA for each disease and the corresponding proportion of the global disease burden (Figure 6). While such disparities may be driven by unobserved factors (e.g., historical or donor-driven priorities, political economy), they point to aid–burden misalignment, whereby some diseases receive less aid relative to their share of global disease burden. We interpret aid–burden misalignment as a descriptive measure of divergence rather than as evidence of misallocation: first, departures from aid–burden alignment may reflect legitimate priorities (e.g., equity, contagion control, or system strengthening). Second, DALY aggregation—blending severity and prevalence—embeds value choices that shape interpretation [42]. Accordingly, we treat aid–burden alignment as a decision-relevant reference point rather than a prescriptive criterion for optimal funding allocation. We find that HIV/AIDS and STIs, neglected tropical diseases and malaria, and nutritional deficiencies received disproportionately high ODA volumes compared to the respective disease burden. For example, HIV/AIDS and STIs account for 6% of the global disease burden, but received 34% of total health aid, and similarly large disparities are present for neglected tropical diseases and malaria (5% of burden vs. 15% of aid) and nutritional deficiencies (3% of burden vs. 12% of aid). In contrast, NCDs account for 59.5% of the overall disease burden but received only a small fraction of global health aid (2.5%). Such disparities are especially pronounced, for instance, for cardiovascular diseases (17% of burden vs. 1% of aid), diabetes and kidney diseases (5.4% of burden vs. 0.2% of aid), neoplasms (10% of burden vs. *<*1% of aid), and mental disorders (6% of burden vs. *<*1% of aid), which received low funding despite the substantial negative impact on global health.

**Fig. 6.**
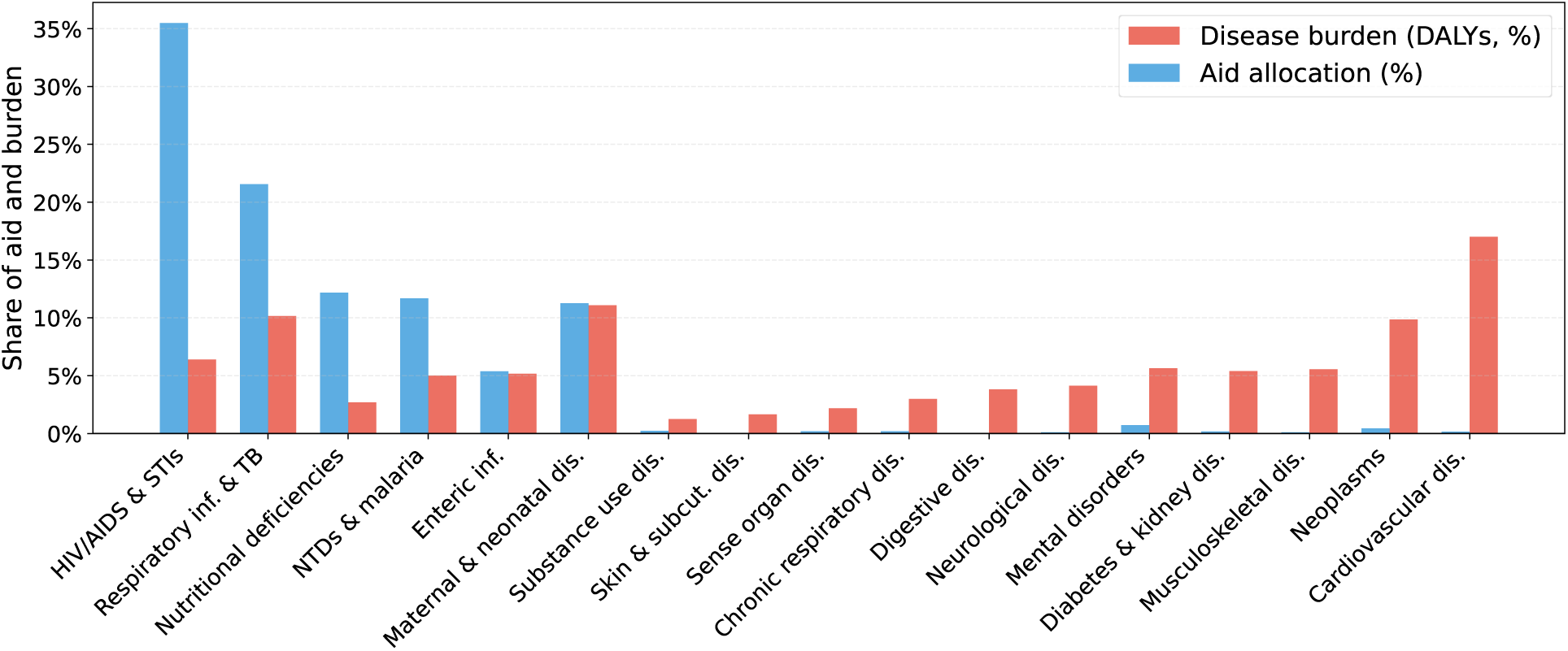
Disparities between aid volume and disease burden. Shown are (a) the proportion of total health aid allocated to each disease category (in blue) and (b) the proportion of the accumulated global disease burden (in red). Total health aid is based on the accumulative disbursement from 2000 to 2021. Disease burden is based on DALYs from 2000 to 2021 using the latest available data from the Global Burden of Disease study at the time of our analysis. Deviations between these proportions point to disparities; that is, diseases receiving less aid relative to their burden may reflect misalignment of funding relative to burden, while those receiving more may reflect historical or donor-driven priorities. Abbreviations: STIs, sexually transmitted infections; TB, tuberculosis; NTDs, neglected tropical diseases.

To analyze how well aid allocation between countries is aligned with country-level disease burden, we analyzed the correlation between per capita aid and disease burden using Spearman’s rank correlation coefficient (Figure 7). These correlations reveal how closely funding aligns with needs across diseases and country income groups (i.e., least developed countries [LDCs], lower-middle-income countries [LMICs], and upper-middle-income countries [UMICs], as per the country income group categorization of the OECD [44]). Overall, CMNNDs show stronger and more consistent positive correlations compared to NCDs (Figure 7, right column). For instance, HIV/AIDS and STIs (*ρ*= 0.71; *p <* 0.001) and NTDs and malaria (*ρ*= 0.78; *p <* 0.001) exhibit strong alignment. In contrast, several NCDs show negative correlations (i.e., skin and subcutaneous diseases, sense organ diseases, and musculoskeletal disorders), meaning that greater disease burden is associated with less aid. Results are robust to using disease incidence instead of DALYs: correlations between incidence and disease-specific per capita aid show largely consistent alignment patterns (Supplementary Figure S3).

**Fig. 7.**
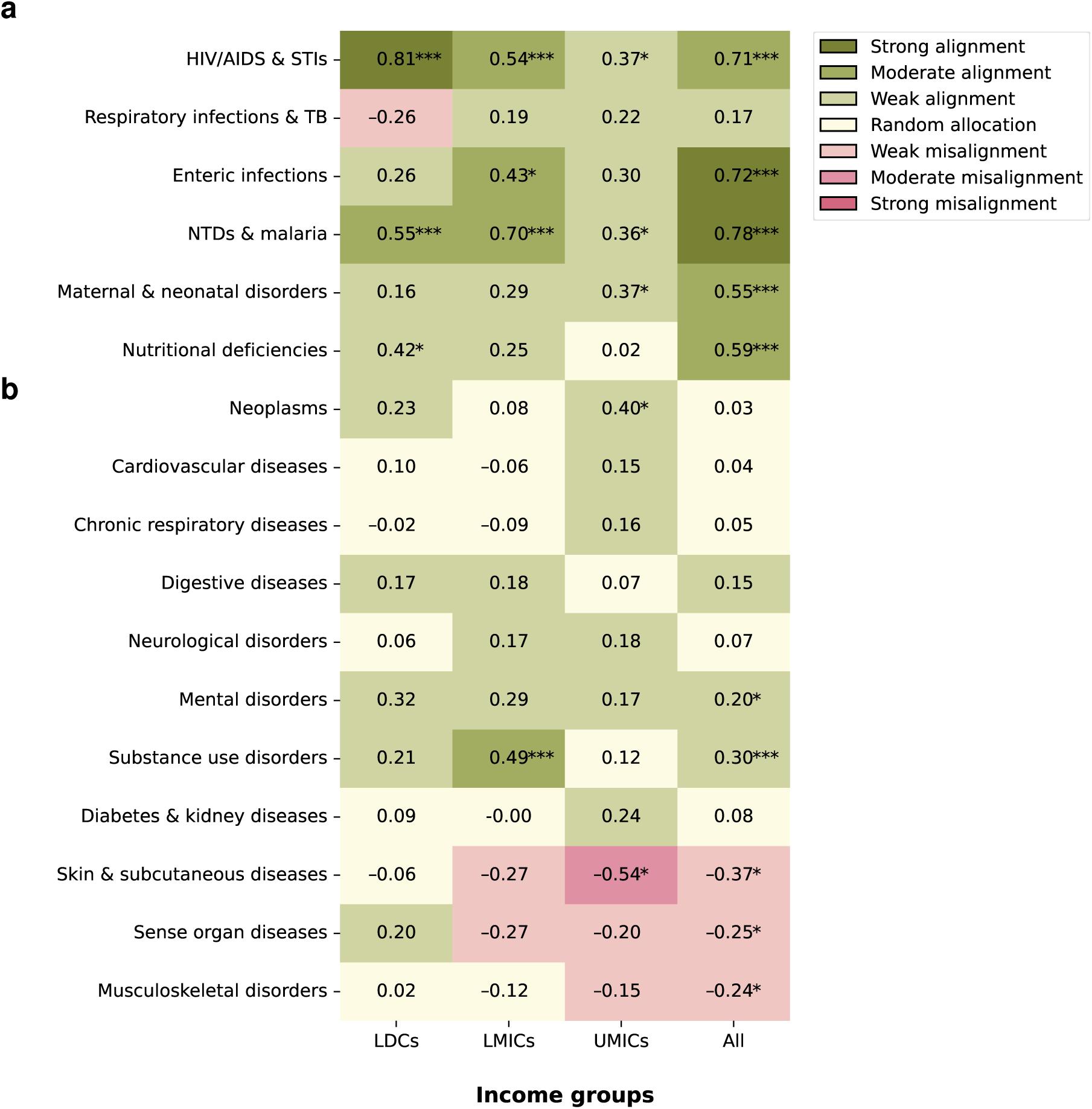
Correlation between disease-specific aid and burden across income groups. Shown are Spearman’s rank correlation coefficients between per capita aid (accumulated over the time period 2000–2021) and disease burden across country income groups: least developed countries (LDCs), lower-middle-income countries (LMICs), upper-middle-income countries (UMICs), and all countries combined. **a**, The top six rows report CMNNDs, which generally show positive correlations across most income groups. **b**, In contrast, NCDs (bottom eleven rows) exhibit more heterogeneity in the correlations. Green indicates positive and red negative correlations. Asterisks denote statistically significant correlations based on a two-sided Spearman rank test (^∗^ *p <* 0.05; ^∗∗^ *p <* 0.01; ^∗∗∗^ *p <* 0.001). Legend interpretation: −1 to −0.7: strong misalignment of health aid with disease burden; −0.7 to −0.3: moderate misalignment; −0.3 to −0.1: weak misalignment, −0.1 to 0.1: indicative of random allocation; 0.1 to 0.3: weak alignment, 0.3 to 0.7, moderate alignment; and 0.7 to 1.0 strong alignment. Abbreviations: STIs, sexually transmitted infections; TB, tuberculosis; NTDs, neglected tropical diseases.

We also observe substantial variation across income groups (Figure 7, columns 1–3). For example, HIV/AIDS and STIs show a strong positive correlation in all income groups, particularly in LDCs. In contrast, respiratory infections and TB show weaker or even negative correlations in lower-income settings. When restricting the analysis to the COVID-19 period (2020–2021), the overall alignment patterns remain similar (Supplementary Figures S4, S5, S6), although respiratory infections and TB show reduced alignment with disease burden. This reflects the temporary disruption in funding patterns during the pandemic. On the other hand, excluding the pandemic years for respiratory infections and TB markedly strengthens the correlation (*ρ* = 0.63; *p <* 0.001; Supplementary Figures S7, S8, S9), which suggest that the COVID-19 aid surge substantially affected alignment for these diseases. Together, these patterns highlight that aid allocation across countries seems well targeted for CMNNDs in lowest-income settings, but also point to persistent disparities in funding for many NCDs.

### Geographic distribution of aid–burden misalignment

To locate potential aid–burden misalignment at the country level, we plot the country-level patterns of disease burden against aid per capita (see Figure 8 for CMNNDs and Supplementary Figure S1 for NCDs). This highlights countries with a mismatch between ODA flows and disease burden, that is, where the disease burden is substantially higher (or lower) than per capita aid. For example, in the case of HIV/AIDS and STIs, the Central African Republic, classified as a least developed country in the OECD CRS database, ranks at the 90th percentile for disease burden, but is only at the 52nd percentile for per capita ODA volume of all LDCs. Funding for NCDs shows similar disparities (Supplementary Figure S1).

**Fig. 8.**
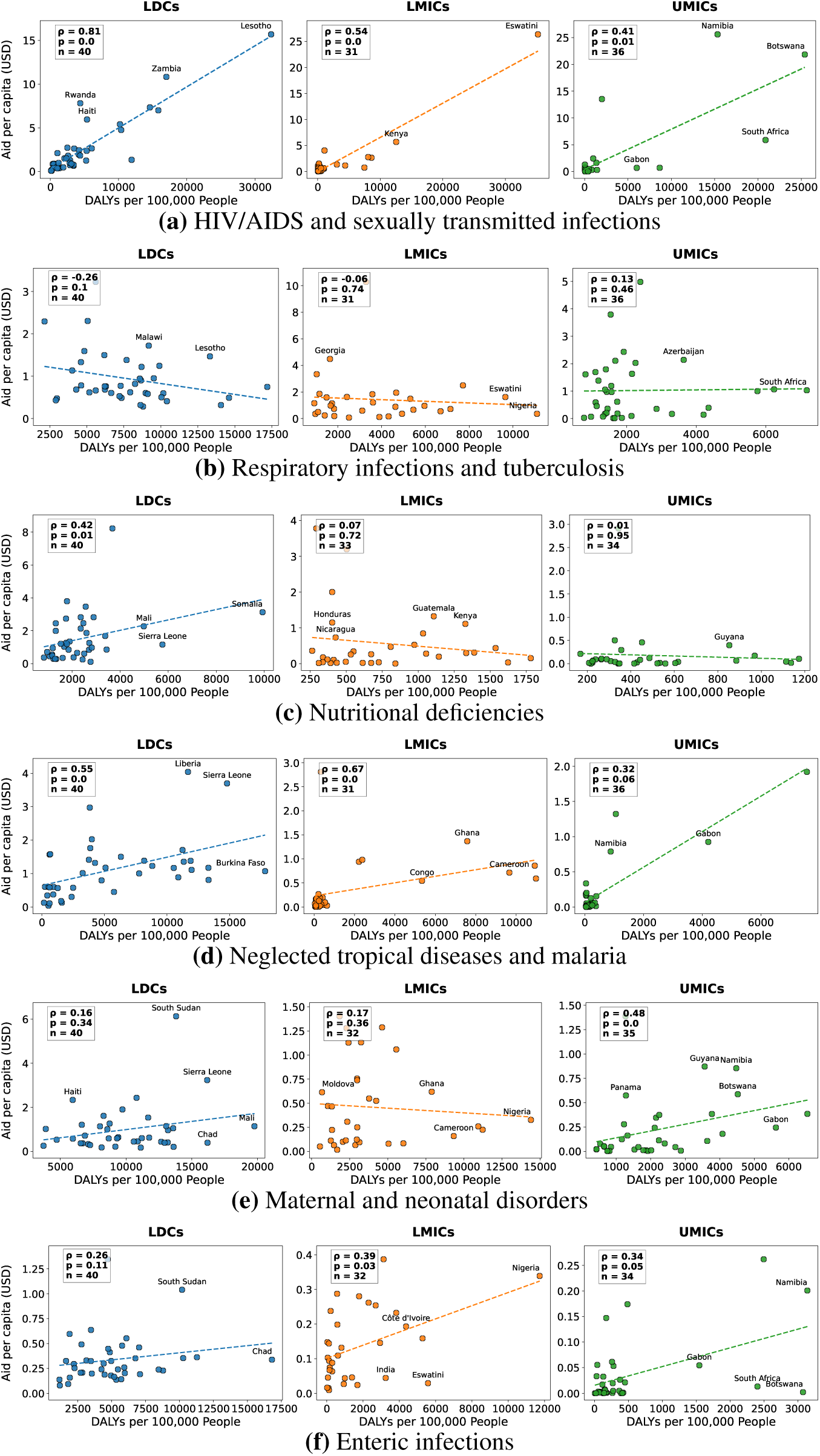
Funding disparities at the country level (for CMNNDs). The plots compare the disease burden (*x*-axis, measured by DALYs per 100,000 population, accumulated for 2000–2021) against the aid per capita (*y*-axis, accumulated for 2000–2021). Each point represents a single country, which allows to identify countries with high disease burden but comparatively low aid and thus to reveal aid–burden misalignment. For each disease, countries/plots are grouped by country income groups: least developed countries (LDCs) (*n* = 40 countries), lower-middle-income countries (LMICs) (*n* = 32) and upper-middle-income countries (UMICs) (*n* = 35). Due to space, we focus on selected diseases with high overall funding: **a**, HIV/AIDS and STIs; **b**, respiratory infections and TB; **c**, maternal and neonatal disorders; **d**, enteric infections; **e**, NTD and malaria; and **f**, nutritional deficiencies. Each plot reports Spearman’s correlation coefficient (*ρ*) and the *p*-value from a two-sided Spearman rank test. The dashed line is the fitted trend line from an ordinary least squares regression. Funding disparities for NCDs are presented in Supplementary Figure S1.

We find regions with aid allocated disproportionately to burden (see Figure 9 for CMNNDs and Supplementary Figure S2 for NCDs). Several regions show substantial aid–burden misalignment across multiple diseases, notably in Central Africa, West Africa, and parts of South Asia. Here, we refer to “aid–burden misalignment” as the difference between a country’s percentile rank in disease burden (DALYs per 100,000 population) and its percentile rank in per capita aid, calculated within income groups to account for potential heterogeneity in macroeconomic factors such as the price of labor and the general state of health systems. Aid–burden misalignment captures relative disparity within income groups, not necessarily absolute underfunding. For example, countries from Southern Africa exhibit aid–burden misalignment for enteric infections, nutritional deficiencies, respiratory infections and TB, maternal and neonatal disorders, but these countries appear relatively well funded for HIV/AIDS and STIs, which reflects historic donor priorities in these regions [45–48]. The geographic distribution of aid–burden misalignment for NCDs is shown in Supplementary Figure S2.

**Fig. 9.**
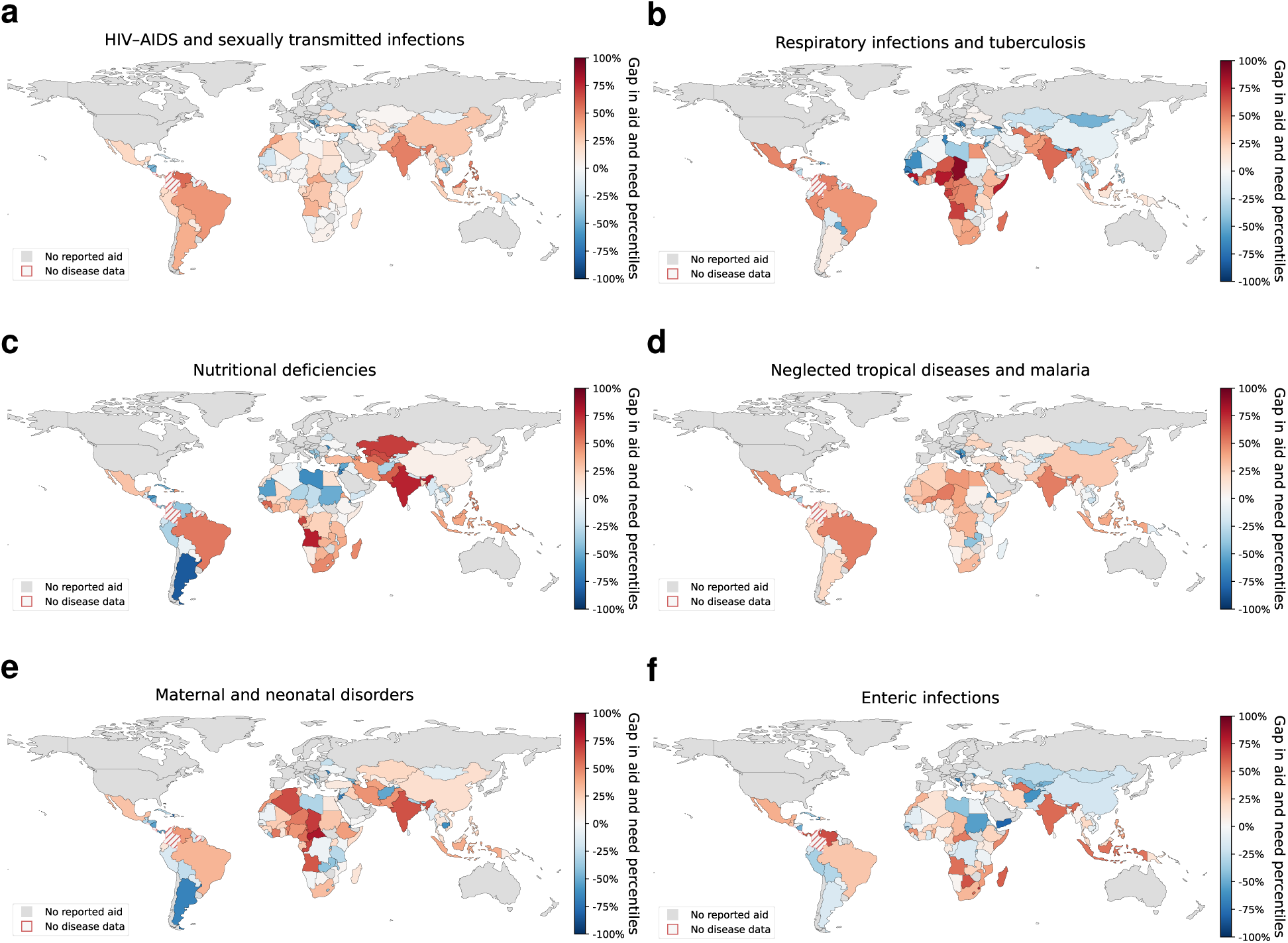
Geographic distribution of aid–burden misalignment (for CMNNDs). The maps show the relative aid–burden misalignment by disease. Here, we refer to “aid–burden misalignment” as the difference between a country’s percentile in disease burden (DALYs per 100,000 population) and its percentile in per capita aid, calculated within income groups. Aid–burden misalignment captures relative disparity within income groups, not necessarily absolute underfunding. In other words, an aid–burden misalignment of 10 percentiles means that the country’s disease burden is 10 percentiles higher than its aid level. Put simply, assuming 100 peer countries, this country would need to climb 10 places in the aid rank to have a similar rank in terms of aid and burden. Due to space, we focus on selected diseases with high overall funding: **a**, HIV/AIDS and STIs; **b**, respiratory infections and TB; **c**, maternal and neonatal disorders; **d**, enteric infections; **e**, NTD and malaria; and **f**, nutritional deficiencies. Red color denotes aid–burden misalignment, where a country’s aid rank is lower than its burden rank. Blue color denotes that a country’s aid rank is higher than the burden rank. Gray indicates no reported aid; red striped areas indicate no disease data. The geographic distribution of aid–burden misalignment for NCDs is shown in Supplementary Figure S2.

## Discussion

Our analysis shows a skew in aid toward communicable diseases, with LDCs and LMICs exhibiting a stronger aid-burden alignment than UMICs in these disease categories. One potential reason for this is that the greater domestic capacity of UMICs to manage disease burden was reflected in donor priorities. HIV/AIDS in particular has consistently received the largest share of global health aid prior to the COVID-19 pandemic, reflecting long-standing prioritization by international donors [38]. Communicable diseases offer clearer, shorter-term impact paths, fitting donors’ preference for vertical, high-visibility results [49]. However, such priorities may displace funding for other important areas of healthcare [50, 51], including NCDs, which continue to receive a disproportionately low share of ODA relative to their growing global burden [52]. Furthermore, NCDs would often involve systemic and multisectoral approaches that would benefit multiple disease categories simultaneously. While the WHO and international donor organizations agreed in 2007 that health system strengthening should become a greater priority [53–55], our analysis indicates that funding disparities still reflect historical priorities, suggesting that general improvements to global health systems in years prior to the COVID-19 pandemic need to continue. However, amid recent cuts to WHO and USAID funding, which threaten progress in HIV, malaria, maternal health, and other disease categories [49, 55, 56], retaining and reallocating scarce resources equitably will become even more difficult. These developments underscore the necessity for timely and comprehensive monitoring of health aid flows to ensure investments align with evolving global health needs.

We found a considerable increase in the funding for respiratory infections and tuberculosis during the COVID-19 pandemic. While emergency funding surged and the introduction of infection control measures may have provided broader benefits for infectious disease control [57], many low-income countries experienced sharp disruptions in routine health services [58, 59], including tuberculosis case detection, HIV testing and treatment initiation, and malaria prevention campaigns [60–63]. Low-income countries would thus have needed an increase in aid for several disease categories to cope with the additional burden of the pandemic. However, aid for diseases unrelated to respiratory infections did not show a significant in- or decrease, suggesting that the pandemic did not trigger a broader shift in funding priorities. Furthermore, our sensitivity analysis revealed that the correlation between disease burden and aid became lower for respiratory infections and tuberculosis during the 2020-2021 period (Supplementary Figures S4, S5, and S6), suggesting that the COVID-19 pandemic exposed and possibly exacerbated inequities in development aid allocation. The impact of the pandemic was especially severe in low-income countries with regions such as sub-Saharan Africa facing a “triple burden” of tuberculosis, HIV, and COVID-19 [64]. Fragile health systems and aid misalignments left underserved populations even more vulnerable and the disparities in health aid and infrastructure contributed to an unequal recovery, which may increase global health inequities in the years following the pandemic.

While HIV/AIDS has consistently received the largest share of ODA prior to the COVID-19 pandemic, ODA distribution is uneven, and our analysis showed substantial geographical disparities in the alignment between HIV/AIDS-related ODA and disease burden. TB, despite its long-standing burden, continues to attract considerable funding, yet eradication remains challenging due to persistent poverty and the lack of an effective vaccine [65]. Notably, the Global Fund’s recent change in allocation methodology, increasing the share of funds for TB in high-burden countries from 18% to 22%, represents a positive shift [66]. At the same time, the burden of NCDs is rising rapidly in many low- and middle-income countries, with increasing prevalence of conditions like cardiovascular diseases and mental disorders [67]. Yet, we found that pronounced aid–burden mis-alignment remains for many NCDs. In sum, our results point to persistent relative misalignment between CMNNDs and NCDs: aid remains misaligned with their respective burden shares, even as NCD-directed ODA has risen over time. This indicates challenges in adapting to slow structural changes.

Our machine learning pipeline offers several strengths. First, it generates a comprehensive, fine-grained dataset of ODA flows across 157 countries and 17 disease categories. As a result, our dataset provides substantially broader coverage across diseases and regions as compared to prior efforts, which typically focus on a limited set of diseases (e.g., HIV/AIDS, TB, and malaria [32–39, 68]) and specific geographical contexts (e.g., certain African countries [32–35]). Second, our pipeline leverages a state-of-the-art, open-source large language model, which enables accurate and consistent classification of health-related ODA. By making our code publicly available, we also allow for seamless adoption by end users across various institutional settings. Third, our machine learning pipeline is scalable, thus allowing for regular updates and timely monitoring of ODA flows. It helps address the delays and resource demands associated with manual coding and strengthen the evidence available for policy-making in global health.

As with others, our study is subject to limitations. First, the textual descriptions of ODA projects vary in quality, specificity, and length, which may introduce bias into the classification process. In addition, a fraction of projects lacked any textual description and could therefore not be categorized by our machine learning pipeline and are excluded from the analysis. For example, diseases such as HIV and TB are linked, therefore many projects may relate to both, and it is challenging to attribute the relative proportion of funding each disease received. Nevertheless, these descriptions provide raw, first-hand data and offer greater granularity compared to earlier efforts, which strengthens the accuracy and depth of our findings. Moreover, previous studies have shown that such data are highly effective for monitoring tasks [69], which corroborates the robustness of our analysis. Second, the reliability of our findings depends on the performance of the underlying machine learning pipeline. In particular, translating non-English descriptions via machine learning may introduce translation error, and classification may conflate the estimated funding gaps. To mitigate this risk, we validated our approach against (i) a conventional keyword-based classification approach and (ii) a manual annotations of 2,060 ODA projects (see Supplementary Material S1), thereby providing evidence for the reliability of our approach. Third, we focus on various causes of disease burden as defined by the Global Burden of Disease study [40, 41] to enable comparability over time but which may limit responsiveness to emerging global health priorities that fall outside the existing taxonomy. Nevertheless, our pipeline can be seamlessly adapted to incorporate new disease categories as needed. Moreover, because some projects are assigned to single primary disease categories, cross-disease interactions and co-benefits (e.g., HIV–TB) can impact estimates of aid–burden misalignment, with the magnitude likely varying by disease area. Projects outside the disease categories (e.g., education, environment, climate) may have health spillovers that we do not attribute to specific diseases, which can shift apparent aid–burden alignment in either direction. Fourth, while we analyze ODA at the disease-category level, we do not disaggregate funding by regions beyond the country level, donors, or funding instruments, which may mask variations in aid effectiveness within categories. We also restrict our analysis to ODA as reported by the Creditor Reporting System of the OECD. CRS provides comprehensive coverage of bilateral and multilateral government donors and includes major institutional and philanthropic actors (such as the Global Fund and the Bill & Melinda Gates Foundation), it has only partial coverage of other private philanthropic contributions; accordingly, our findings primarily reflect government-sourced and large institutional development assistance for health, rather than the full landscape of privately financed global health funding. We track disbursements as reported in the CRS database and do not trace downstream reallocations, which introduces uncertainty in disease-specific estimates and may not fully capture end-use spending relative to the reported allocations. When a project covers multiple disease areas, we apportion funding equally following prior work [24, 27, 32, 37], which may not fully reflect the actual allocation. Fifth, we only measure the alignment between funding needs and aid allocation. However, other drivers in the health system (such as macroeconomic variables) also influence the financial requirements needed to reach specific health targets. Thus, other methods are needed to fully quantify the financial resources required to achieve these targets [70]. Similarly, our analysis focuses on disease burden (as measured by DALYs per 100,000 population) as the primary determinant of funding needs. However, local health system capacity, governance, and infrastructure also influence how funding translates to health outcomes, which are not directly incorporated in this study. Additionally, while DALYs provide a widely used metric to quantify disease burden, this aggregation of severity and prevalence involves inherent value choices that may affect the interpretation of misalignment. Deviations from aid–burden alignment may reflect legitimate priorities (e.g., equity, disease control, or health system strengthening) [42, 71] and should therefore be interpreted as decision-relevant reference points rather than prescriptive allocation criteria.

Addressing the persistent gap between global health needs and ODA requires timely, disease-specific evidence to inform policy decisions. Our machine learning pipeline enables systematic and scalable tracking of the alignment between ODA flows and disease burden and can be regularly updated to reflect new data and emerging health priorities. Our approach thereby supports more targeted, equitable allocation of ODA by aligning funding with health needs and improving health outcomes worldwide.

## Methods

### Data

We obtained data on ODA projects from the Development Cooperation Directorate of the OECD, which provided us with raw data from the Creditor Reporting System (CRS) [44]. The CRS tracks international ODA projects and is considered the most comprehensive data source on global development aid [72]. In particular, the coverage of the CRS is more comprehensive compared to other datasets on health-related aid, which are often limited to specific diseases, geographic regions, or lack granular disease categorizations. We focus on donor contributions (ODA disbursements) and not domestic government or household health spending.

The ODA projects in the CRS cover various forms of development assistance, including financial grants, development loans, and equity investments made by donor organizations. Projects are reported directly by donor organizations at the close of each year in which the projects are active. Each project entry contains a textual description that describes the objectives, implementation, and outcomes of the project (see Supplementary Table S2 for examples). We track disbursements as reported in the CRS database and cannot trace subsequent downstream reallocations.

### Machine Learning Pipeline

We developed a machine learning pipeline to track ODA flows for public health using a state-of-the-art, open-source large language model (LLM). Our pipeline consists of three steps: In step 1, we translate and preprocess the textual descriptions of ODA projects (*translation and preprocessing*). In step 2, we classify the ODA projects into 17 disease categories using a pre-trained LLM, which is guided by a tailored prompt (*disease classification*). Finally, in step 3, we validate our approach against (i) a conventional keyword-based classification approach and (ii) manual annotations of ODA projects (*validation*).

#### Step 1: Translation and preprocessing

Our dataset includes all ODA projects between 2000–2022 (∼4.7 million). Following prior work [69, 70], we preprocessed the data by identifying the languages of the project descriptions using the `langid` and `langdetect` libraries [73, 74], and then translated all non-English descriptions into English using the Google Translate API [75]. We removed all invalid entries, such as projects with missing textual descriptions, missing funding amounts, or negative funding amounts. Finally, we concatenated the translated project title, long description, and short description into a single text field, which was then input into the machine learning pipeline. To compute per capita aid flows, we used population statistics from the World Bank [76]. The initial dataset contained 4,726,638 projects. After filtering out invalid entries (i.e., 1,012,703 projects; 21.4%), the final cleaned dataset included 3,713,935 projects. The ODA project preprocessing and classification flow is illustrated in Supplementary Figure S10.

#### Step 2: Disease classification

We classified disease burden into 17 categories adapted from the Global Burden of Disease (GBD) cause hierarchy. Specifically, we selected all Level 2 causes from (i) CMNNDs and (ii) NCDs. We did not include injury-related categories and the residual Level 2 buckets (“other infectious diseases”, “other non-communicable diseases”) in the taxonomy because they have received comparatively little focus in health aid initiatives, and their heterogeneity limits interpretability and decision relevance in the context of development assistance for health. The resulting 17-category taxonomy is provided in Supplementary Table S1.

The classification method assigns multiple labels to a project when applicable, i.e., a project may belong to more than one disease category. For example, a project addressing both HIV and malnutrition is categorized under both “HIV/AIDS and STIs” and “nutritional deficiencies”. Since the CRS database does not report within-project allocations to specific disease areas, we distribute the funding equally among the applicable categories to prevent double-counting of the project funding, consistent with prior work [24, 27, 32, 37]. In the aforementioned example, 50% of the project funding would be allocated to “HIV/AIDS and STIs” and 50% to “nutritional deficiencies”. Similarly, if an aid activity extends over multiple years, it is reported on a pro-rata basis per year, with the proportional aid disbursement attributed to the corresponding years. This approach ensures that each project’s total funding is accounted for only once, while accurately reflecting that development aid projects can pursue multiple objectives.

For the classification task, we used Meta-Llama-3.1-70B-Instruct-Turbo [77] via the Together AI API [78]. This LLM is a state-of-the-art, open-source model, which, at the time of our analysis, ranked among the top-performing LLMs across various benchmarks [79]. We also experimented with other LLMs, including GPT-4, Claude Sonnet and Haiku, Mistral 7B, other Llama 3.1 variants (i.e., 70B-Instruct and 8B-Instruct). Eventually, our choice to select Llama-3.1-70B was based on three key considerations: (i) accuracy in structured classification tasks, (ii) scalability in terms of computational costs to enable a large-scale analysis of millions of aid projects, and (iii) consistent classification across varying project description lengths and complexities.

Using this LLM, we then classified the projects based on their textual descriptions into the 17 disease categories from above using a tailored prompt. The prompt design followed best practices in prompt engineering and prior research [80–82], and included a task description, a list of possible labels, and the description of the ODA project to be classified. The model was instructed to adopt a conservative classification strategy to reduce false positives, using fallback categories (“Other” and “General Health”) where applicable. Additional implementation details, including the full prompt and model hyperparameters, are provided in Supplementary Material S3 and Supplementary Material S4, respectively.

#### Step 3: Validation

To assess the accuracy and reliability of our ML approach, we employed a dual validation strategy. First, we benchmarked our approach against a conventional keyword-based classification method, which is interpretable, but unable to capture complex semantics as effectively as our LLM approach. The conventional keyword-based approach, inspired by [36, 39], relies on the predefined terms from our disease categories (see Supplementary Material S1). We then searched for exact matches (case-insensitive) within the title, long description, and short description of each project. Again, aid projects were assigned to one or more categories based on the matches. Projects without keyword match were classified as “Other”. Overall, the keyword-based approach shows large agreement with the results from our machine learning approach. However, there are a few exceptions, which we identified in a manual assessment as issues with the keyword-based search, thereby again highlighting the benefits of our machine learning approach.

Second, we manually annotated 2,060 aid projects and compared the labels to the LLM-generated classifications (see Supplementary Material S1); however, this approach lacks the scalability of our machine learning pipeline. Again, we find strong agreement, with an overall accuracy of 0.93 (micro-averaged *F*_1_-score: 0.94; macro-averaged *F*_1_-score: 0.93) and robust performance across disease categories (both precision and recall *>* 0.9 for the majority of disease categories).

### Analysis

For our analysis of how disease-specific development aid aligns with disease burden, we use disability-adjusted life years (DALYs) to measure disease burden [83, 84]. A DALY combines years of life lost from premature death and years lived with disability. The DALY estimates originate from the Global Burden of Disease study, which systematically quantifies health loss due to diseases across 204 countries [85]. The GBD study provides annual disease burden estimates from 1990 to 2021 and uses standardized estimation approaches to ensure that disease burden metrics can be meaningfully compared across locations and time periods [86]. We then matched the aid and disease burden data at the country level (using ISO-3 country codes). Our ODA data spans 2000–2022; however, comparisons with disease burden are limited to the period 2000–2021, which was the latest available data from the Global Burden of Disease study at the time of our analysis.

To assess whether disease burden aligns with the volume of health-related ODA, we computed Spearman’s rank correlation coefficient. Low correlation may indicate disparities in aid allocation, potentially reflecting historical or donor-driven priorities, and may signal cases where disease burden is high but aid is relatively low, that is, aid–burden misalignment. All statistical tests are based on two-sided Spearman’s rank correlation tests. We selected Spearman’s method over Pearson’s to account for potential non-linear relationships and non-normal data distributions. Correlations were computed separately for each disease category and for each country income group. Income group classifications follow the OECD framework. Our analysis included least-developed countries (LDCs), lower-middle-income countries (LMICs), and upper-middle-income countries (UMICs), but excluded more advanced developing countries and territories (MADCTs) and other low-income countries due to limited data coverage. We further excluded microstates (e.g., Vatican City, Nauru, Tuvalu, etc.), as their small populations can result in disproportionately high per capita aid volumes, thereby distorting aid–burden misalignment estimates. We interpreted correlation coefficients according to established guidelines in statistical literature [87, 88].

To identify countries with aid–burden misalignment, we computed a disease-specific misalignment metric. This metric captures the relative position of each country within its income group by comparing disease burden and aid received. For each disease, countries are ranked based on two measures: (i) per capita aid received (aid rank percentile) and (ii) disease burden, measured as DALYs per 100,000 population (burden rank percentile). Aid–burden misalignment is calculated as the difference between these two percentiles. Positive values indicate countries with higher disease burden, but lower aid levels compared to their peers. This metric is designed to capture relative disparities, rather than absolute funding adequacy. It is a descriptive, rank-based indicator and should not be interpreted as a prescriptive criterion for funding allocation. For example, an aid–burden misalignment of +10 percentiles indicates that a country’s disease burden rank is 10 percentiles higher than its aid rank among countries in the same income group. Countries were ranked within their respective income groups to account for differences in economic capacity, which affect the ability to respond to health challenges. This reflects that an equivalent disease burden will create a relatively greater strain on countries with fewer domestic resources. This approach follows established practices in global health financing [89], and aligns with the resource allocation frameworks of major international health financing institutions such as the Global Fund and Gavi, the Vaccine Alliance [45, 90–93]. The rationale for taking into account economic capacity in health burden assessment is well-established [94–96], with the Lancet Commission on Investing in Health and the WHO Commission on Macroeconomics and Health emphasizing the importance of economic context for both burden assessment and resource allocation [97, 98].

### Sensitivity analysis

To evaluate the sensitivity of our results to key modeling choices, we conducted several supplementary analyses. First, we studied a different metric, namely, disease incidence as an alternative to DALYs (Supplementary Figure S3). Second, to assess the impact of the COVID-19 pandemic on the alignment between aid and burden, we separately analyzed the years 2020–2021 (Supplementary Figures S4, S5, S6). Third, to account for possible changes in funding priorities during the COVID-19 pandemic among donors, we excluded data from the years 2020 and 2021 for respiratory infections and TB from our analysis (Supplementary Figures S7, S8, S9). In call cases, we recomputed disease-specific per capita aid and Spearman correlations with burden using the same procedure as in the main analysis.

### Robustness checks

To assess the robustness of our disease-classification pipeline, we re-ran the classification on a randomly sampled 1% of projects with different random seeds and an alternative LLM, summarized agreement using Cohen’s kappa (*κ*) and the label consistency rate (LCR), and found consistently high run-to-run and inter-model agreement. In addition, we conducted targeted robustness analyses for specific failure modes, including (i) a chunk-based variant of the pipeline applied to projects with long descriptions, (ii) a negation-focused validation set of projects containing statements in which disease keywords occur in a negated context (e.g., “no tuberculosis”, “not HIV”, “excluding malaria”), and (iii) a separate set of short, non-English, and semantically ambiguous project descriptions, where predictions from the pipeline were compared against new manual labels. Across these subsets, overall accuracy and micro-averaged *F*_1_-scores remained high and closely matched the main validation results (Supplementary Materials S2).

## Data availability

All generated data to replicate our analyses is available via https://osf.io/ce3q2/. The raw aid data is available via the Creditor Reporting System from the OECD (https://data-explorer.oecd.org/). The disease burden data is available from the Global Burden of Disease study [40, 41].

## Code availability

All code to replicate our analyses is available via https://github.com/forsterkerstin/tracking-health-aid-disparities.

## Acknowledgments

S.F. acknowledges funding from the Swiss National Science Foundation (SNSF) via Grant 186932.

## Author contributions

F.S., K.F., and S.F. conceptualized the machine learning pipeline. F.S. and K.F. performed data analysis and visualized the results. F.S., K.F., N.B., and S.F. wrote the paper. All the authors contributed to the interpretation of the results and approved the final manuscript.

## Supplementary Tables

**Table S1:**
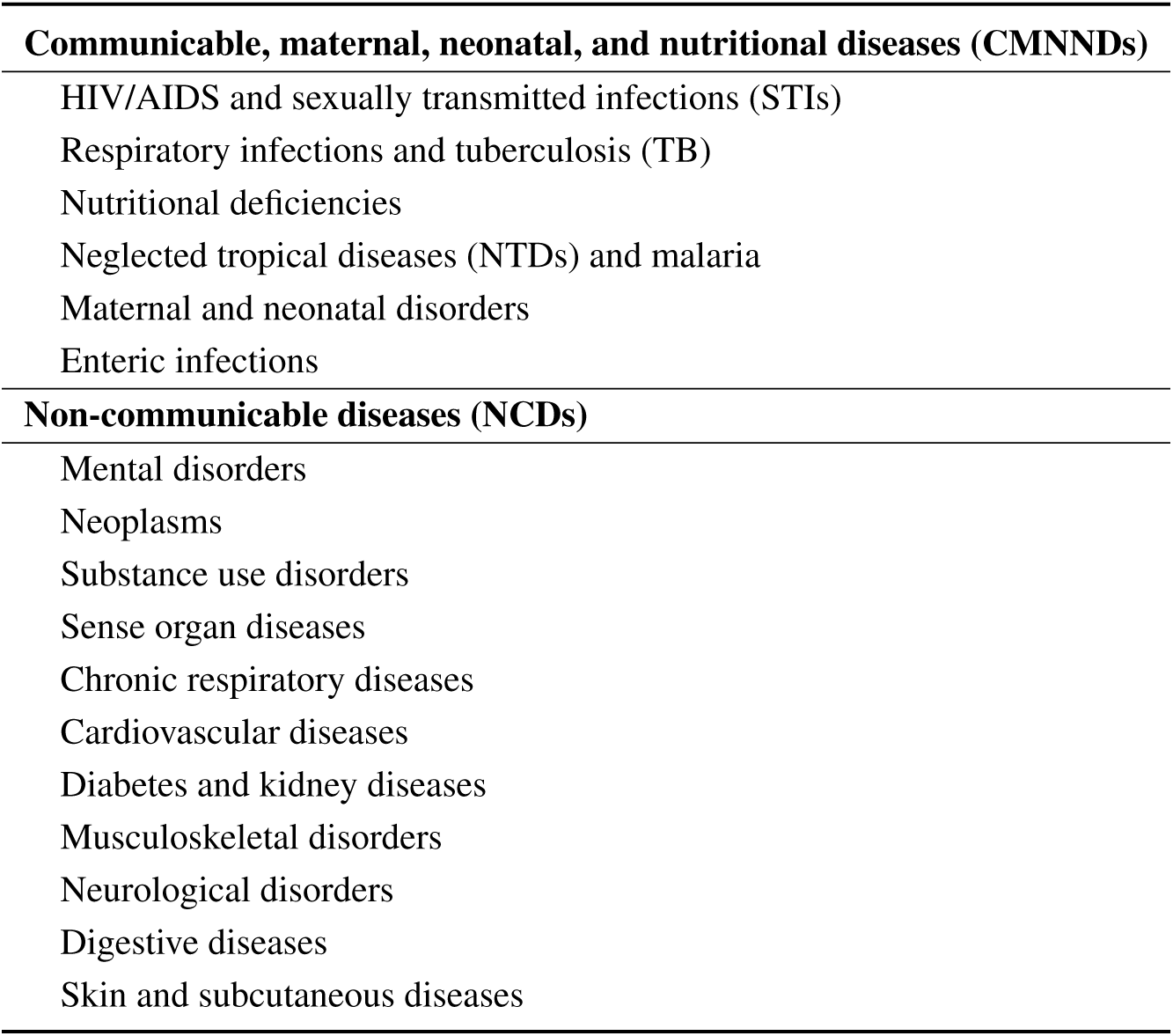
Classification of causes of disease burden. The categorization is based on the causes of disease burden in the Global Burden of Disease study [40, 83]. Specifically, we selected all Level 2 causes from (i) CMNNDs and (ii) NCDs. We did not include injury-related categories and the residual Level 2 buckets (“other infectious diseases”, “other non-communicable diseases”) in the taxonomy because they have received comparatively little focus in health aid initiatives and their heterogeneity limits interpretability and decision relevance in the context of development assistance for health.

| **Communicable, maternal, neonatal, and nutritional diseases (CMNNDs)** |
| --- |
| HIV/AIDS and sexually transmitted infections (STIs) |
| Respiratory infections and tuberculosis (TB) |
| Nutritional deficiencies |
| Neglected tropical diseases (NTDs) and malaria |
| Maternal and neonatal disorders |
| Enteric infections |
| **Non-communicable diseases (NCDs)** |
| Mental disorders |
| Neoplasms |
| Substance use disorders |
| Sense organ diseases |
| Chronic respiratory diseases |
| Cardiovascular diseases |
| Diabetes and kidney diseases |
| Musculoskeletal disorders |
| Neurological disorders |
| Digestive diseases |
| Skin and subcutaneous diseases |

**Table S2:** Example ODA project with metadata and project description.

| Parameter | Details |
| --- | --- |
| Year | 2017 |
| Donor | UNDP |
| Recipient country | Angola |
| Income group | LDCs |
| Project title | Reducing the Burden of HIV/AIDS |
| Project description | *Strengthened the institutional capacity of the national HIV&AIDS programme, National health information system, epidemiological surveillance and expansion of counseling and testing centres* |
| Year | 2021 |
| Donor | Global Alliance for Vaccines and Immunization |
| Recipient country | Uganda |
| Income group | LDCs |
| Project title | New vaccine support (NVS) |
| Project description | *Vaccine against rotavirus, which causes diarrhoea.* |
| Year | 2020 |
| Donor | France |
| Recipient | Niger |
| Income group | LDCs |
| Project title | P209 - Fonds Muskoka - Niger - ONU femmes |
| Project description | *The French Muskoka Fund (FFM) has been operating since 2011 in West and Central Africa to accelerate the reduction of maternal and infant mortality and improve reproductive, sexual, Maternal, Neonatal, Child, Adolescent and Nutrition (RMNCIA-N) health outcomes.* |
| Year | 2021 |
| Donor | Spain |
| Recipient country | Sudan |
| Income group | LDCs |
| Project title | Vaccine support for covid-19 |
| Project description | *Donation via COVAX of 2.325.600 doses of Janssen COVID-19 vaccines to Sudan.* |

## Supplementary Figures

**Fig. S1.**
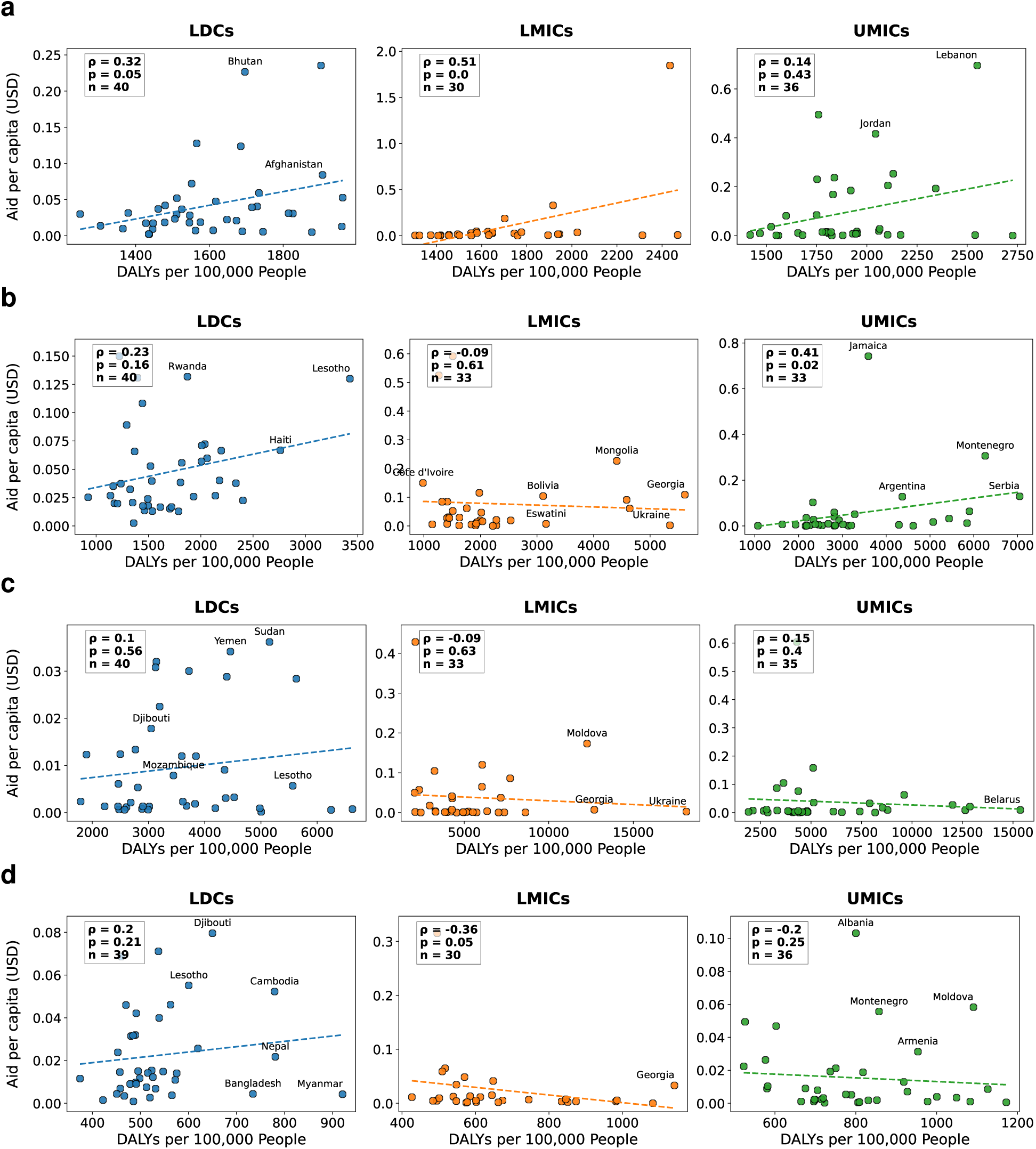
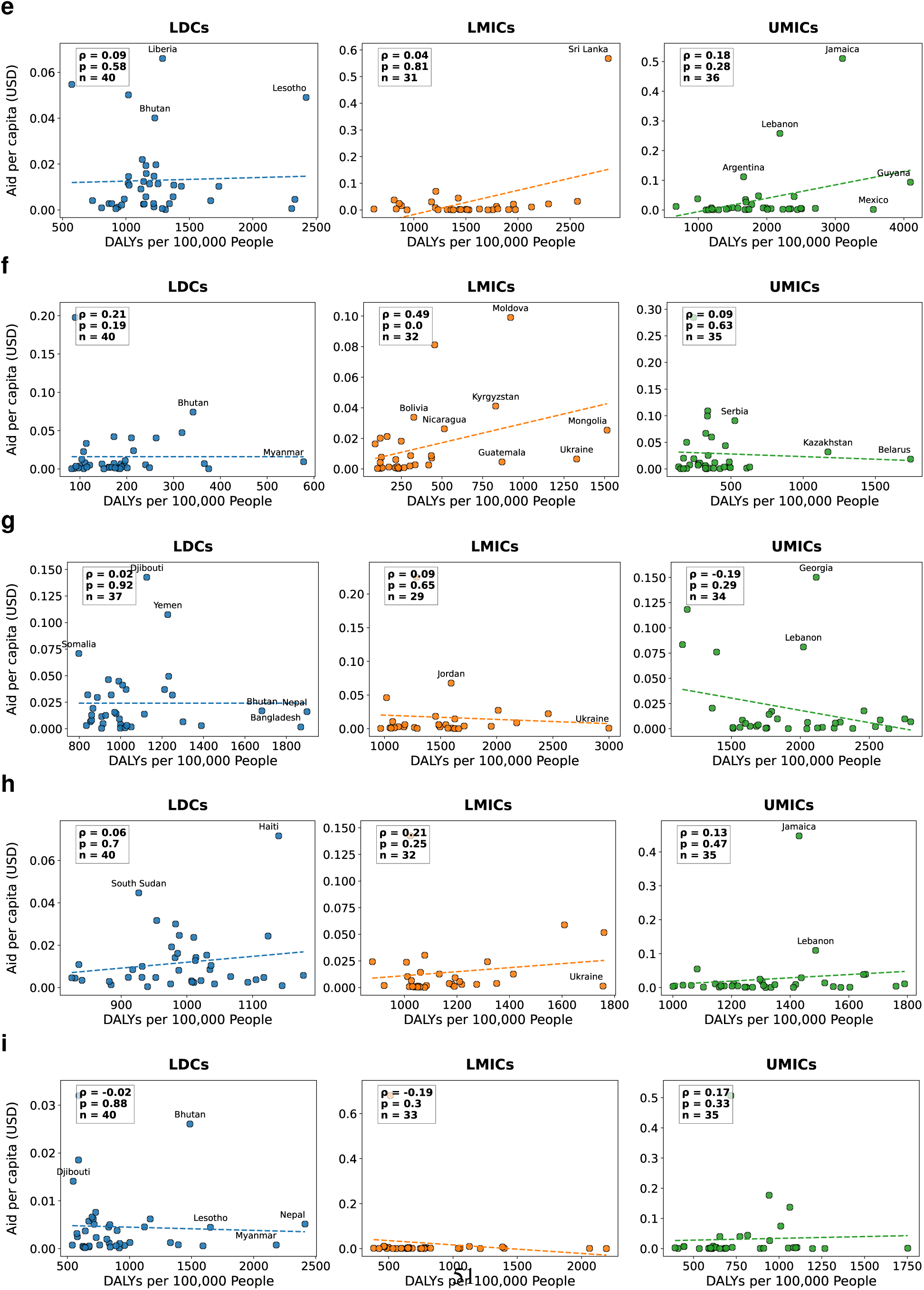
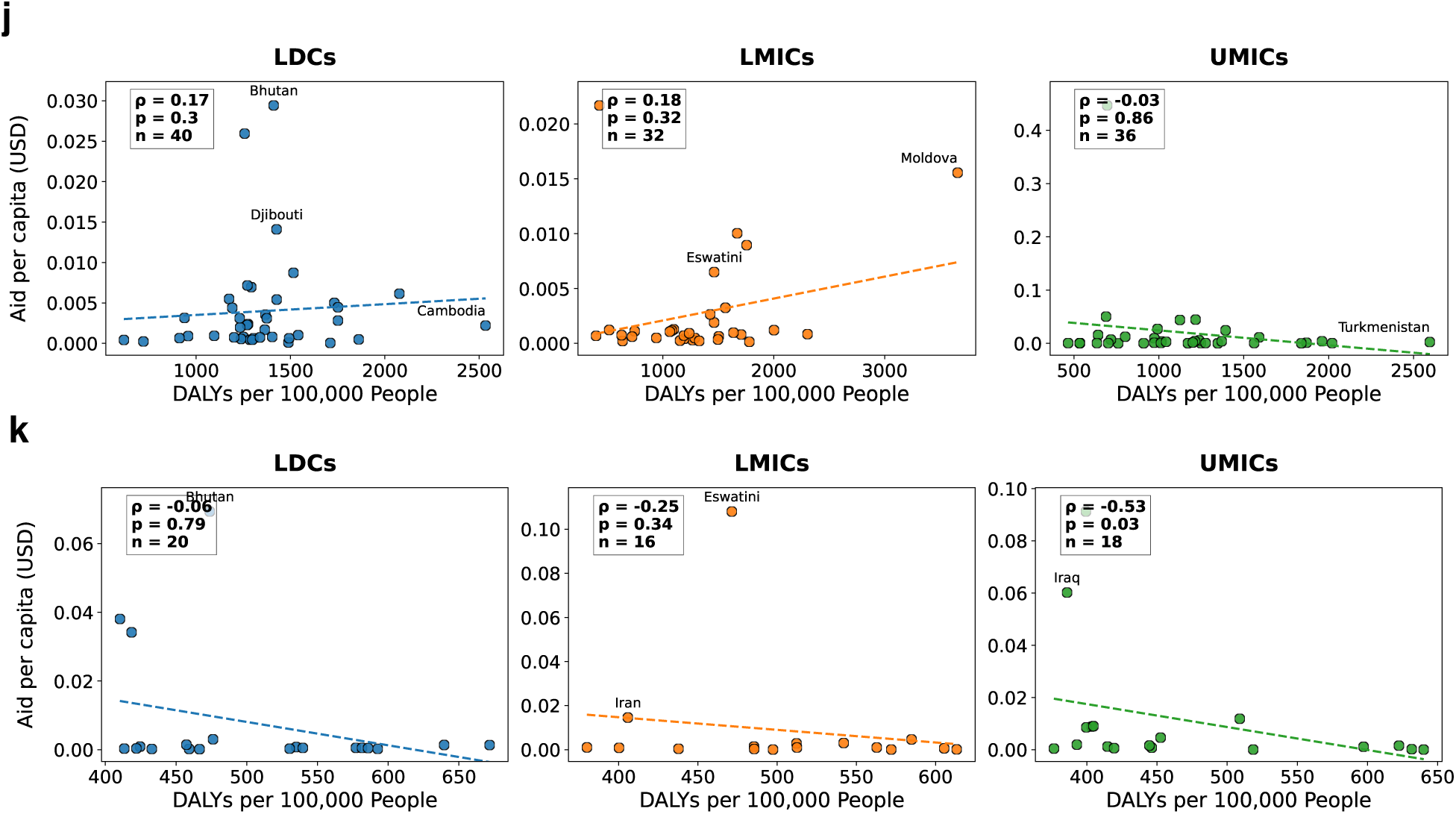
Funding disparities at the country level (for NCDs). The plots compare the disease burden (*x*-axis, measured by DALYs per 100,000 population, accumulated for 2000–2021) against the aid per capita (*y*-axis, accumulated for 2000–2021). Each point represents a single country, which allows to identify countries with high disease burden but comparatively low aid and thus to reveal aid–burden misalignment. For each disease, countries/plots are grouped by country income groups: least developed countries (LDCs) (*n* = 40 countries), lower-middle-income countries (LMICs) (*n* = 32) and upper-middle-income countries (UMICs) (*n* = 35). The figure show: **a**, mental disorders; **b**, neoplasms; **c**, cardiovascular diseases; **d**, sense organ diseases; **e**, diabetes and kidney diseases; **f**, substance use disorders; **g**, musculoskeletal disorders; **h**, neurological disorders; **i**, chronic respiratory diseases; **j**, digestive diseases; and **k**, skin and subcutaneous diseases. Each plot reports Spearman’s correlation coefficient (*ρ*) and the *p*-value from a two-sided Spearman rank test. The dashed line is the fitted trend line from an ordinary least squares regression.

**Fig. S2.**
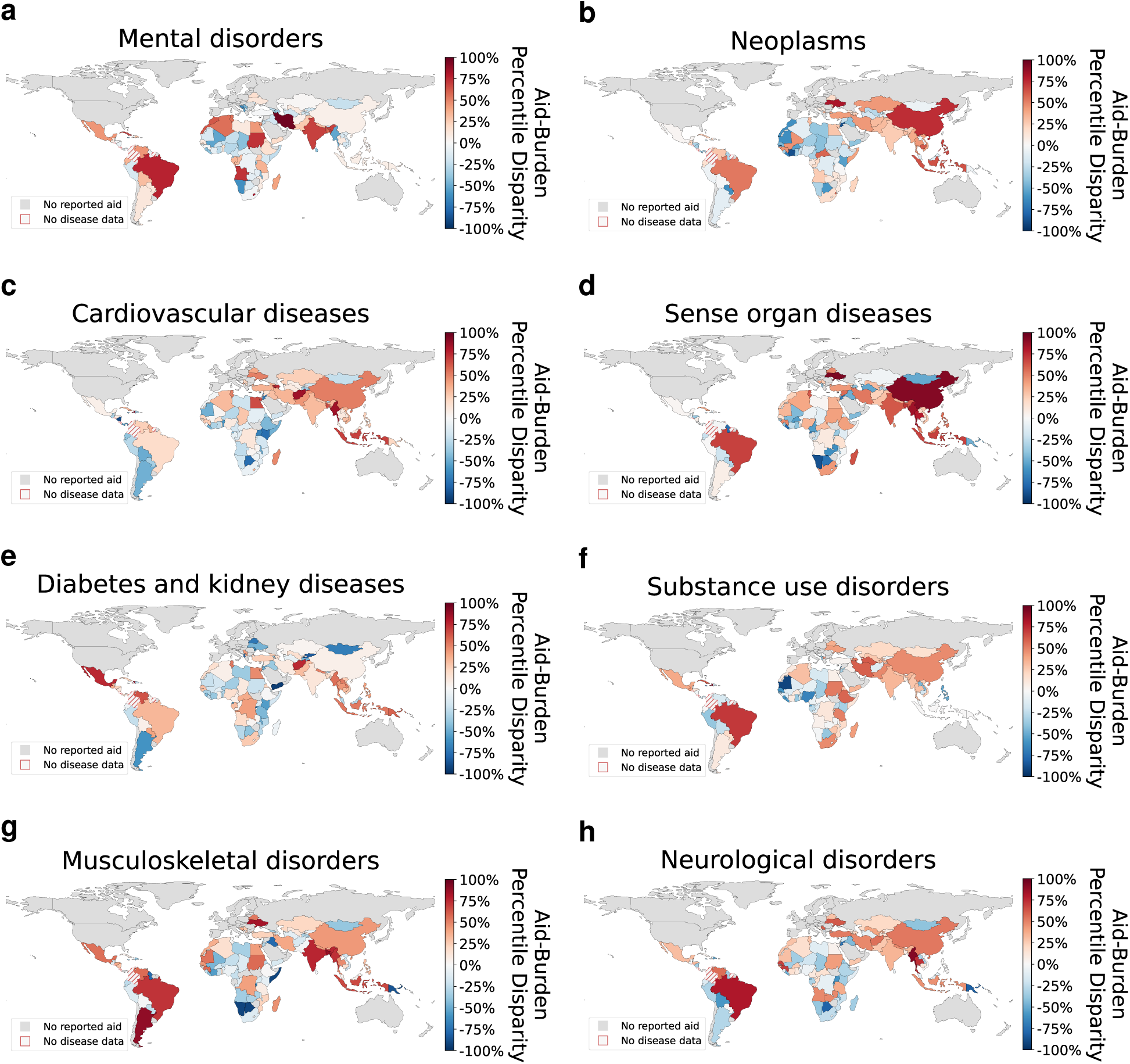
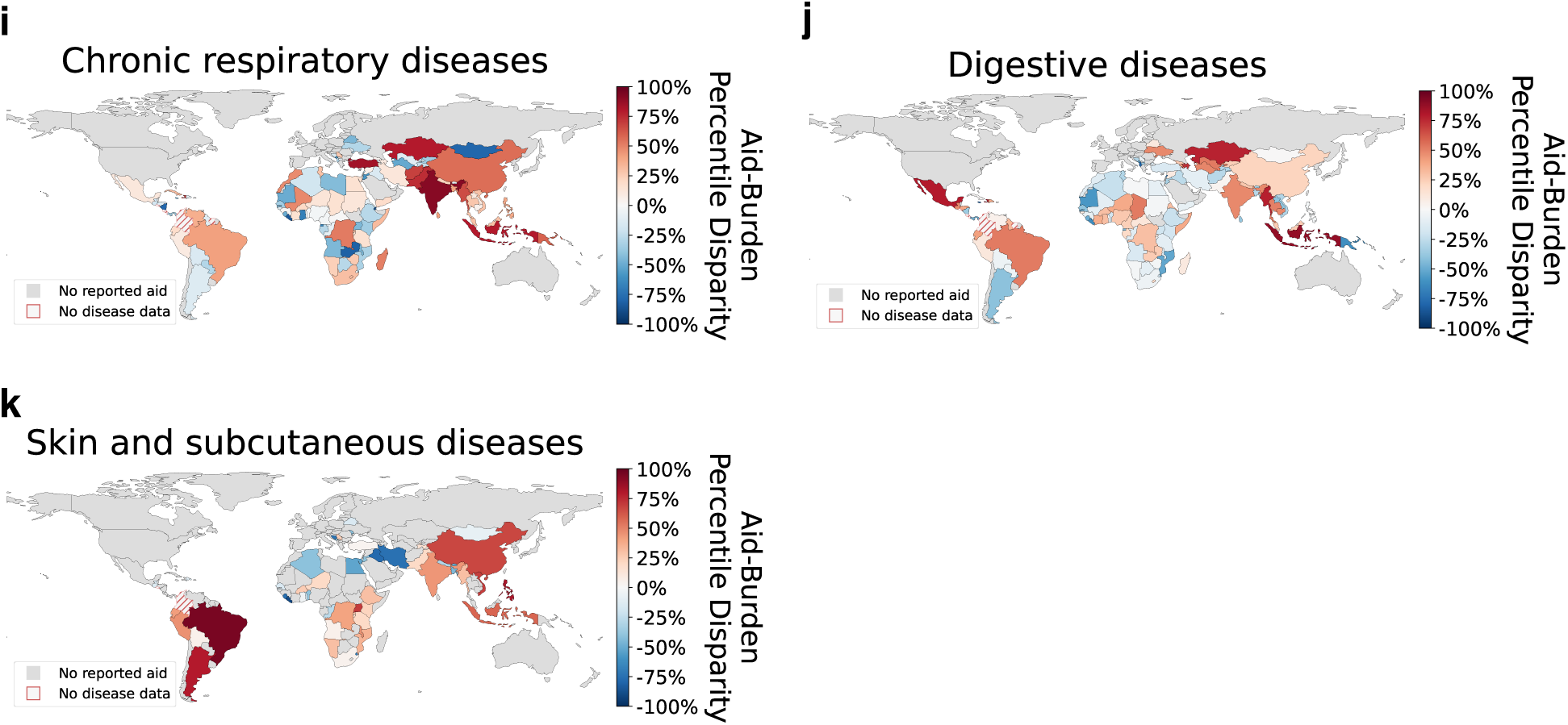
Geographic distribution of aid–burden misalignment (for NCDs). The maps show the relative aid–burden misalignment by disease. Here, we refer to “aid–burden misalignment” as the difference between a country’s percentile in disease burden (DALYs per 100,000 population) and its percentile in per capita aid, calculated within income groups. In other words, an aid–burden misalignment of 10 percentiles means that the country’s disease burden is 10 percentiles higher than its aid level. Put simply, assuming 100 peer countries, this country would need to climb 10 places in the aid rank to have a similar rank in terms of aid and burden. Red color denotes aid– burden misalignment, where a country’s aid rank is lower than its burden rank. Blue color denotes that a country’s aid rank is higher than the burden rank. Gray indicates no reported aid; red striped areas indicate no disease data.

**Fig. S3.**
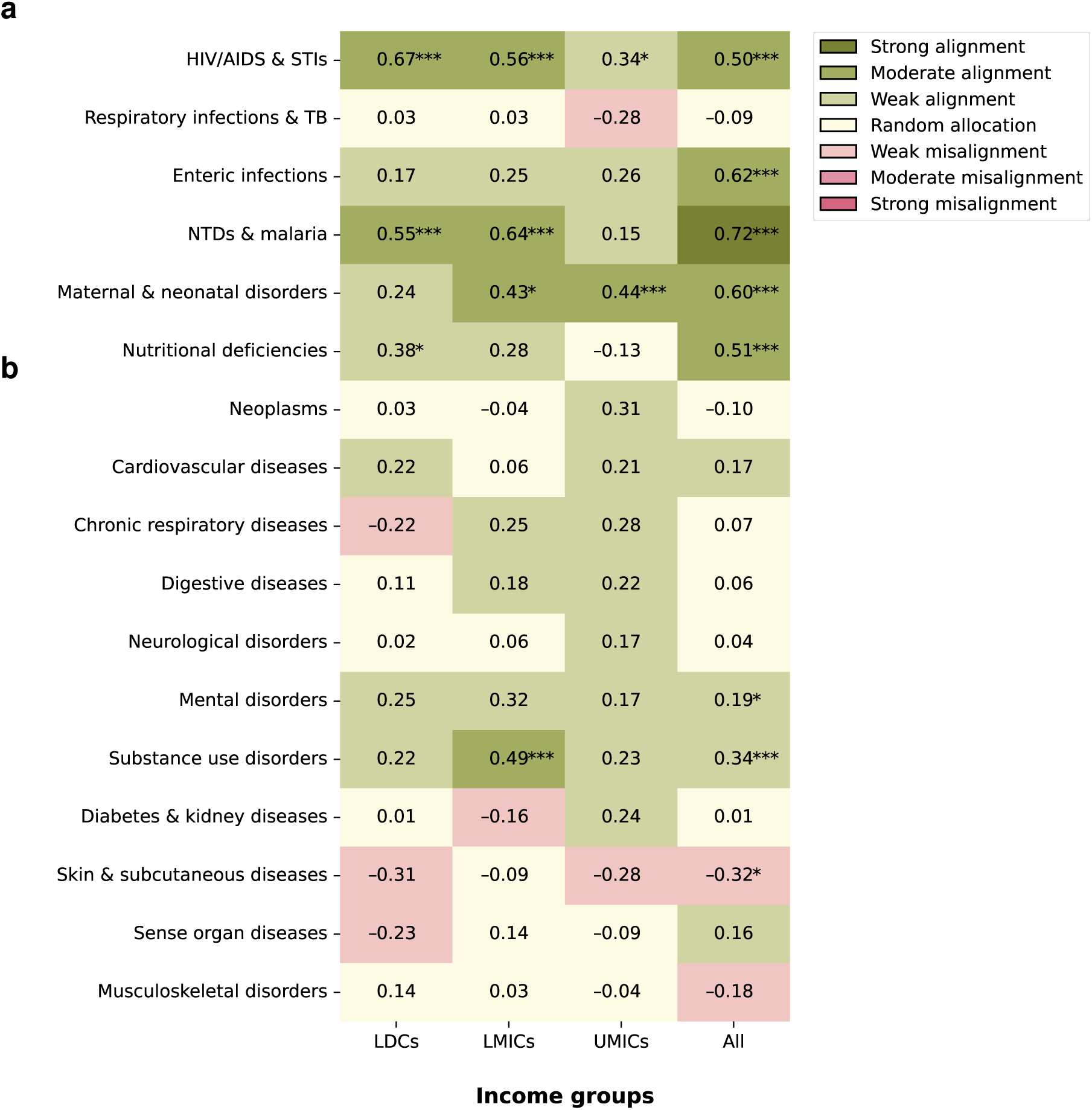
Correlation between disease-specific aid and incidence across income groups. Shown are Spearman’s rank correlation coefficients between per capita aid (accumulated over the time period 2000–2021) and disease incidence (as reported in the Global Burden of Disease study) across country income groups: least developed countries (LDCs), lower-middle-income countries (LMICs), upper-middle-income countries (UMICs), and all countries combined. **a**, The top six rows report CMNNDs, which generally show positive correlations across most income groups. **b**, In contrast, NCDs (bottom eleven rows) exhibit more heterogeneity in the correlations. CMNNDs (top six rows) predominantly show positive correlations across most income groups, while NCDs (bottom eleven rows) display more varied patterns. Green indicates positive and red negative correlations. Asterisks denote statistically significant correlations based on a two-sided Spearman rank test (^∗^ *p <* 0.05; ^∗∗^ *p <* 0.01; ^∗∗∗^ *p <* 0.001). Legend interpretation: −1 to −0.7: strong misalignment of health aid with disease burden; −0.7 to −0.3: moderate misalignment; −0.3 to −0.1: weak misalignment, −0.1 to 0.1: indicative of random allocation; 0.1 to 0.3: weak alignment, 0.3 to 0.7, moderate alignment; and 0.7 to 1.0 strong alignment. Abbreviations: STI, sexually transmitted infections; TB, tuberculosis;

**Fig. S4.**
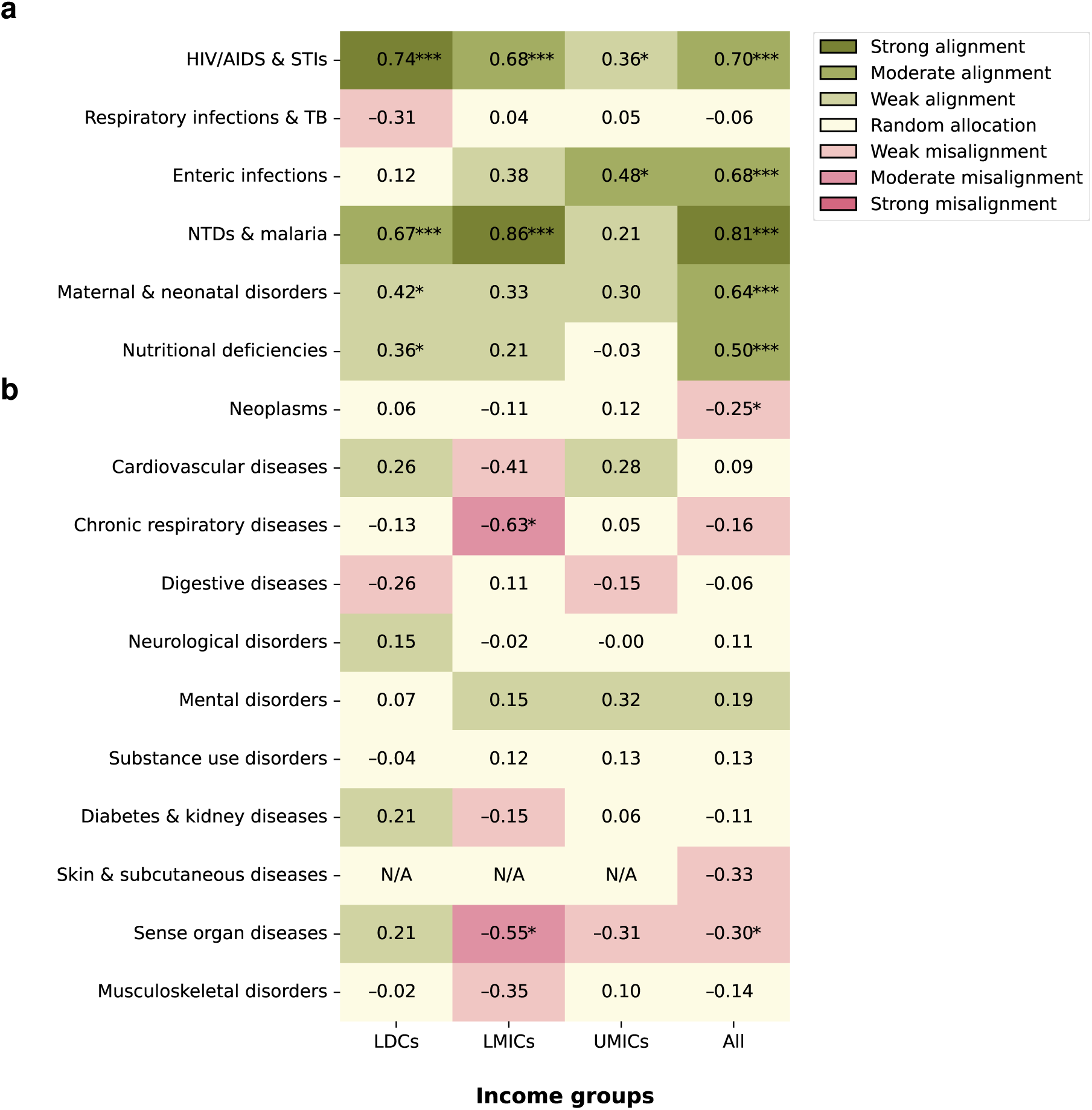
Correlation between disease-specific aid and burden across income groups (for 2020– 2021). Shown are Spearman’s rank correlation coefficients between per capita aid (accumulated over the time period 2020–2021) and disease burden across country income groups: least developed countries (LDCs), lower-middle-income countries (LMICs), upper-middle-income countries (UMICs), and all countries combined. **a**, The top six rows report CMNNDs. **b**, The bottom 11 rows show NCDs. Green indicates positive and red negative correlations. Asterisks denote statistically significant correlations based on a two-sided Spearman rank test (^∗^ *p <* 0.05; ^∗∗^ *p <* 0.01; ^∗∗∗^ *p <* 0.001). Legend interpretation: −1 to −0.7: strong misalignment of health aid with disease burden; −0.7 to −0.3: moderate misalignment; −0.3 to −0.1: weak misalignment, −0.1 to 0.1: indicative of random allocation; 0.1 to 0.3: weak alignment, 0.3 to 0.7, moderate alignment; and 0.7 to 1.0 strong alignment. Abbreviations: STI, sexually transmitted infections; TB, tuberculosis;

**Fig. S5.**
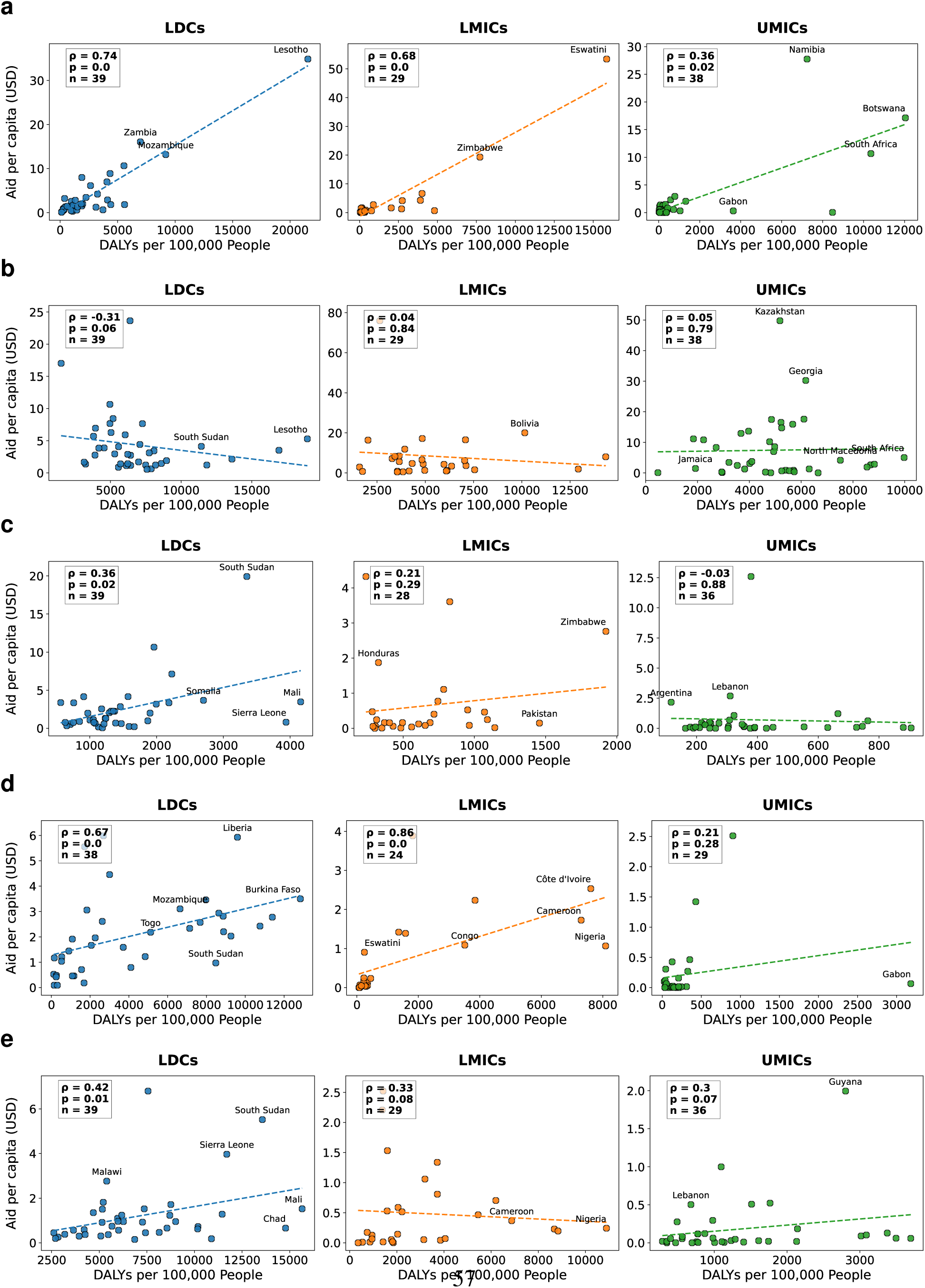
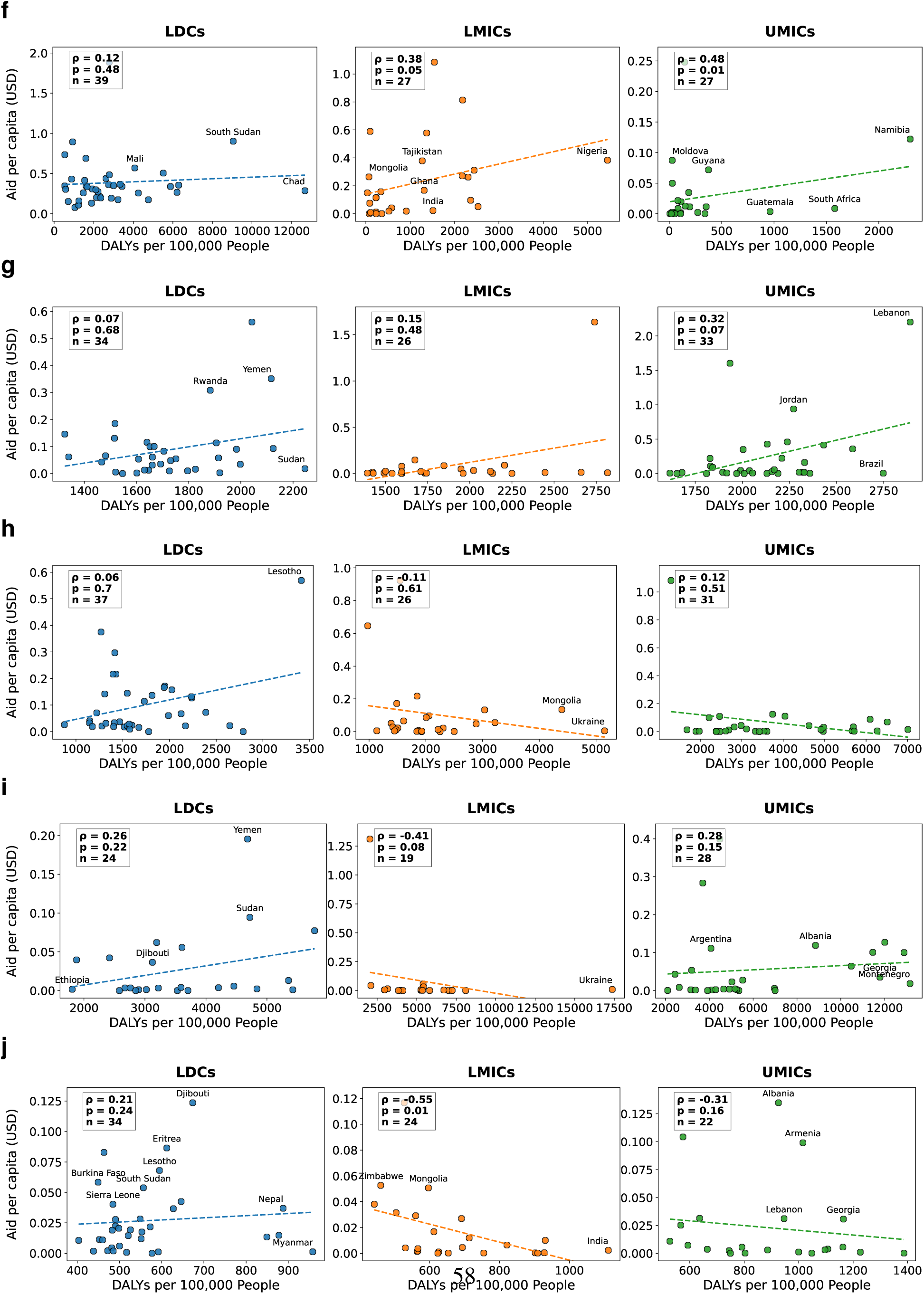
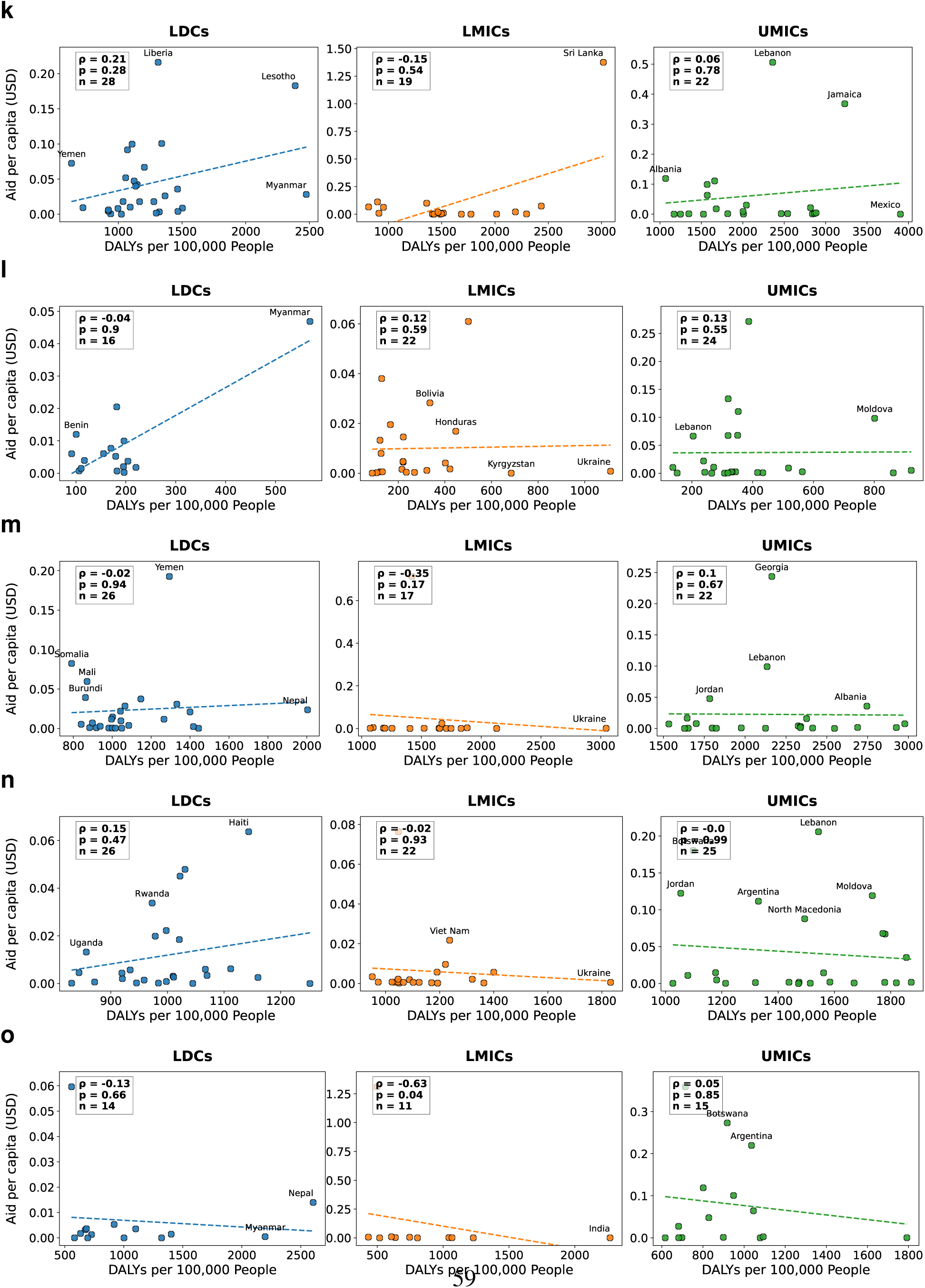
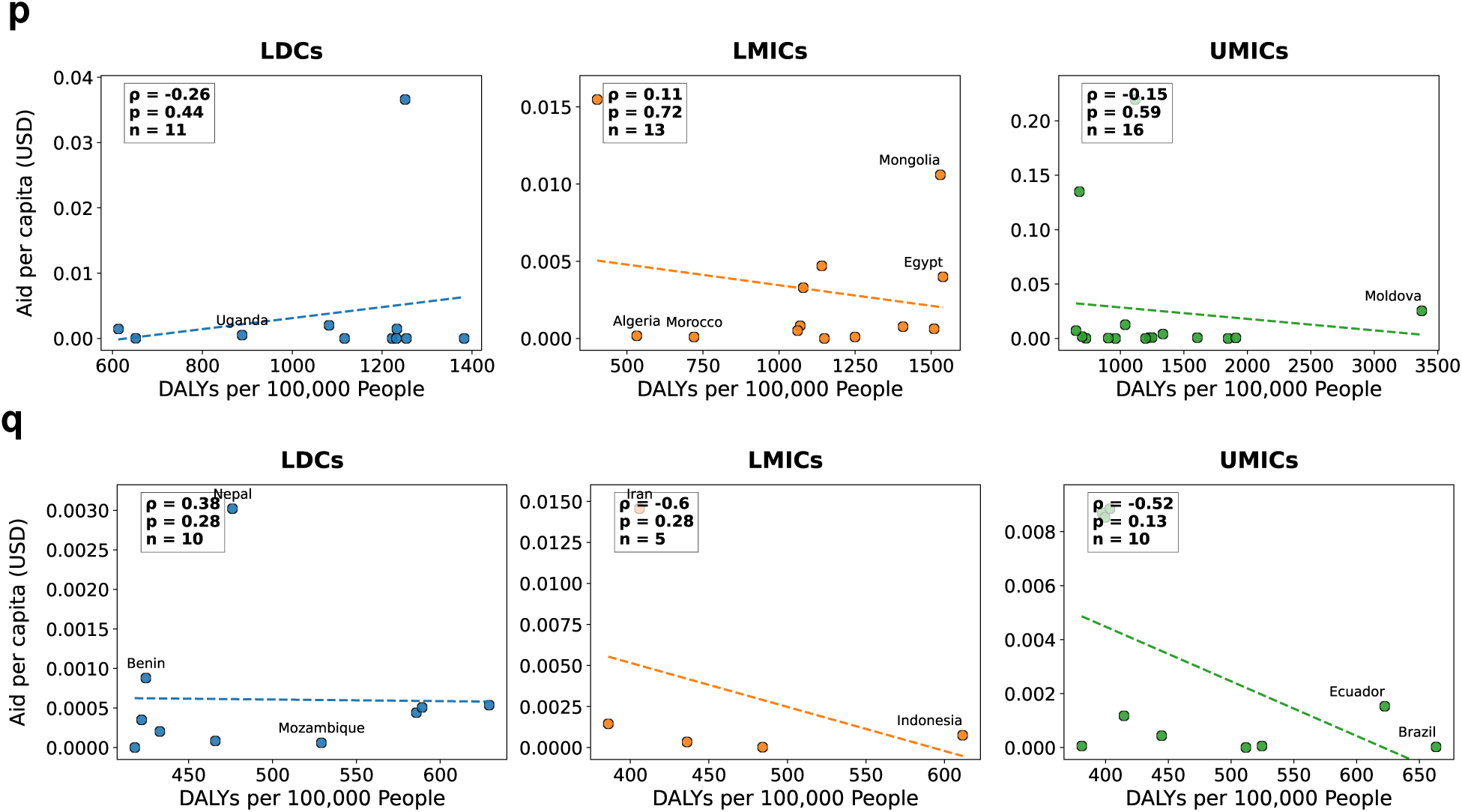
Funding disparities at the country level (for 2020–2021). The figure is analogous to Figure 8 in the main paper, except for the different time frame. The figure shows: **a**, HIV/AIDS and sexually transmitted infections; **b**, respiratory infections and tuberculosis; **c**, nutritional deficiencies; **d**, neglected tropical diseases and malaria; **e**, maternal and neonatal disorders; **f**, enteric infections; **g**, mental disorders; **h**, neoplasms; **i**, cardiovascular diseases; **j**, sense organ diseases; **k**, diabetes and kidney diseases; **l**, substance use disorders; **m**, musculoskeletal disorders; **n**, neurological disorders; **o**, chronic respiratory diseases; **p**, digestive diseases; and **q**, skin and subcutaneous diseases.

**Fig. S6.**
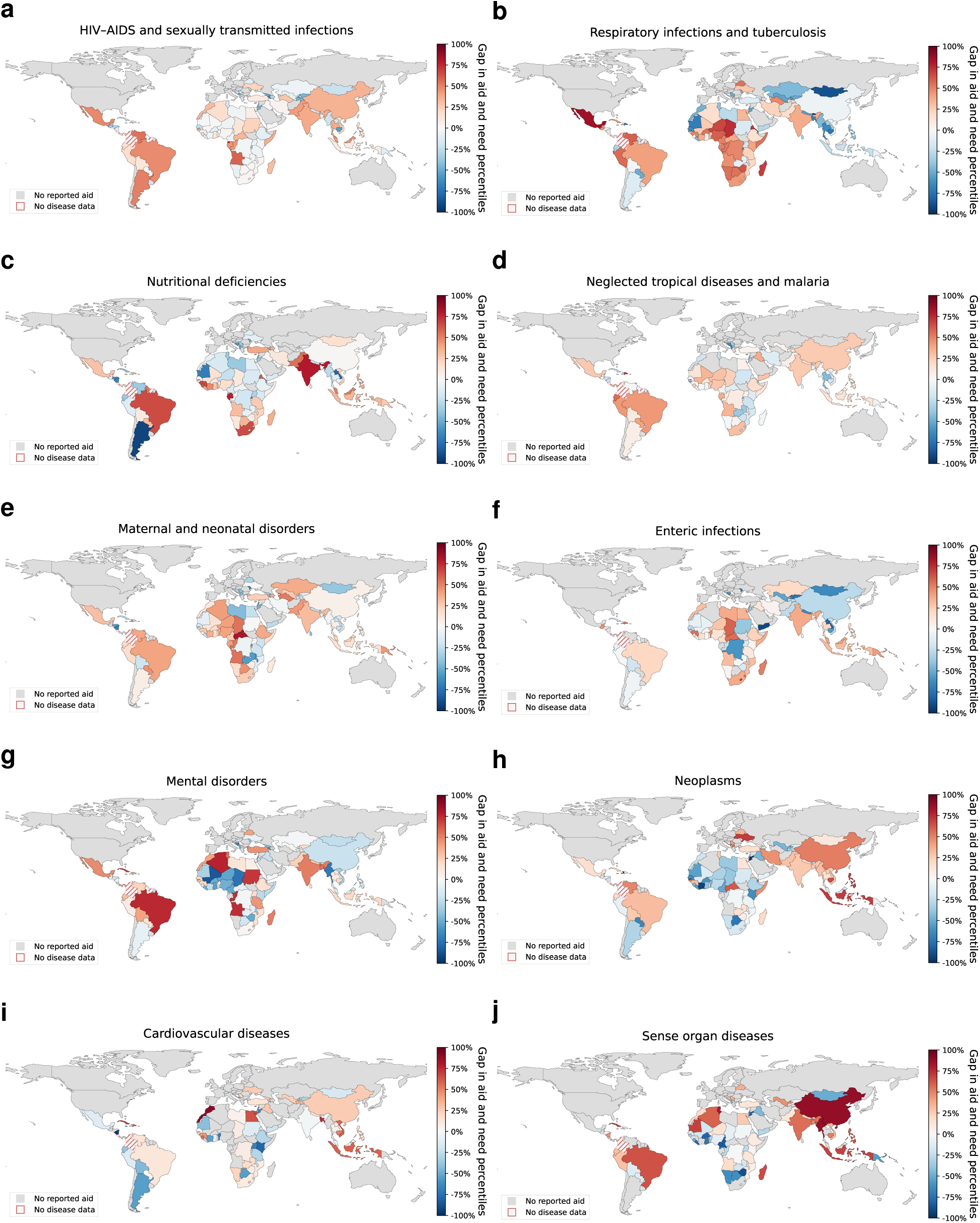
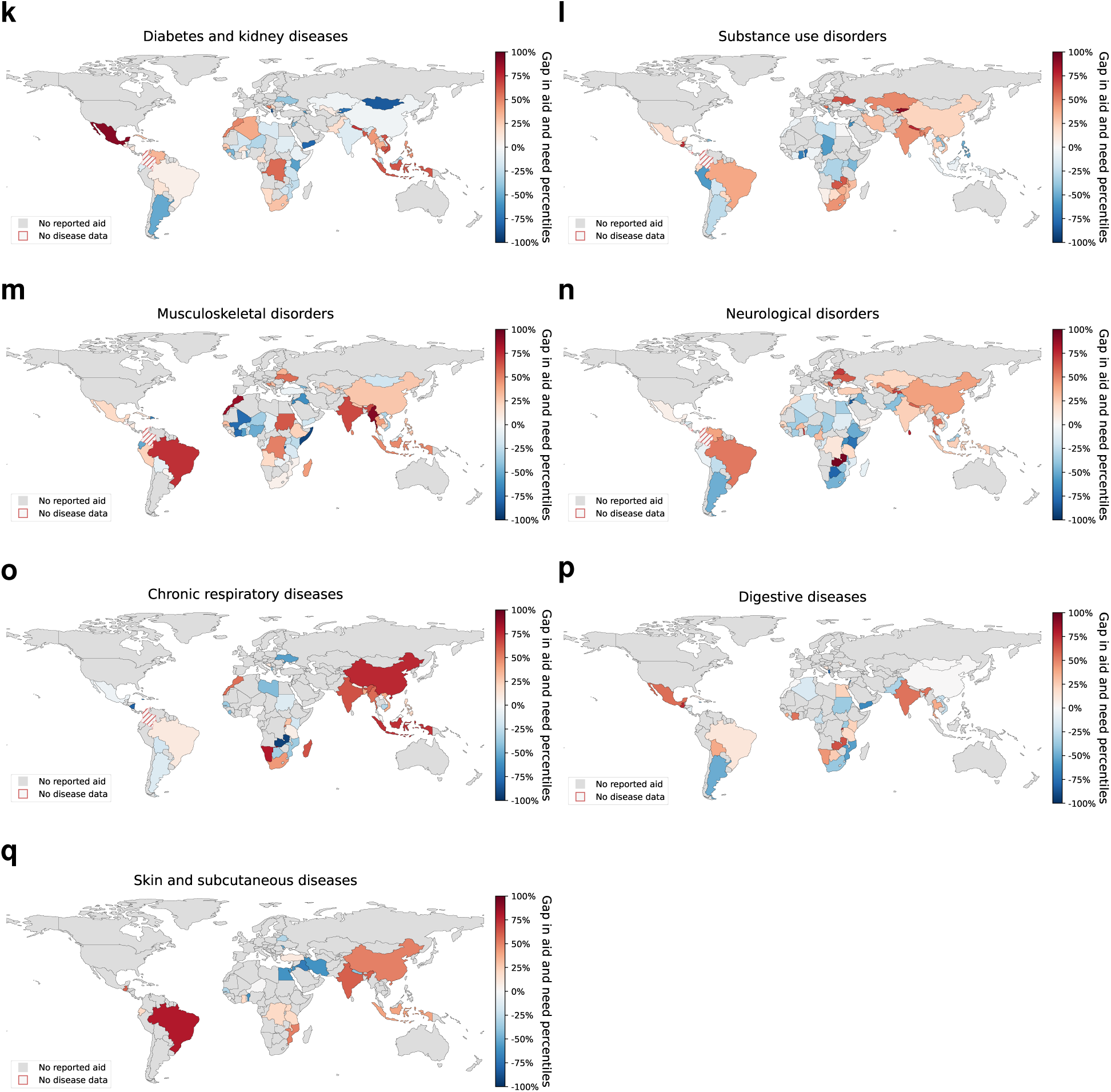
Geographic distribution of aid–burden misalignment (for 2020–2021). The maps show the relative aid–burden misalignment by disease. Here, we refer to “aid–burden misalignment” as the difference between a country’s percentile in disease burden (DALYs per 100,000 population) and its percentile in per capita aid, calculated within income groups. The figure is analogous to Figure 9 in the main paper, except for the different time frame.

**Fig. S7.**
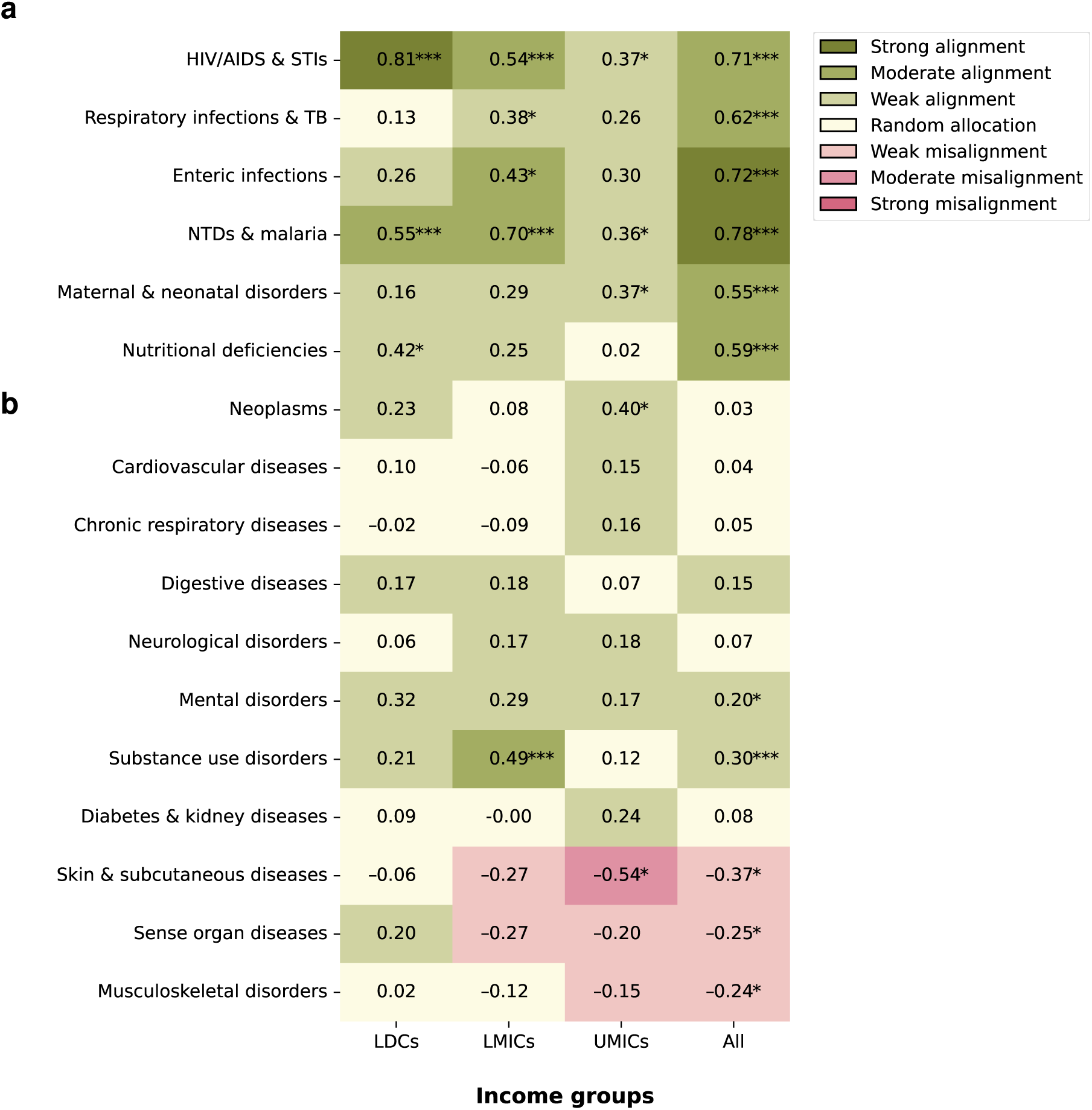
Correlation between disease-specific aid and burden across income groups (as a sensitivity analysis discounting for the surge in aid earmarked to COVID-19). Shown are Spearman’s rank correlation coefficients between per capita aid (accumulated over the time period 2000–2021) and disease burden across country income groups: least developed countries (LDCs), lower-middle-income countries (LMICs), upper-middle-income countries (UMICs), and all countries combined. However, the 2020 and 2021 values for respiratory infections and TB are replaced with the values for 2019 to assess the sensitivity of the correlation to effects from the COVID-19 pandemic. All other values are unchanged. **a**, The top six rows report CMNNDs. **b**, The bottom 11 rows show NCDs. Green indicates positive and red negative correlations. Asterisks denote statistically significant correlations based on a two-sided Spearman rank test (^∗^ *p <* 0.05; ^∗∗^ *p <* 0.01; ^∗∗∗^ *p <* 0.001). Legend interpretation: −1 to −0.7: strong misalignment of health aid with disease burden; −0.7 to −0.3: moderate misalignment; −0.3 to −0.1: weak misalignment, −0.1 to 0.1: indicative of random allocation; 0.1 to 0.3: weak alignment, 0.3 to 0.7, moderate alignment; and 0.7 to 1.0 strong alignment. Abbreviations: STIs: sexually transmitted infections; TB, tuberculosis; NTDs: neglected tropical diseases.

**Fig. S8.**
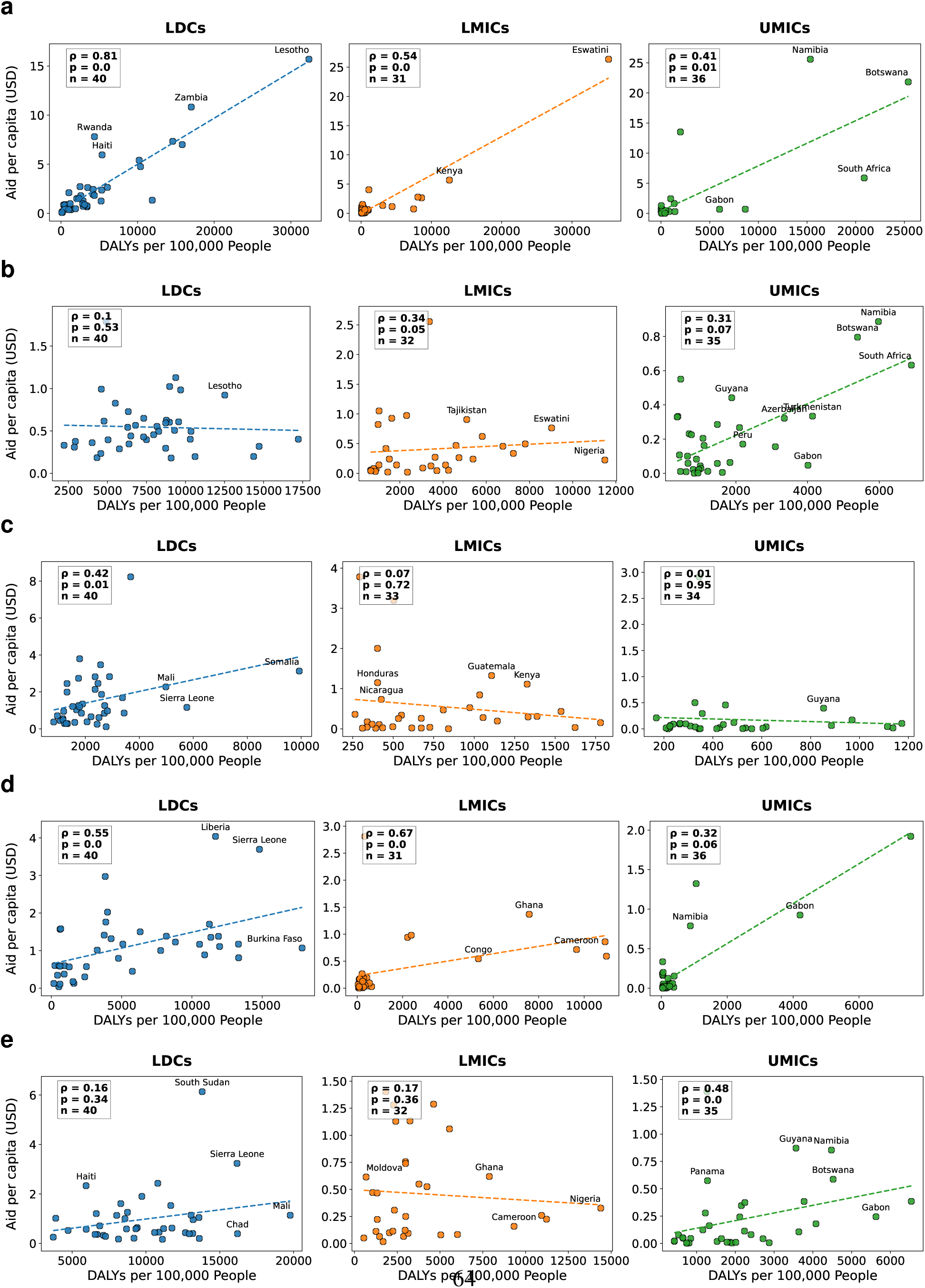
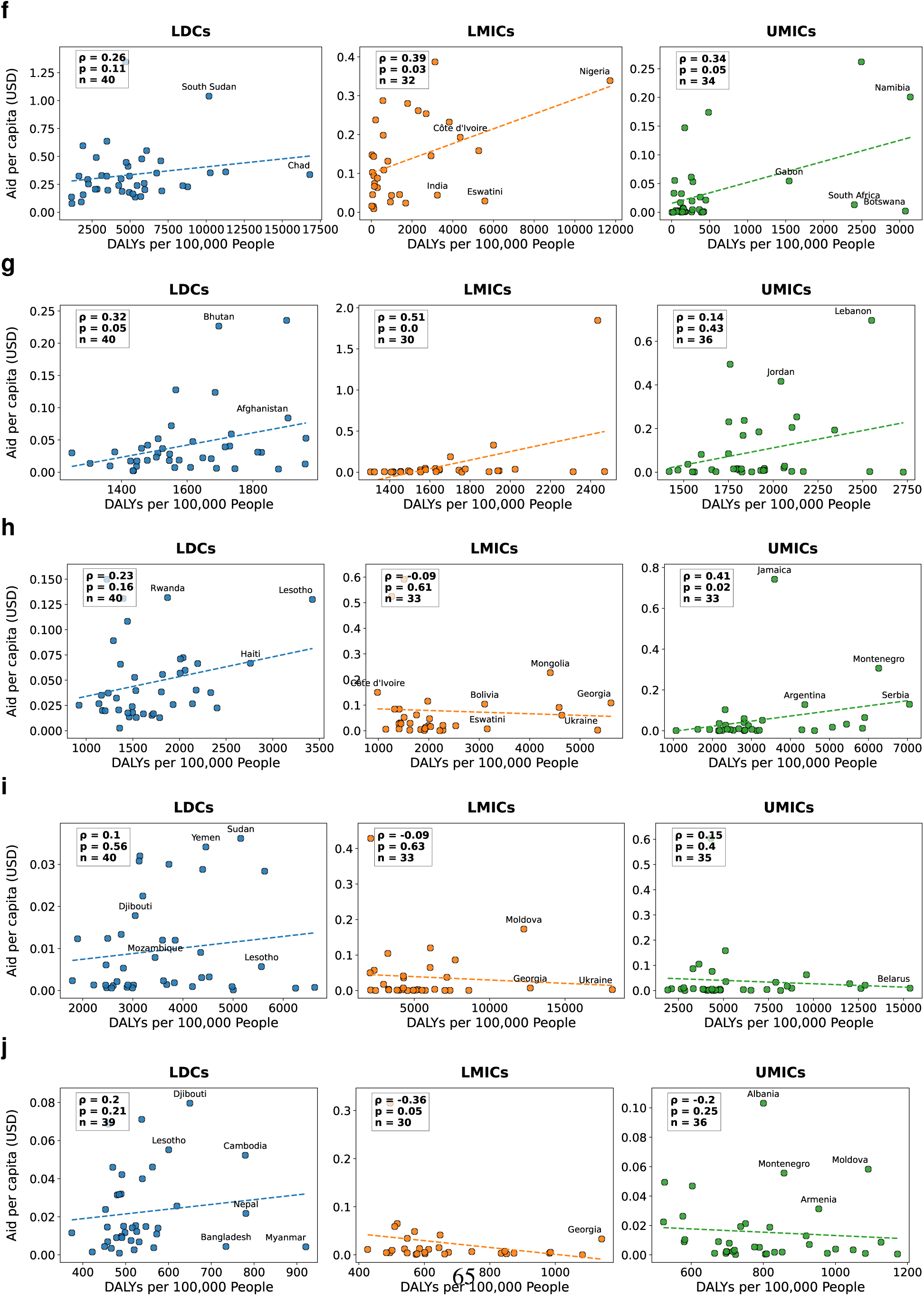
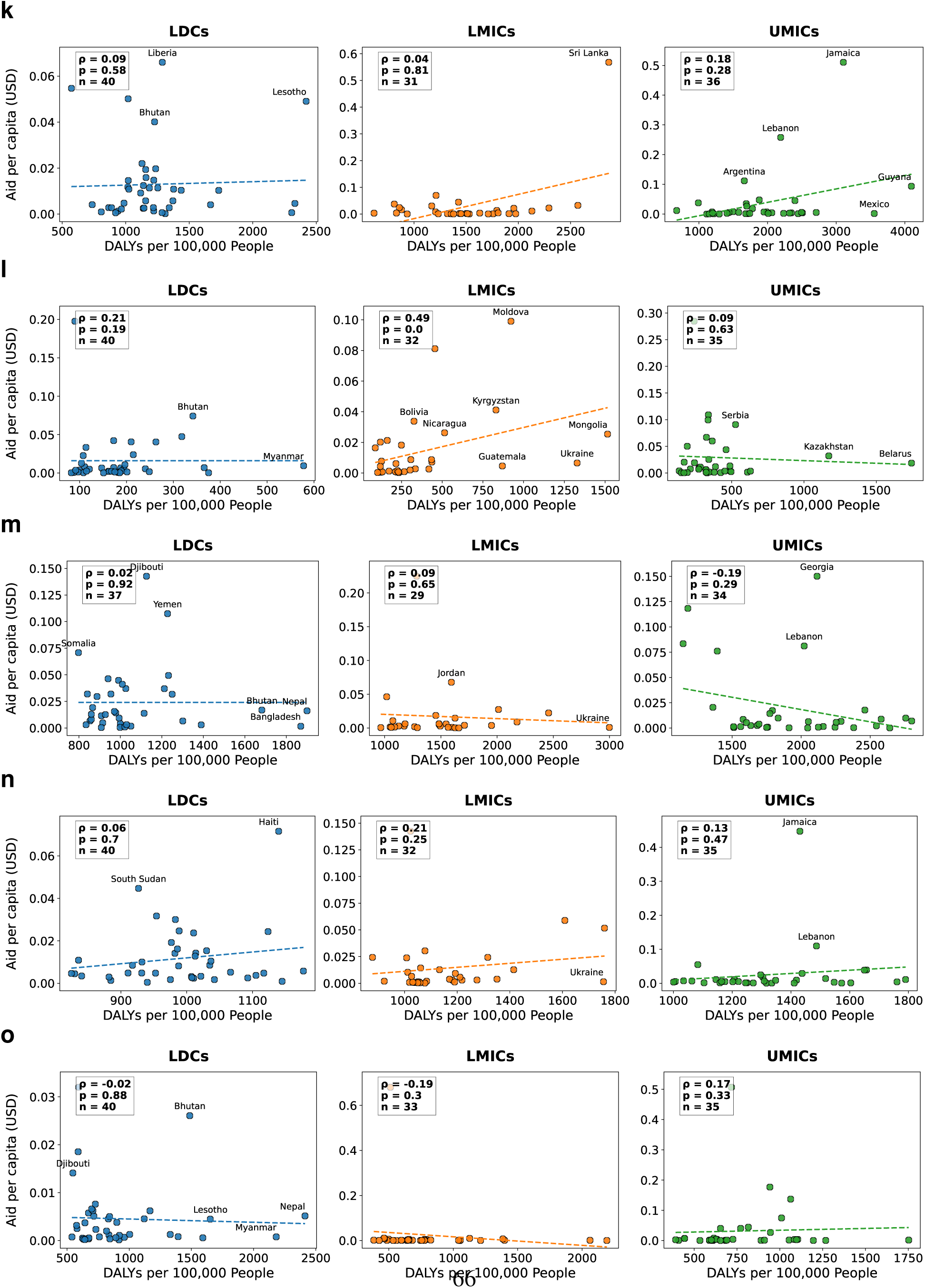
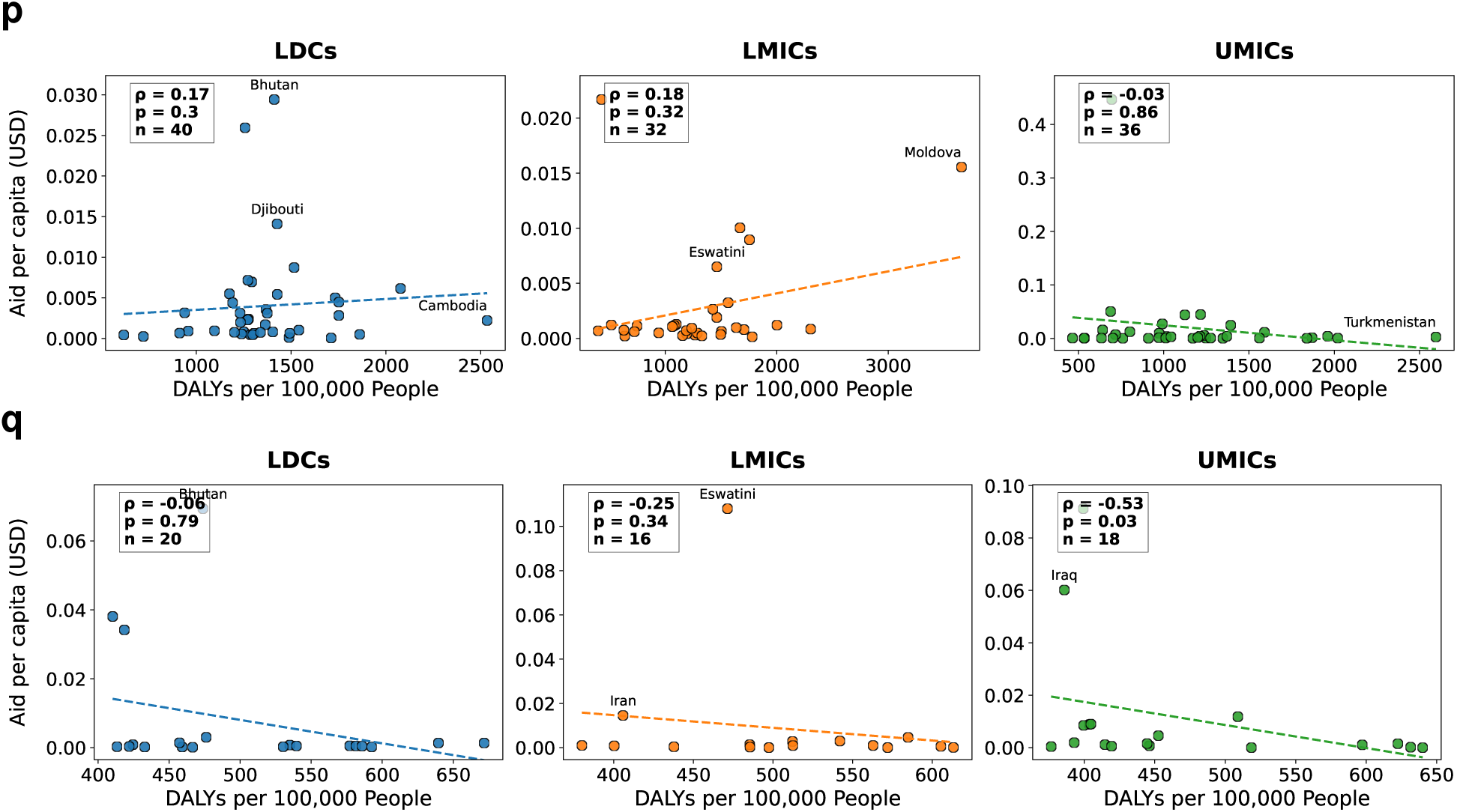
Funding disparities at the country level (without the years 2020 and 2021 for respiratory infections and TB). Otherwise analogous to Figure 8 in the main paper. The figure shows: **a**, HIV-AIDS and sexually transmitted infections; **b**, respiratory infections and tuberculosis; **c**, nutritional deficiencies; **d**, neglected tropical diseases and malaria; **e**, maternal and neonatal disorders; **f**, enteric infections; **g**, mental disorders; **h**, neoplasms; **i**, cardiovascular diseases; **j**, sense organ diseases; **k**, diabetes and kidney diseases; **l**, substance use disorders; **m**, musculoskeletal disorders; **n**, neurological disorders; **o**, chronic respiratory diseases; **p**, digestive diseases; and **q**, skin and subcutaneous diseases.

**Fig. S9.**
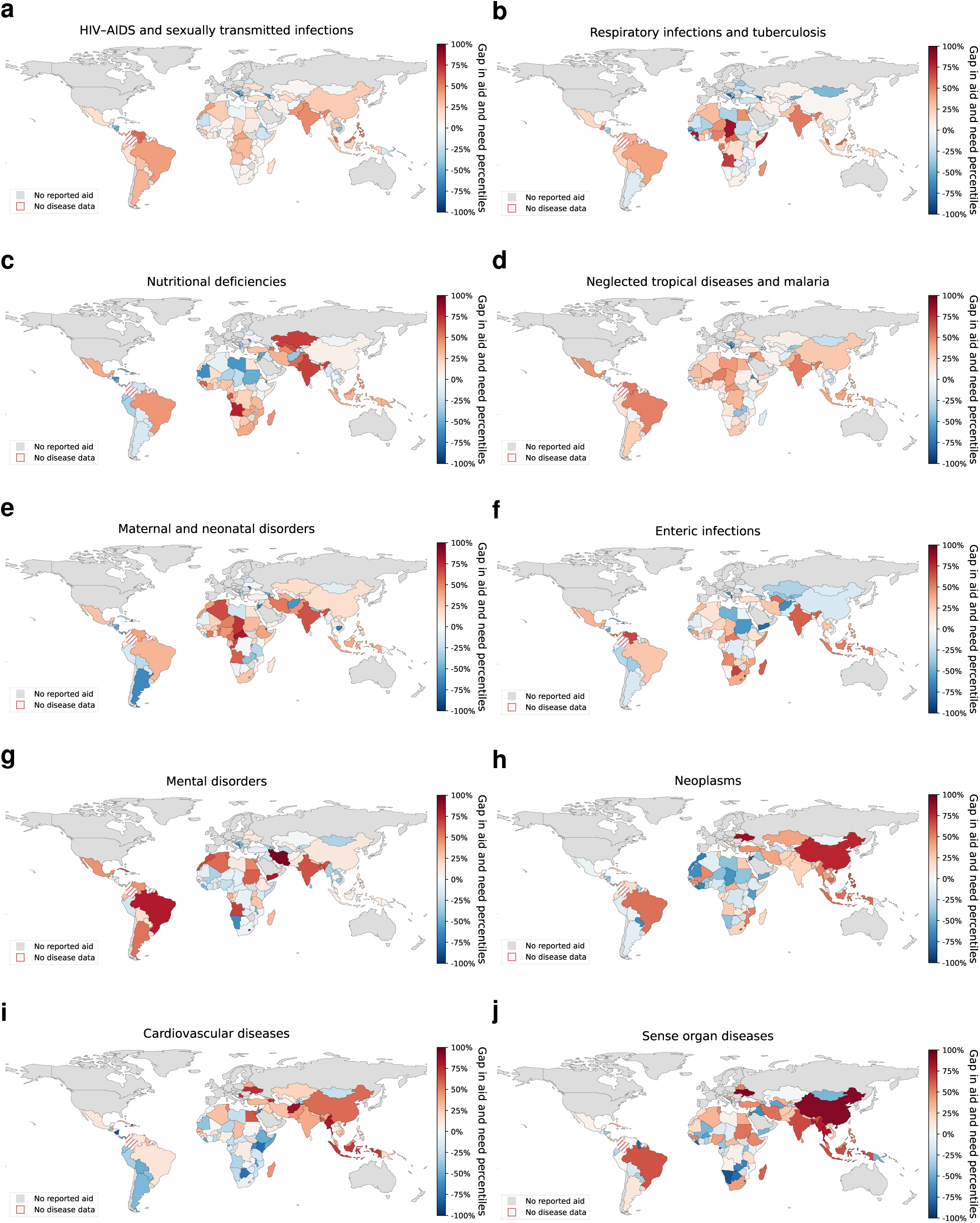
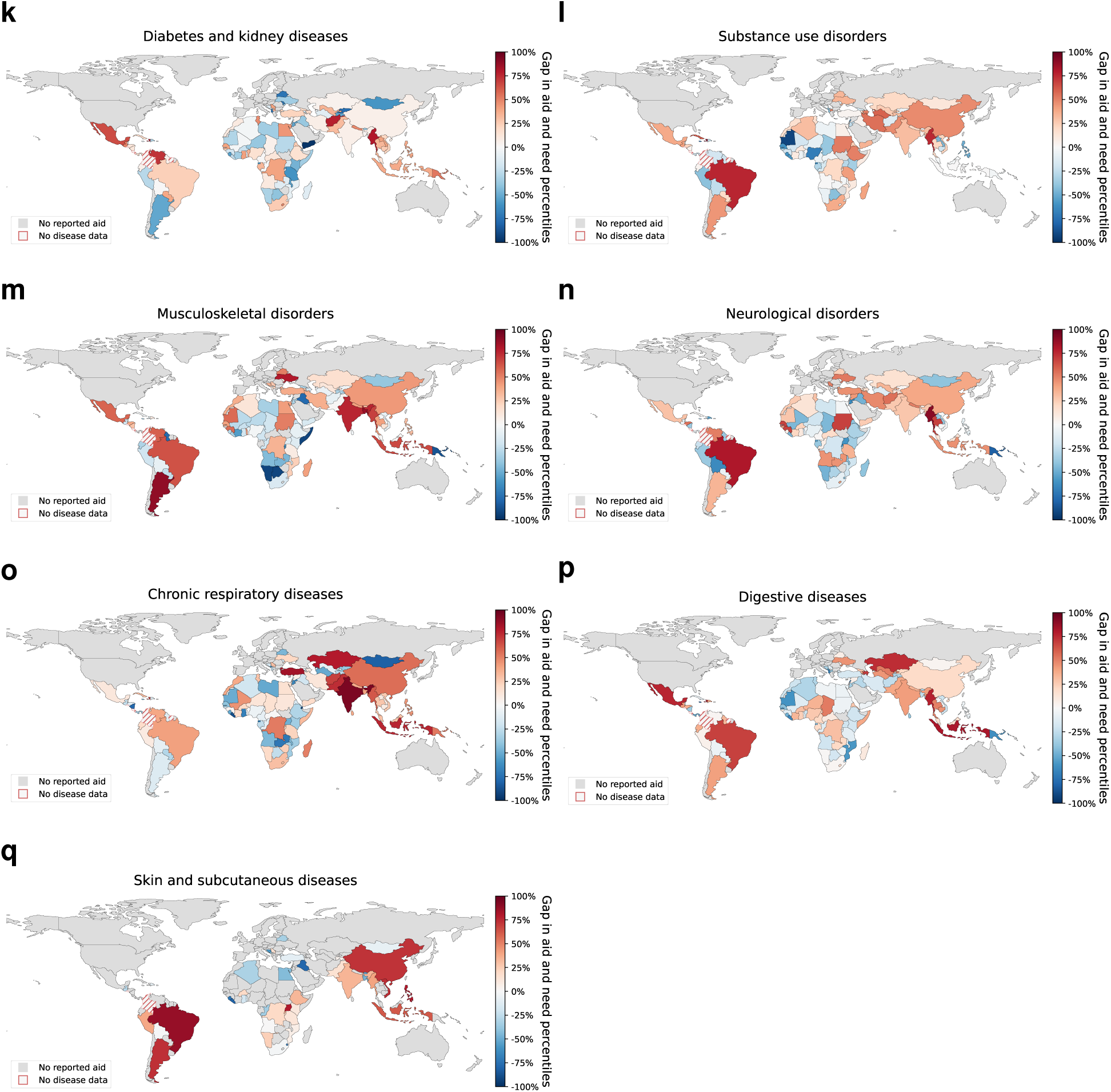
Geographic distribution of aid–burden misalignment without the years 2020 and 2021 for respiratory infections and TB. The maps show the relative aid–burden misalignment by disease. Here, we refer to “aid–burden misalignment” as the difference between a country’s percentile in disease burden (DALYs per 100,000 population) and its percentile in per capita aid, calculated within income groups. The figure is analogous to Figure 9 in the main paper, except for the different time frame.

**Fig. S10.**
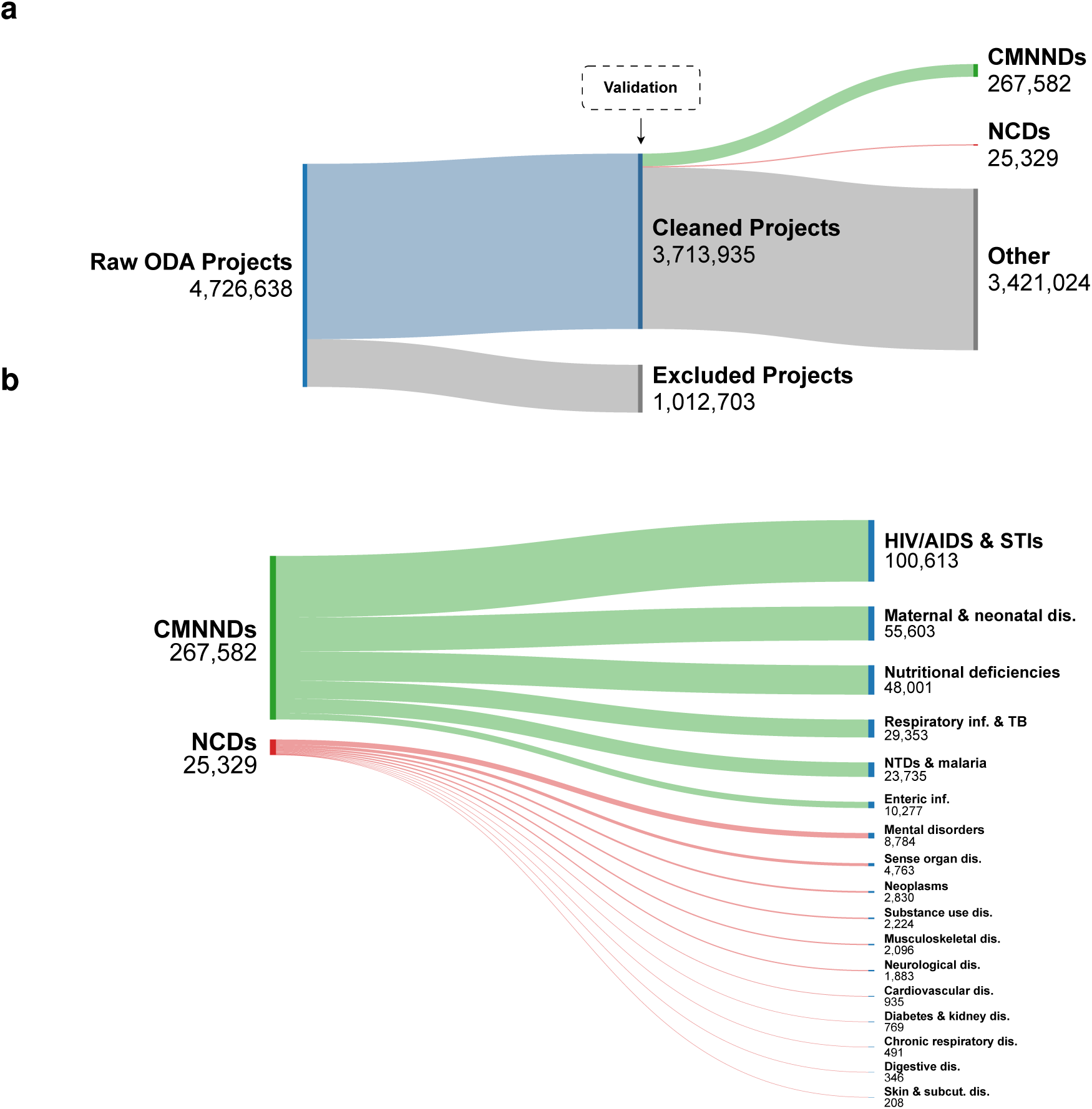
ODA project preprocessing and classification flow. **a**, Flow of ODA projects from the raw corpus through preprocessing (i.e., removing invalid entries) to disease classification. Validation occurred after cleaning and before disease classification. **b**, Distribution of projects across the 17 disease categories. Ribbon width is proportional to the number of projects.

## Supplementary Materials

### S1 Comparison against validation datasets

To assess the accuracy and reliability of our ML approach, we employed a two-edged validation strategy. First, we benchmarked our approach against a conventional keyword-based classification method (inspired by, e.g., [39]), which is interpretable, but unable to capture complex semantics as effectively as our LLM approach. Second, we manually annotated 2,060 aid projects and compared the labels to the LLM-generated classifications; however, this approach lacks the scalability of our machine learning pipeline.

#### S1.1 Comparison against keyword-based approach

##### Keyword list

To identify disease-related projects, we developed comprehensive keyword lists for each disease category. For this, we drew upon multiple authoritative sources, including sub-categories from the Global Burden of Disease study, the PubMed Medical Subject Headings (MeSH) [99], and well-known disease synonyms. We selected keywords carefully to ensure they are specific to each disease category, so that we avoid overly broad terms that might include unrelated projects (i.e., to reduce false positives). For example, we considered the term “TB” for tuberculosis, but we excluded it because it often appears in unrelated contexts. To ensure the accuracy and relevance of our search terms, we consulted a panel of three experts (two public health researchers and one medical professional) to review and approve the final keyword lists.

To illustrate our approach, we elaborate our approach for the keywords that we selected for the category nutritional deficiencies. The keywords for this disease category include terms such as `malnutrition`, `undernutrition`, `stunting`, and specific deficiencies such as `iodine deficiency` and `zinc deficiency`. We derived these keywords from Global Burden of Disease (GBD) study subcategories, to ensure our search terms align closely with the study’s health categories. For instance, the GBD subcategories *“A.7.3 Vitamin A deficiency”* and *“A.7.4 Dietary iron deficiency”* inform our inclusion of the terms `vitamin A deficiency` and `iron deficiency`.

We provide the full list of keywords in Supplementary Material S5 and in our GitHub repository. For illustration purposes, we list the keywords for HIV/AIDS and STIs, including a brief justification, in Supplementary Table S3).

**Table S3:** Keywords used as search terms for HIV/AIDS and sexually transmitted infections.

| Keyword | Reasoning |
| --- | --- |
| sexually transmitted infection | Direct descriptor of the subcategory |
| STI / STIs | Acronym for sexually transmitted infection |
| STD / STDs | Acronym for sexually transmitted disease |
| sexually transmitted disease | Alternative term for sexually transmitted infection |
| safe sex | Practice to prevent transmission of STIs |
| HIV | Acronym for human immunodeficiency virus, the cause of AIDS |
| AIDS | Acronym for acquired immunodeficiency syndrome, which is caused by HIV |
| acquired immunodeficiency syndrome | Full name of AIDS |
| antiretroviral | Type of medication used to treat HIV/AIDS |
| human immunodeficiency virus | Full name of HIV |
| syphilis | Name of the specific sexually transmitted infection |
| treponema pallidum | Scientific name of the bacterium causing syphilis |
| chancre | Primary symptom of syphilis infection |
| chlamydial infection | Name of the specific sexually transmitted infection |
| chlamydia | Common name for chlamydial infection |
| gonococcal infection | Name of the specific sexually transmitted infection |
| gonorrhea | Common name for gonococcal infection |
| trichomoniasis | Name of the specific sexually transmitted infection |
| trichomonas vaginalis | Scientific name of the parasite causing trichomoniasis |
| genital herpes | Name of the specific sexually transmitted infection |
| genital ulcer | Common symptom of genital herpes |

##### Classification

We then searched for exact matches (case-insensitive) within the title, long description, and short description of each project. Again, aid projects were assigned to one or more categories based on the matches. Projects without keyword match were classified as “Other”.

##### Results

The comparison between the LLM-based and keyword-based classifications reveals substantial agreement across disease categories, as evidenced by a strong diagonal pattern in our comparative analysis (Supplementary Figure S11). This alignment is particularly pronounced for well-defined disease categories such as HIV/AIDS and STIs and Neglected tropical diseases. However, we observe some divergence between methods for closely related conditions, such as chronic respiratory diseases and respiratory infections and TB, which we attribute to that the keyword-based search may not fully capture the semantics of the underlying aid description, which we confirm empirically in the next section using our manually-annotated dataset.

**Fig. S11.**
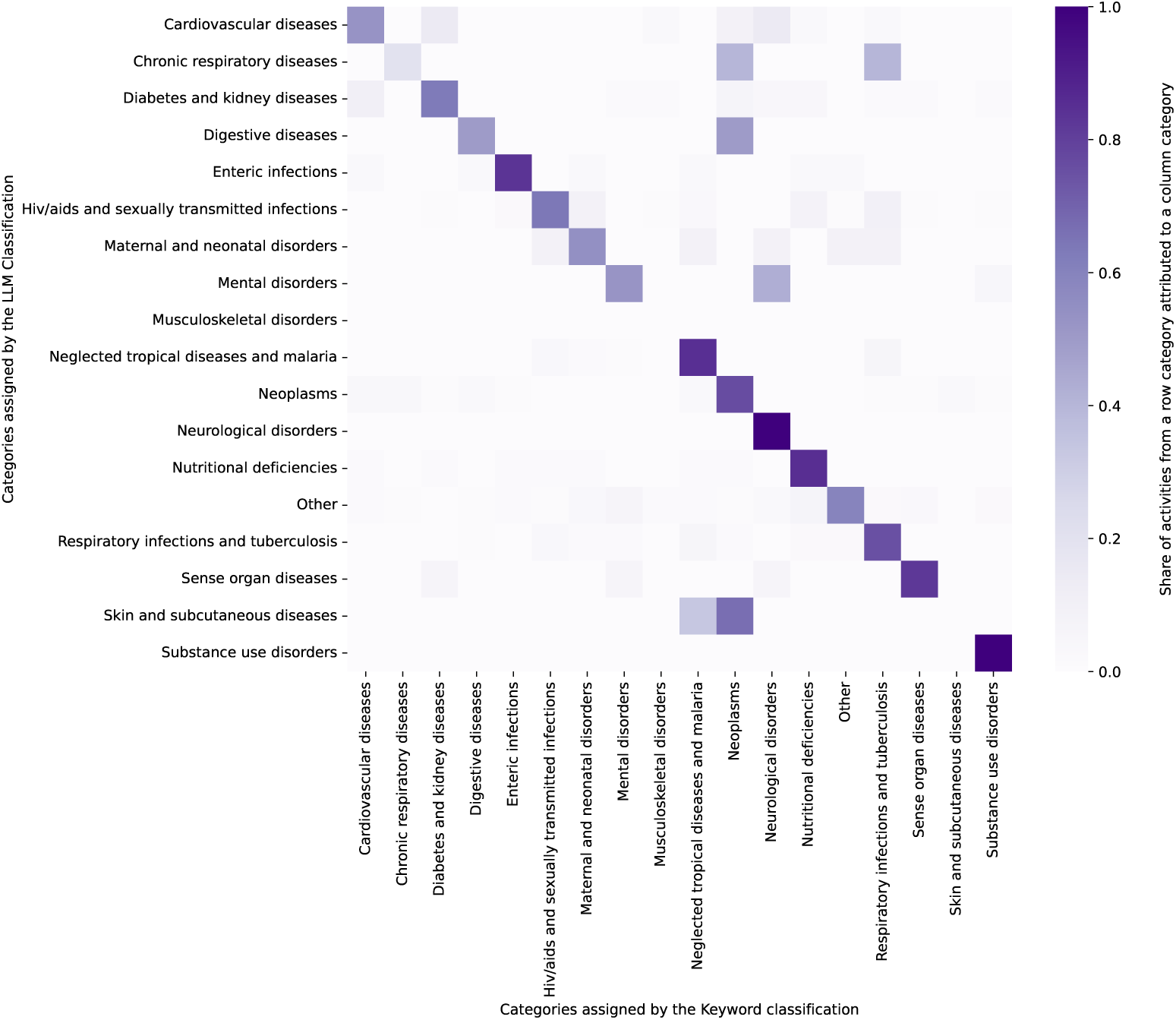
Comparison between our machine learning pipeline based on LLMs and keyword-based classification. The comparison shows how aid projects (“activities”) are classified into disease categories based on the machine learning approach (y-axis) and based on the keyword-based approach (x-axis). The cells show the percentage of aid projects from a specific disease category (as per machine learning pipeline) that are classified into a specific disease category (as per the keyword-based approach). Hence, rows should add up to 1.0 (=100%). An exception is musculoskeletal diseases, where we struggled to identify aid projects in the OECD CRS data using keywords. Upon manual inspection, this is often because simple keywords are not very meaningful in this context; this is confirmed in our comparison of the keyword-based approach against the manually-annotated validation dataset (see Supplementary Table S7). Therein, we find: On the one hand, the problems are due to the fact that the keyword-based approach struggles with classifying projects earmarked to musculoskeletal diseases (and not due to problems of our machine learning pipeline). As a case in point, the keyword-based approach has both a precision and recall of 0.00. On the other hand, this again highlights the benefits of our machine learning pipeline. Abbreviations: LLM, large language model.

#### S1.2 Comparison against manually annotated dataset

##### Validation dataset

We manually classified a subset of 2,060 aid projects to compare the labels against our machine learning pipeline (and against the keyword-based approach). The projects were randomly sampled from the study corpus. This ensured that the validation set reflected the corpus-level distribution of disease areas and funding priorities; we additionally verified ex post that all disease categories were represented in the sample (Supplementary Table S4). The sample size was chosen to balance the feasibility of manual annotation with the need for statistically precise performance estimates. A sensitivity analysis demonstrates that the size of the validation dataset is sufficient to obtain robust estimates of classification performance (Supplementary Figure S12).

To ensure precise annotations, we excluded water, sanitation, and hygiene (WASH) projects unless they explicitly focus on enteric conditions such as cholera. As expected, there is a subset of projects that are not relevant to specific disease categories (e.g., as they focus on other sustainability targets such as climate action) and are thus labeled as “Others”. We also see frequencies that reflect the overall funding priorities (e.g., there are generally more labeled samples for CMNNDs than for NCDs).

Manual annotation was conducted by a primary annotator with domain knowledge in global health metrics and development aid projects, and independently classified by a second annotator. We report inter-annotator agreement using Cohen’s kappa (*κ*) and label consistency rate (LCR), i.e., the proportion of items receiving identical labels across annotators. To obtain a 95% confidence interval for *κ*, we applied a non-parametric bootstrap: we resampled the paired annotation labels with replacement 1,000 times, recomputed *κ* for each bootstrap sample, and took the central 95% of the resulting distribution as the bounds of the interval. For the LCR, we treated the number of matching labels as a binomial count and derived 95% confidence intervals using the Wilson score method, which yields well-calibrated intervals based on the observed proportion and the total number of annotated items. We observe substantial agreement across annotators (*κ* = 0.91, 95% CI: [0.90, 0.93]; LCR = 0.94, 95% CI: [0.93, 0.95]), which strengthens confidence in the reliability of the manually-annotated validation dataset. A category-specific confusion matrix is shown in Supplementary Figure S13.

**Fig. S12.**
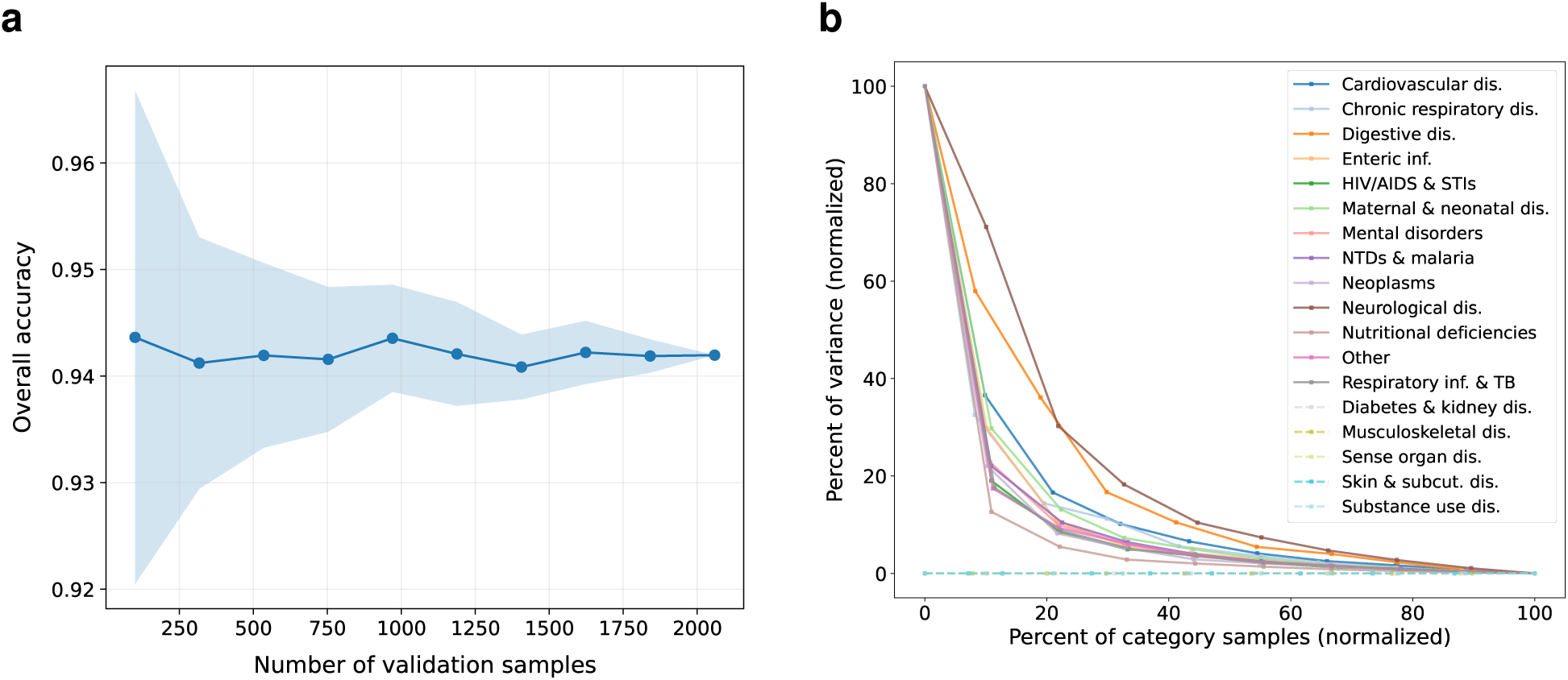
Sensitivity analysis of classification performance. **a**, Convergence of overall classification performance, showing accuracy as a function of the number of validation samples; the line shows mean accuracy and the shaded band the 95% bootstrap confidence interval. Accuracy increases rapidly and plateaus at approximately 0.94 once several hundred samples are included, indicating that the overall performance estimate has effectively converged and that the size of the validation set (*n* = 2,060) is sufficient. **b**, Sensitivity analysis of category-level performance, showing the variance of per-category accuracies versus the number of labeled projects per category. Both axes are min–max normalized to 0–100% within each category to enable comparison across categories with different sample sizes and variance ranges. The variance decreases sharply and stabilizes at a low level as per-category sample sizes approach those in the final validation set.

**Fig. S13.**
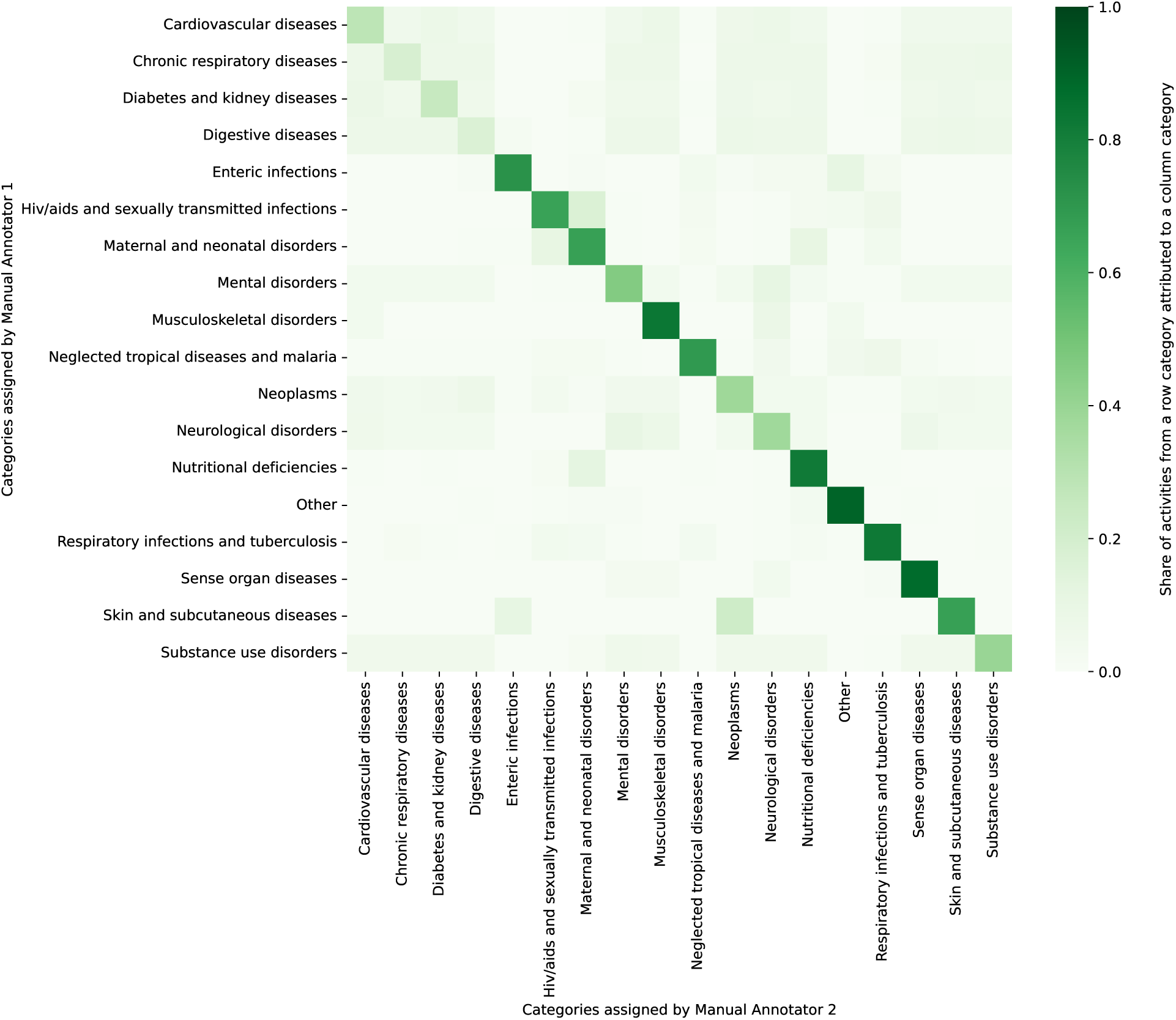
Comparison between manual annotators. The comparison shows how aid projects (“activities”) are classified into disease categories by Manual Annotator 1 (rows) and Manual Annotator 2 (columns). Each cell reports the share of activities from a row category that is assigned to the corresponding column category, so rows sum to 1. Values on the diagonal indicate agreement between annotators for a given disease category, while off-diagonal cells indicate disagreement.

**Table S4:** Frequency of labels in the manually-annotated validation dataset.

| Label | Frequency |
| --- | --- |
| Respiratory infections and tuberculosis | 182 |
| Neglected tropical diseases and malaria | 144 |
| Nutritional deficiencies | 142 |
| Mental disorders | 103 |
| HIV/AIDS and sexually transmitted infections | 100 |
| Neoplasms | 78 |
| Maternal and neonatal disorders | 75 |
| Enteric infections | 63 |
| Sense organ diseases | 58 |
| Cardiovascular diseases | 50 |
| Neurological disorders | 50 |
| Substance use disorders | 46 |
| Diabetes and kidney diseases | 38 |
| Chronic respiratory diseases | 24 |
| Musculoskeletal disorders | 22 |
| Digestive diseases | 18 |
| Skin and subcutaneous diseases | 6 |
| Other | 987 |

##### Performance metrics (overall)

The validation of our machine learning approach against the manually-labeled dataset demonstrates the strong performance of our pipeline (Supplementary Table S5). In particular, our machine learning pipeline has a superior performance compared to the keyword-based approach: overall accuracy (0.93 vs 0.71), precision (0.95 vs 0.73), recall (0.94 vs 0.71), and *F*_1_-score (0.94 vs 0.72). The superior performance extends to both micro- and macro-averaged performance metrics, indicating a reliable performance across both common and rare disease categories.

##### Performance metrics (stratified by disease categories)

Further, we performed a stratified analysis of the performance by different disease categories for both our machine learning approach (Supplementary Table S6) and the keyword-based approach (Supplementary Table S7). The analysis confirms that the LLM method maintains high recall (*>* 0.95) across the majority of disease categories, including chronic respiratory diseases, diabetes and kidney diseases, sense organ diseases, and skin and subcutaneous diseases. In contrast, the keyword-based approach shows clear limitations, including large failure rates (e.g., a precision and recall of both 0.00) in classifying musculoskeletal disorders and skin conditions, and notably low recall for chronic respiratory diseases (0.04), digestive diseases (0.06), and maternal and neonatal disorders (0.09).

**Table S5:** Overall classification performance based on manually-labeled validation data. Abbreviations: ML, machine learning.

| Performance metric | Our ML approach | Keyword-based approach |
| --- | --- | --- |
| Accuracy | 0.93 | 0.71 |
| Micro-averaged precision | 0.95 | 0.73 |
| Micro-averaged recall | 0.94 | 0.71 |
| Micro-averaged $F_1$-score | 0.94 | 0.72 |
| Macro-averaged precision | 0.91 | 0.81 |
| Macro-averaged recall | 0.96 | 0.44 |
| Macro-averaged $F_1$-score | 0.93 | 0.50 |

**Table S6:**
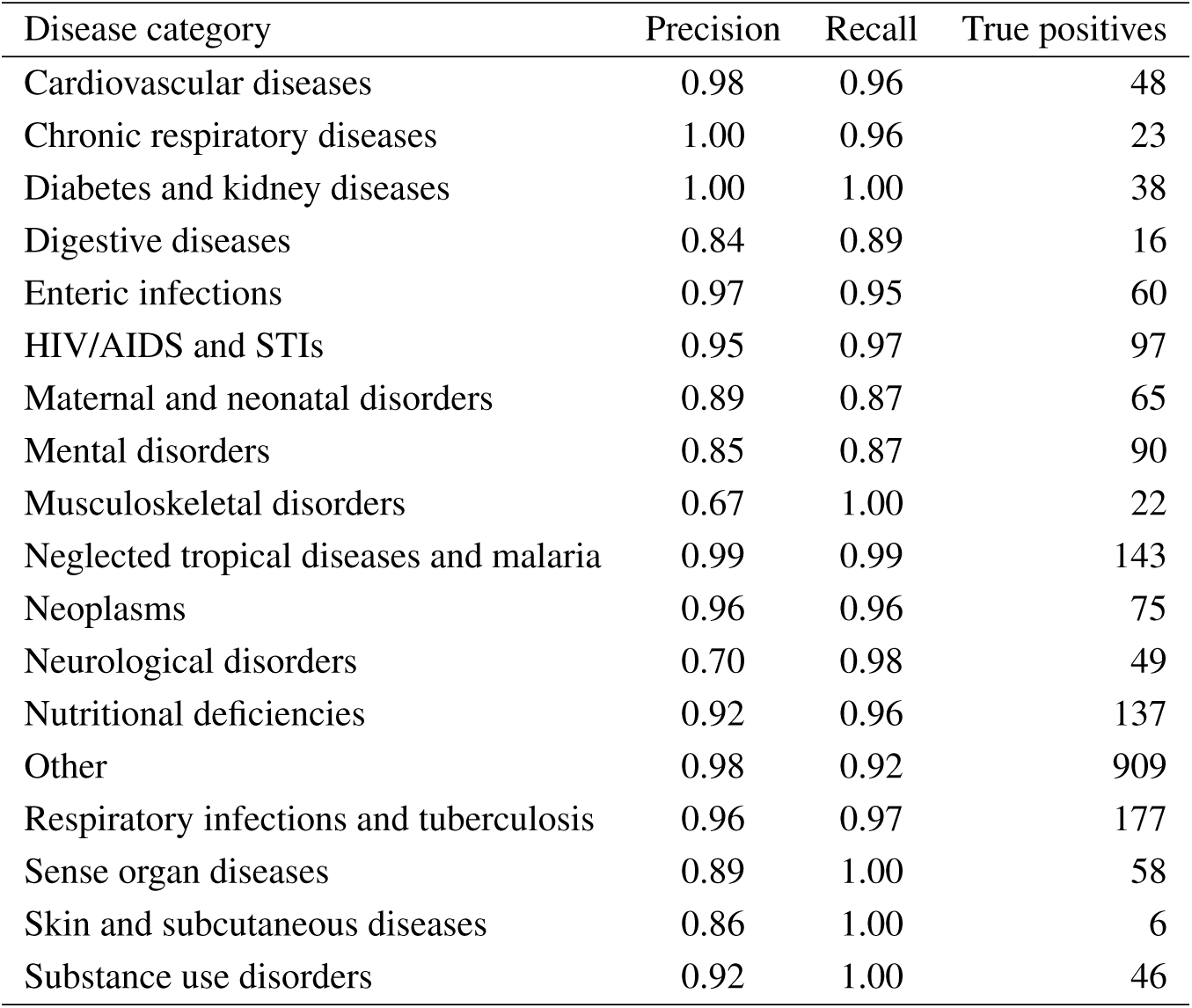
Classification performance by disease categories for the machine learning approach. Results are compared against the manually-annotated validation dataset.

| Disease category | Precision | Recall | True positives |
| --- | --- | --- | --- |
| Cardiovascular diseases | 0.98 | 0.96 | 48 |
| Chronic respiratory diseases | 1.00 | 0.96 | 23 |
| Diabetes and kidney diseases | 1.00 | 1.00 | 38 |
| Digestive diseases | 0.84 | 0.89 | 16 |
| Enteric infections | 0.97 | 0.95 | 60 |
| HIV/AIDS and STIs | 0.95 | 0.97 | 97 |
| Maternal and neonatal disorders | 0.89 | 0.87 | 65 |
| Mental disorders | 0.85 | 0.87 | 90 |
| Musculoskeletal disorders | 0.67 | 1.00 | 22 |
| Neglected tropical diseases and malaria | 0.99 | 0.99 | 143 |
| Neoplasms | 0.96 | 0.96 | 75 |
| Neurological disorders | 0.70 | 0.98 | 49 |
| Nutritional deficiencies | 0.92 | 0.96 | 137 |
| Other | 0.98 | 0.92 | 909 |
| Respiratory infections and tuberculosis | 0.96 | 0.97 | 177 |
| Sense organ diseases | 0.89 | 1.00 | 58 |
| Skin and subcutaneous diseases | 0.86 | 1.00 | 6 |
| Substance use disorders | 0.92 | 1.00 | 46 |

**Table S7:** Classification performance by disease categories for the keyword-based approach. Results are compared against the manually-annotated validation dataset.

| Disease category | Precision | Recall | True positives |
| --- | --- | --- | --- |
| Cardiovascular diseases | 0.74 | 0.40 | 20 |
| Chronic respiratory diseases | 0.20 | 0.04 | 1 |
| Diabetes and kidney diseases | 0.81 | 0.76 | 29 |
| Digestive diseases | 0.50 | 0.06 | 1 |
| Enteric infections | 0.91 | 0.48 | 30 |
| HIV/AIDS and STIs | 0.75 | 0.77 | 77 |
| Maternal and neonatal disorders | 0.88 | 0.09 | 7 |
| Mental disorders | 0.95 | 0.18 | 19 |
| Musculoskeletal disorders | 0.00 | 0.00 | 0 |
| Neglected tropical diseases and malaria | 0.97 | 0.83 | 120 |
| Neoplasms | 0.89 | 0.74 | 58 |
| Neurological disorders | 1.00 | 0.24 | 12 |
| Nutritional deficiencies | 0.91 | 0.29 | 41 |
| Other | 0.67 | 0.99 | 979 |
| Respiratory infections and tuberculosis | 0.86 | 0.81 | 147 |
| Sense organ diseases | 0.93 | 0.24 | 14 |
| Skin and subcutaneous diseases | 0.00 | 0.00 | 0 |
| Substance use disorders | 1.00 | 0.11 | 5 |

##### Comparison (confusion matrix)

We further compare how different aid projects in the manually-annotated dataset are classified in comparison to our machine learning approach (see Supplementary Figure S14 for a row-normalized confusion matrix) shows high precision and low misclassification rates, indicated by strong diagonal dominance with minimal off-diagonal elements. Where misclassification occurs, it primarily appears between clinically related conditions.

**Fig. S14.**
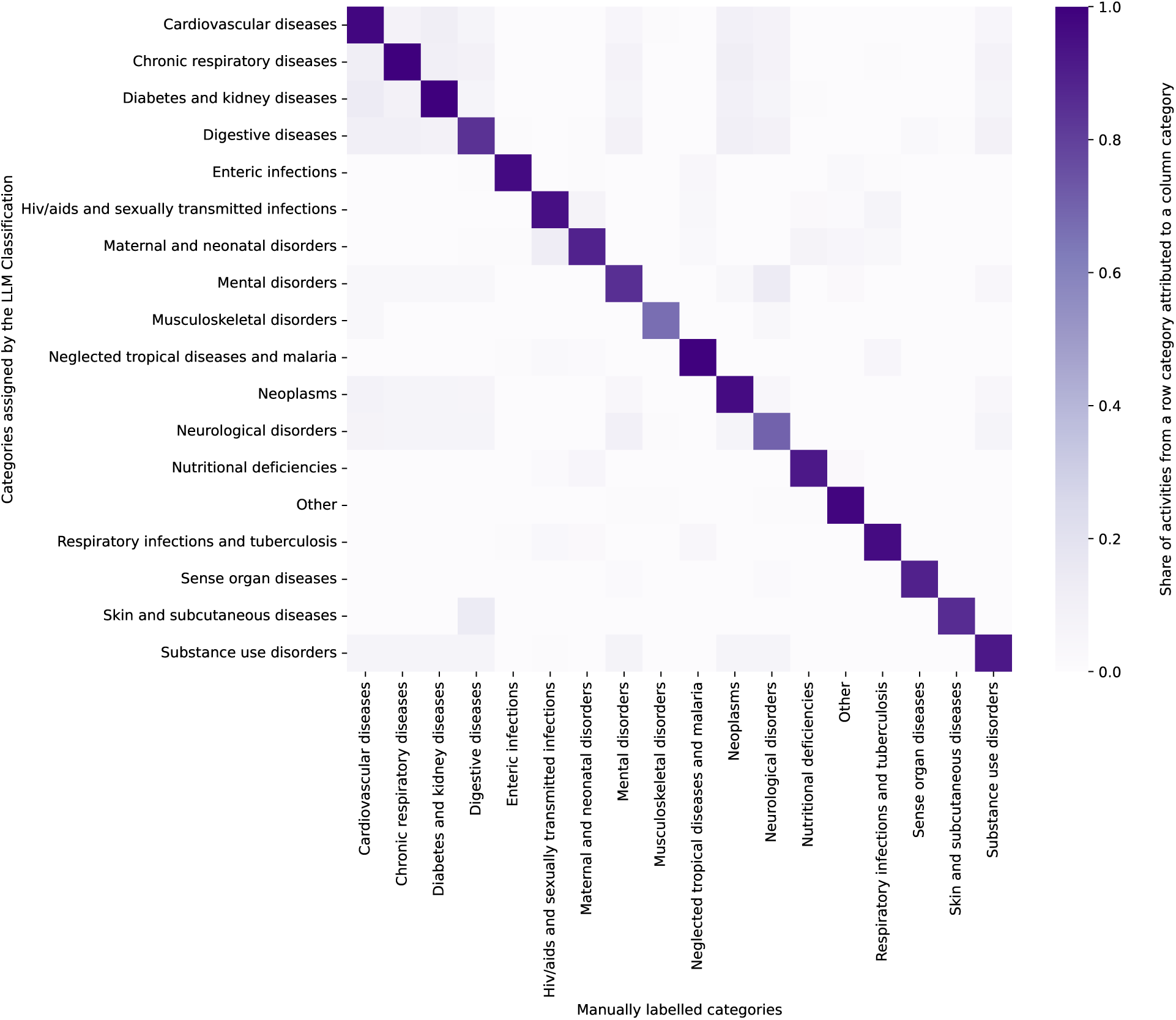
Comparison between our machine learning pipeline and the manually-annotated dataset. The comparison shows how aid projects (“activities”) are classified into disease categories based on our machine learning approach (y-axis) and based on the validation dataset (x-axis). The cells show the percentage of aid projects from a specific disease category (as per machine learning pipeline) that are classified into a specific disease category (as per the validation dataset). Hence, rows should add up to 1.0 (=100%). The diagonal represents the precision of our machine learning pipeline, while the other cells show the percentage of manually-annotated aid projects from one disease category that the LLM assigned to another category. Abbreviations: LLM, large language model.

### S2 Robustness checks

To ensure the robustness of our approach, we quantified the uncertainty and reproducibility of the disease-classification pipeline. For a randomly sampled 1% of projects, we repeated the classification with different random seeds and with an alternative LLM (gpt-4.1-mini-2025-04-14) instead of the main model (Llama-3.1-70B). Agreement across runs and models was summarized using Cohen’s kappa (*κ*) and the label consistency rate (LCR), defined as the proportion of items receiving identical labels across runs. Across random seeds, we observed *κ* = 0.88 (95% CI: [0.87, 0.88]) and an LCR of 0.90 (95% CI: [0.89, 0.90]); across LLMs, *κ* = 0.88 (95% CI: [0.88, 0.88]) and an LCR of 0.90 (95% CI: [0.90, 0.90]). Category-specific confusion matrices (Supplementary Figures S15–S18) show that disagreements are concentrated in a small number of closely related categories, while the vast majority of projects receive stable labels. Together, these analyses indicate high run-to-run reproducibility and substantial inter-model agreement, supporting the robustness of our main results.

**Fig. S15.**
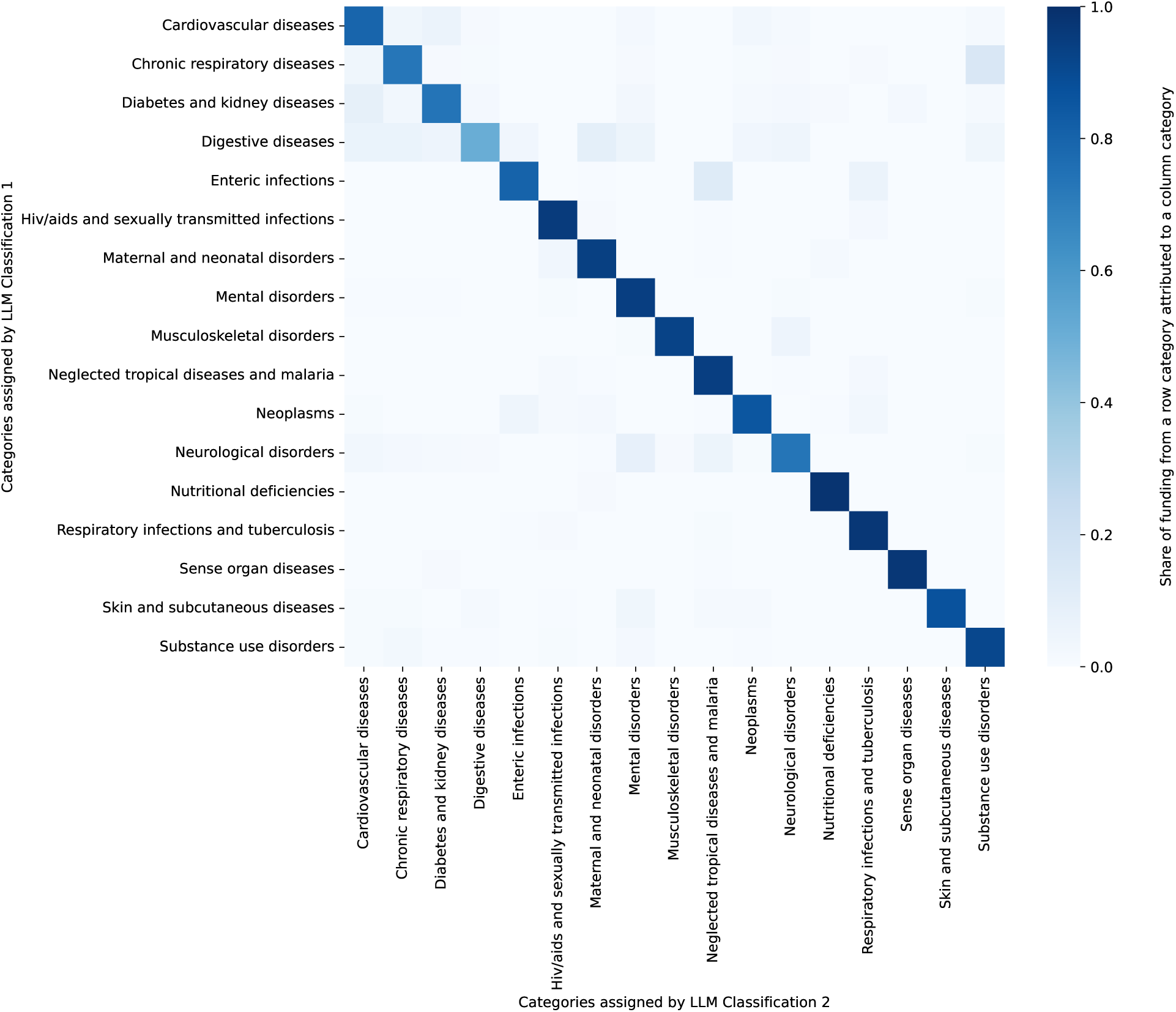
Comparison between classifications from different random seeds, weighted by funding. The comparison shows how aid projects (“activities”) are classified into disease categories by our machine learning pipeline using different random seeds. Each cell reports the share of activities from a row category that is assigned to the corresponding column category, weighted by funding, so rows sum to 1. Values on the diagonal indicate agreement between annotators for a given disease category, while off-diagonal cells indicate disagreement.

**Fig. S16.**
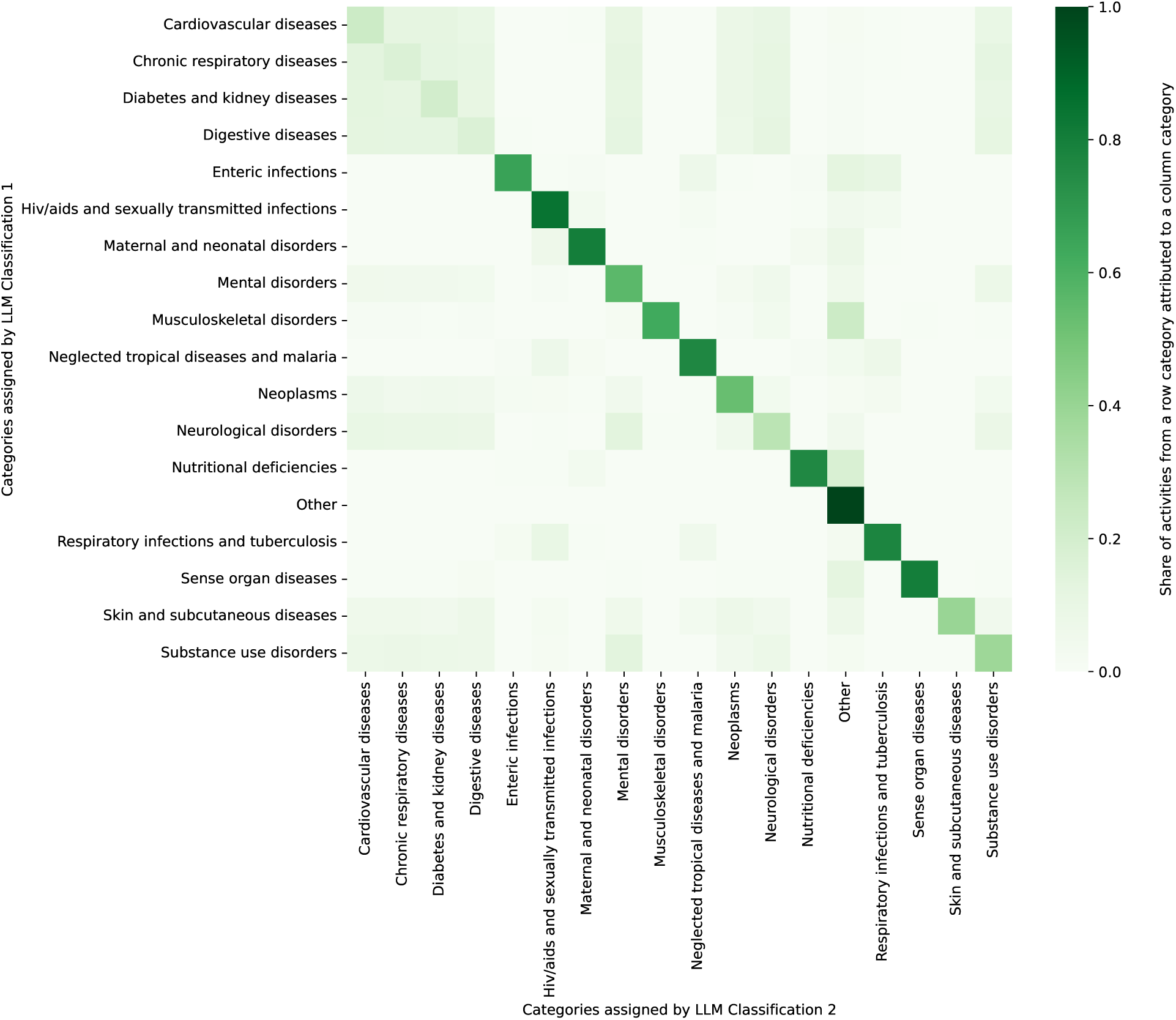
Comparison between classifications from different random seeds. The comparison shows how aid projects (“activities”) are classified into disease categories by our machine learning pipeline using different random seeds. Each cell reports the share of activities from a row category that is assigned to the corresponding column category, so rows sum to 1. Values on the diagonal indicate agreement between annotators for a given disease category, while off-diagonal cells indicate disagreement.

**Fig. S17.**
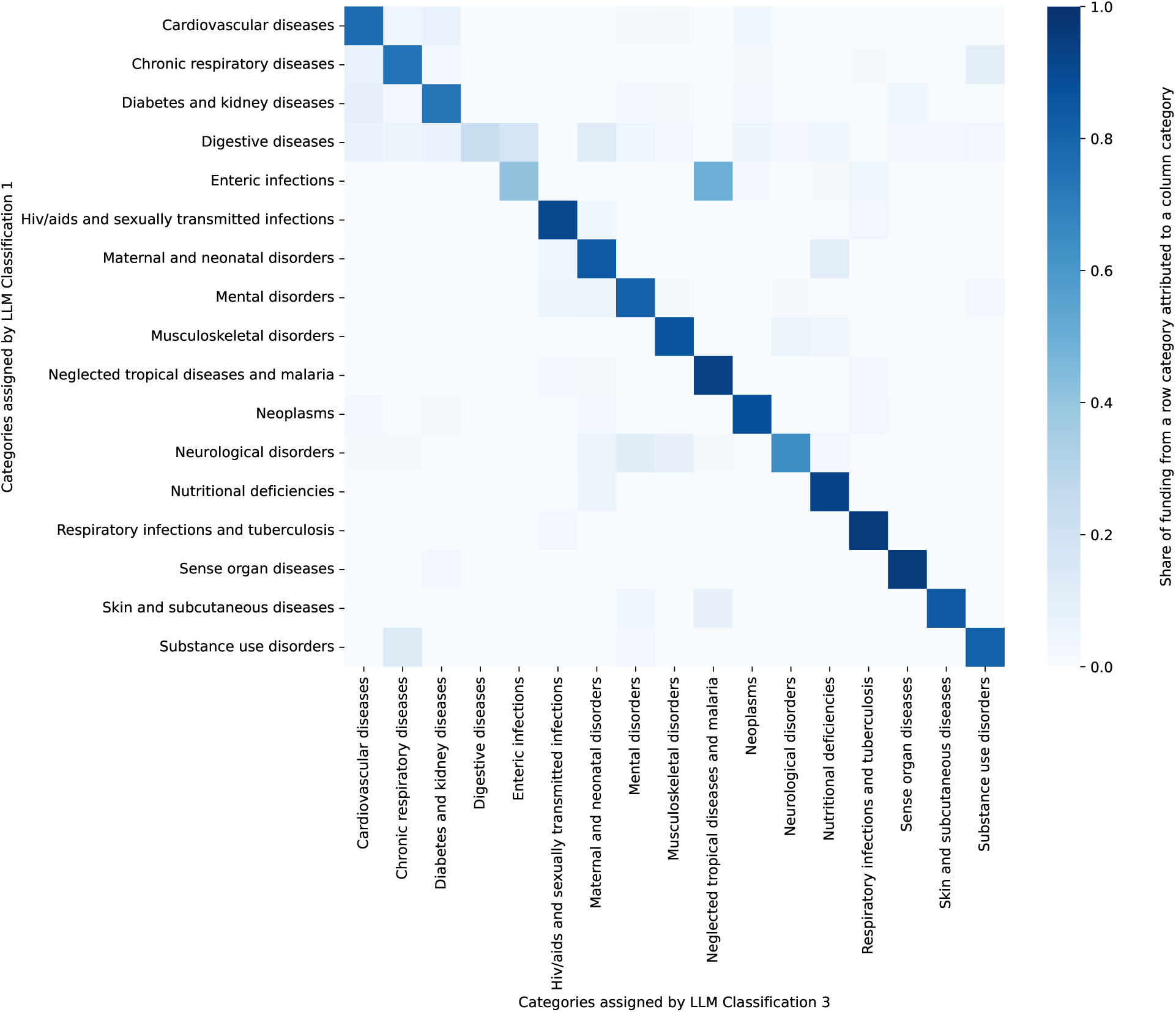
Comparison between classifications from different LLMs, weighted by funding. The comparison shows how aid projects (“activities”) are classified into disease categories by our machine learning pipeline using different LLMs. LLM Classification 1 uses Llama-3.1-70B (rows), LLM Classification 3 uses gpt-4.1-mini-2025-04-14 (columns). Each cell reports the share of activities from a row category that is assigned to the corresponding column category, weighted by funding, so rows sum to 1. Values on the diagonal indicate agreement between annotators for a given disease category, while off-diagonal cells indicate disagreement.

**Fig. S18.**
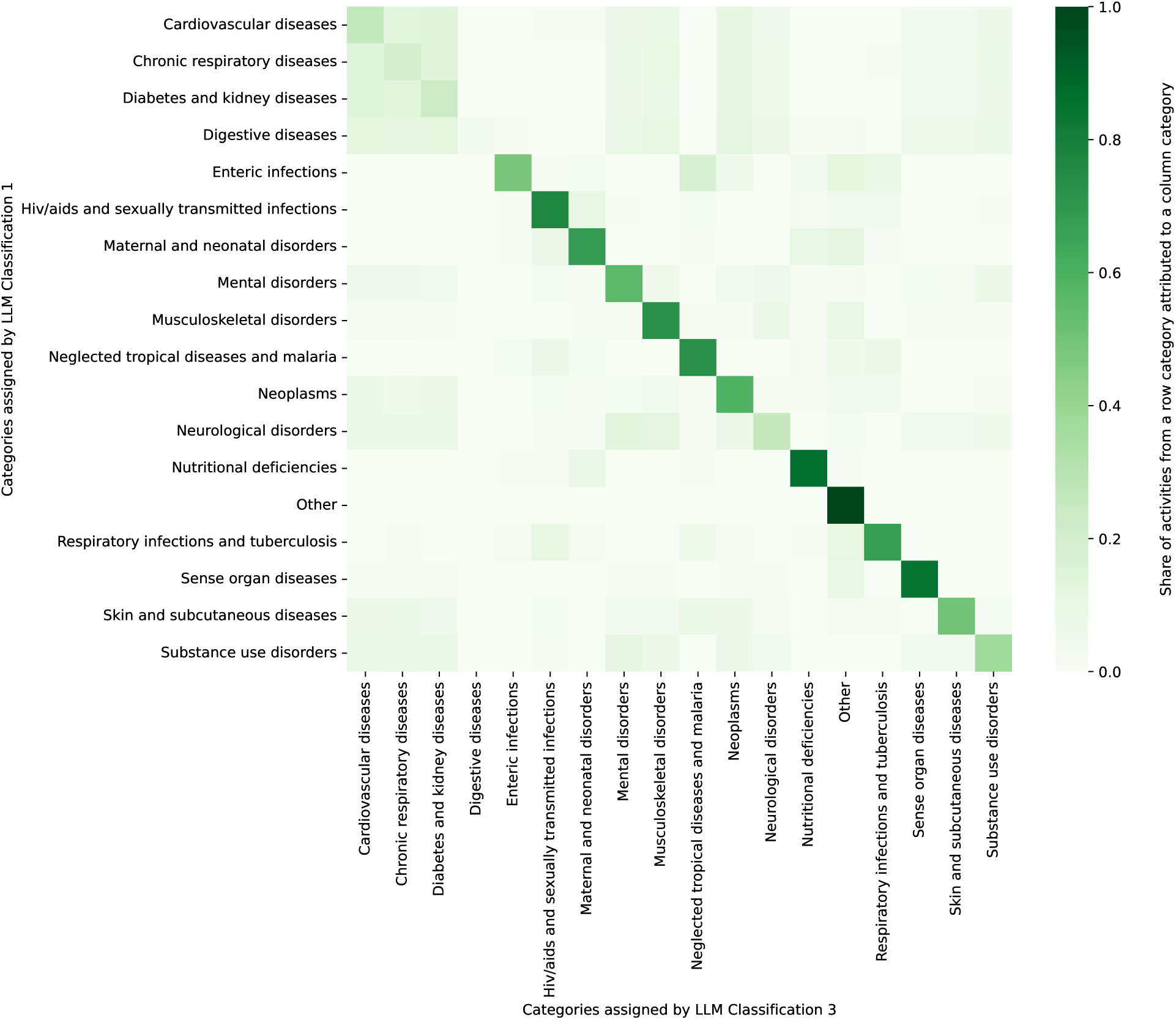
Comparison between classifications from different LLMs. The comparison shows how aid projects (“activities”) are classified into disease categories by our machine learning pipeline using different LLMs. LLM Classification 1 uses Llama-3.1-70B (rows), LLM Classification 3 uses gpt-4.1-mini-2025-04-14 (columns). Each cell reports the share of activities from a row category that is assigned to the corresponding column category, so rows sum to 1. Values on the diagonal indicate agreement between annotators for a given disease category, while off-diagonal cells indicate disagreement.

Additionally, we specifically addressed potential failure modes due to long project descriptions, negation, and short, non-English, and semantically ambiguous project descriptions. First, to evaluate potential context-window truncation and positional bias arising from concatenating the project title, short description, and long description into a single input, we implemented a chunk-based variant of the pipeline on the manual validation set. Specifically, we divided the textual inputs (project title, short description, long description) into four identically sized chunks, classified each chunk separately, and concatenated the chunks and classifications to a single text. The concatenated text was then classified using a tailored prompt (Supplementary Figure S19). The chunk-based classification did not yield a meaningful improvement over the original classification (overall accuracy of 0.72 and micro-averaged *F*_1_-score of 0.83; Supplementary Tables S8 and S9), suggesting that truncation or positional bias did not materially affect results.

Second, we constructed a negation-focused validation set to assess classification performance on projects with negated disease mentions. Using spaCy and NegEx and the 500 disease keywords from the keyword-based classifier, we scanned all projects previously classified as disease-related and flagged descriptions in which a disease keyword occurred in a negated context (e.g., “no tuberculosis”, “not HIV”, “excluding malaria”). This pipeline identified 2,589 projects with at least one negated disease mention. From these, we randomly sampled 500 projects, which were manually labeled and re-classified using a specialized negation-aware prompt that explicitly restricts labels to diseases positively targeted by the project (Supplementary Figure S20). We then compared the original and negation-aware LLM classifications against the manual labels (Supplementary Tables S12, S13 and S14). The negation-aware prompt did not yield a meaningful improvement on this subset (overall accuracy of 0.74 and micro-averaged *F*_1_-score of 0.83; Supplementary Tables S12, S13 and S14). In the full corpus of disease-related projects (*n* = 321,527), only 2,589 descriptions (approximately 0.8%) contained a negated disease mention, indicating that explicit negation is rare and setting an upper bound on cases where negation could influence classification. Third, we created an additional validation set of 2000 projects with non-English descriptions shorter than 100 characters, which were translated using Google Translate and manually annotated. We then evaluated the original pipeline on this short and non-English subset that may be potentially more ambiguous in the classification than the rest of the sample (Supplementary Tables S10 and S11), finding that overall performance remains comparable to the main validation set (overall accuracy of 0.92 and micro-averaged *F*_1_-score of 0.92), with misclassifications concentrated in inherently ambiguous descriptions.

**Fig. S19.**
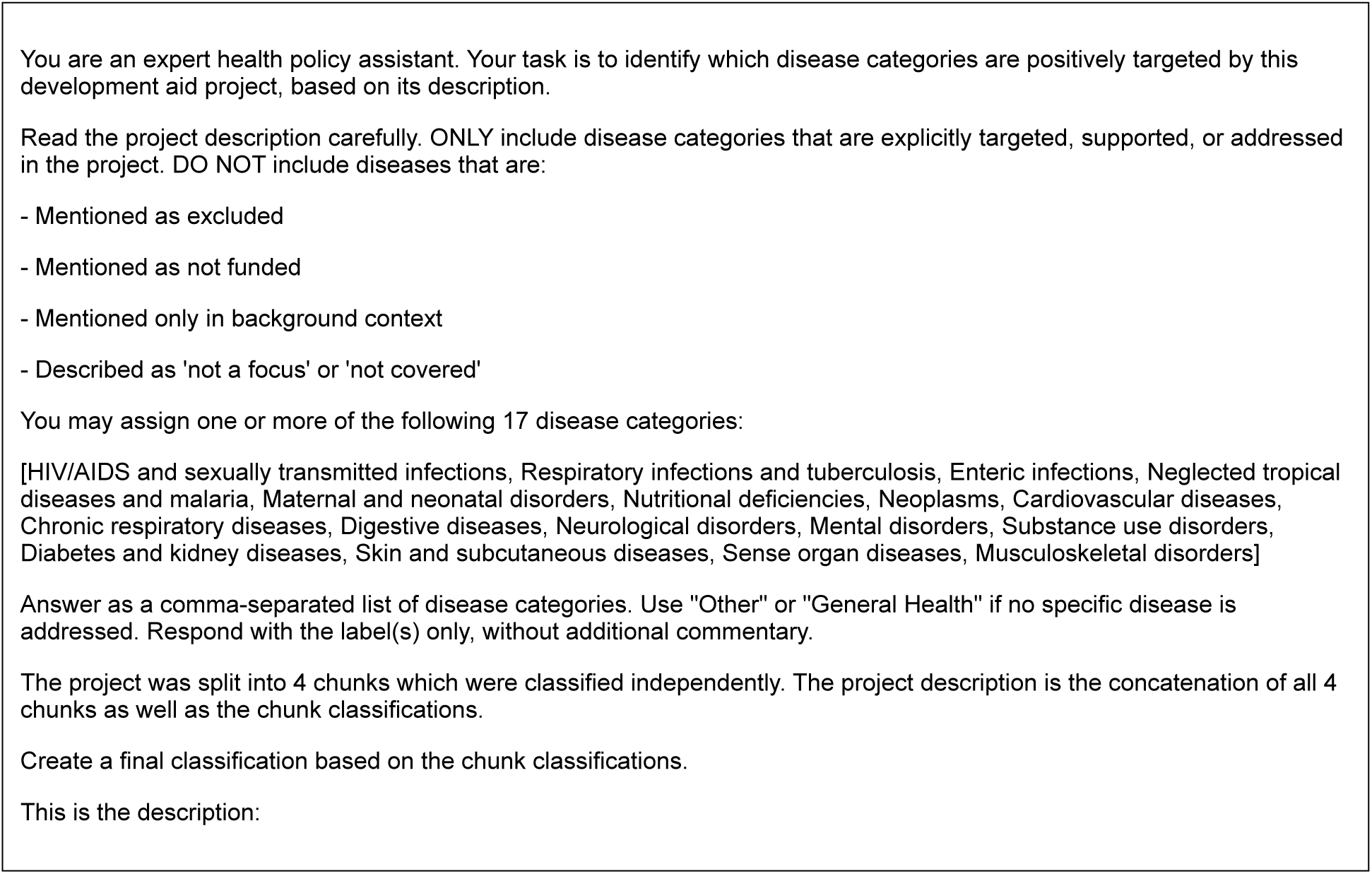
Prompt design for the chunk-based classification.

**Table S8:**
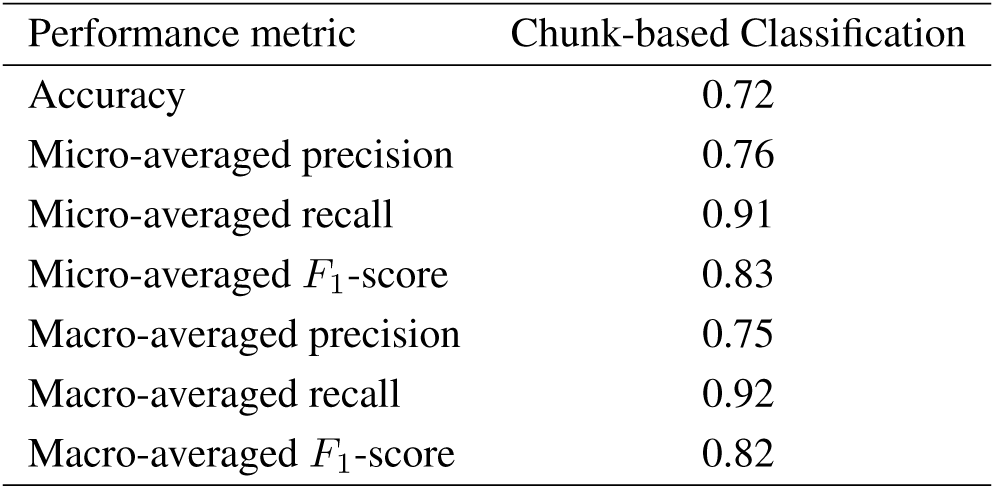
Overall classification performance using chunk-based classification.

**Table S9:** Classification performance by disease category using chunk-based classification.

| Disease category | Precision | Recall | True positives |
| --- | --- | --- | --- |
| Cardiovascular diseases | 0.81 | 0.94 | 47 |
| Chronic respiratory diseases | 0.69 | 0.92 | 24 |
| Diabetes and kidney diseases | 0.83 | 1.00 | 38 |
| Digestive diseases | 0.54 | 0.78 | 14 |
| Enteric infections | 0.69 | 0.78 | 49 |
| HIV/AIDS and STIs | 0.78 | 0.97 | 97 |
| Maternal and neonatal disorders | 0.64 | 0.93 | 68 |
| Mental disorders | 0.75 | 0.89 | 93 |
| Musculoskeletal disorders | 0.72 | 0.95 | 21 |
| Neglected tropical diseases and malaria | 0.91 | 0.99 | 143 |
| Neoplasms | 0.87 | 0.99 | 77 |
| Neurological disorders | 0.58 | 0.92 | 46 |
| Nutritional deficiencies | 0.73 | 0.94 | 134 |
| Other | 0.74 | 0.88 | 866 |
| Respiratory infections and tuberculosis | 0.87 | 0.97 | 175 |
| Sense organ diseases | 0.88 | 0.98 | 57 |
| Skin and subcutaneous diseases | 0.71 | 0.83 | 5 |
| Substance use disorders | 0.70 | 0.98 | 45 |

**Table S10:** Overall classification performance on short, ambiguous, and non-English project descriptions. Abbreviations: ML, machine learning.

| Performance metric | Our ML approach |
| --- | --- |
| Accuracy | 0.92 |
| Micro-averaged precision | 0.95 |
| Micro-averaged recall | 0.90 |
| Micro-averaged $F_1$-score | 0.92 |
| Macro-averaged precision | 0.92 |
| Macro-averaged recall | 0.81 |
| Macro-averaged $F_1$-score | 0.85 |

**Table S11:** Classification performance on short, ambiguous, and non-English project descriptions by disease category.

| Disease category | Precision | Recall | True positives |
| --- | --- | --- | --- |
| Cardiovascular diseases | 1.00 | 0.93 | 27 |
| Chronic respiratory diseases | 1.00 | 0.91 | 20 |
| Diabetes and kidney diseases | 1.00 | 1.00 | 18 |
| Digestive diseases | 1.00 | 0.58 | 11 |
| Enteric infections | 0.94 | 0.82 | 50 |
| HIV/AIDS and STIs | 0.99 | 0.98 | 371 |
| Maternal and neonatal disorders | 0.84 | 0.95 | 141 |
| Mental disorders | 0.94 | 0.94 | 94 |
| Musculoskeletal disorders | 0.92 | 0.44 | 12 |
| Neglected tropical diseases and malaria | 0.93 | 0.97 | 212 |
| Neoplasms | 0.97 | 0.75 | 33 |
| Neurological disorders | 0.83 | 0.70 | 19 |
| Nutritional deficiencies | 0.98 | 0.99 | 315 |
| Other | 1.00 | 0.91 | 485 |
| Respiratory infections and tuberculosis | 0.93 | 0.97 | 138 |
| Sense organ diseases | 0.98 | 0.80 | 59 |
| Skin and subcutaneous diseases | 0.50 | 0.11 | 2 |
| Substance use disorders | 0.93 | 0.89 | 41 |

**Fig. S20.**
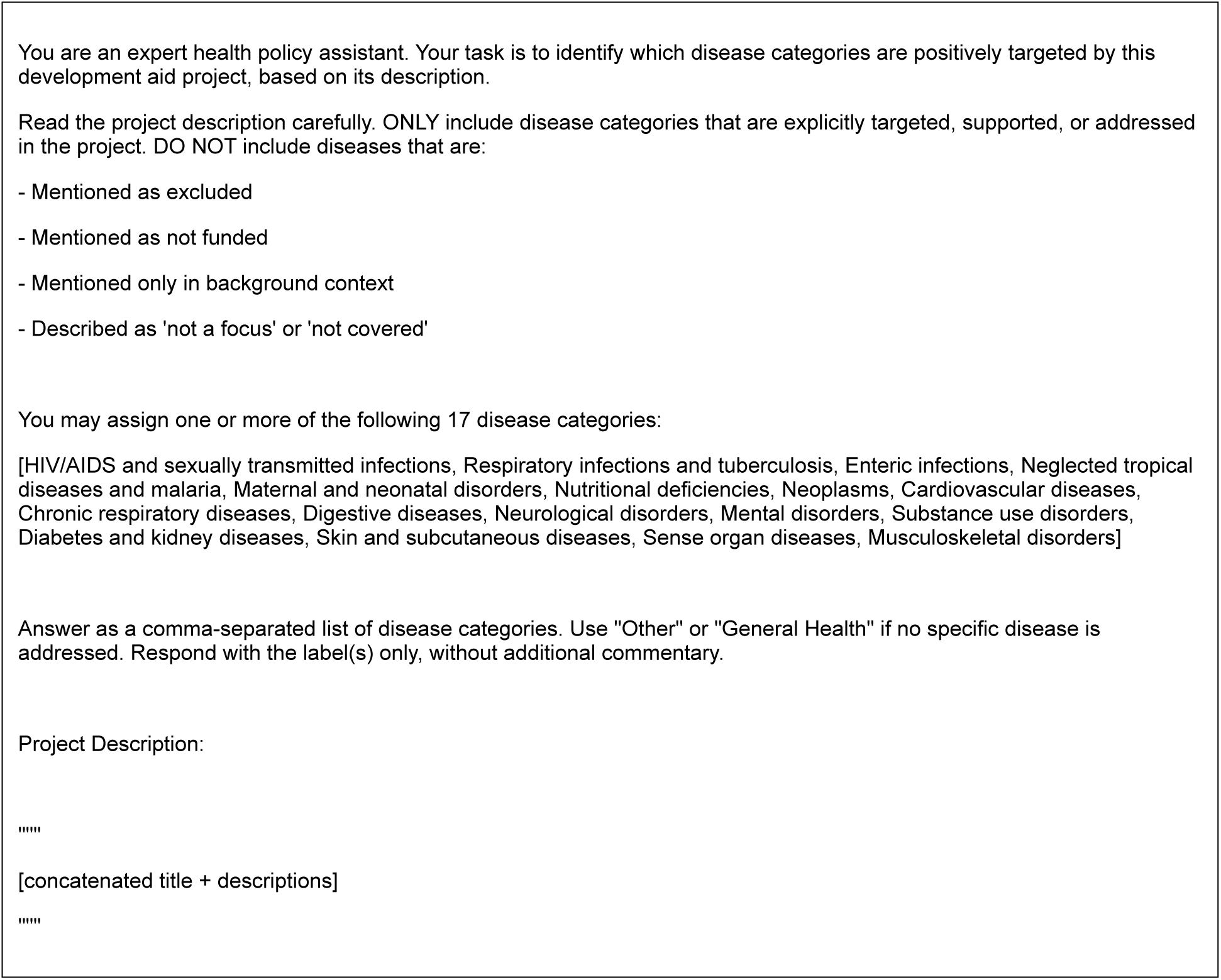
Prompt design for the negation-aware classification.

**Table S12:**
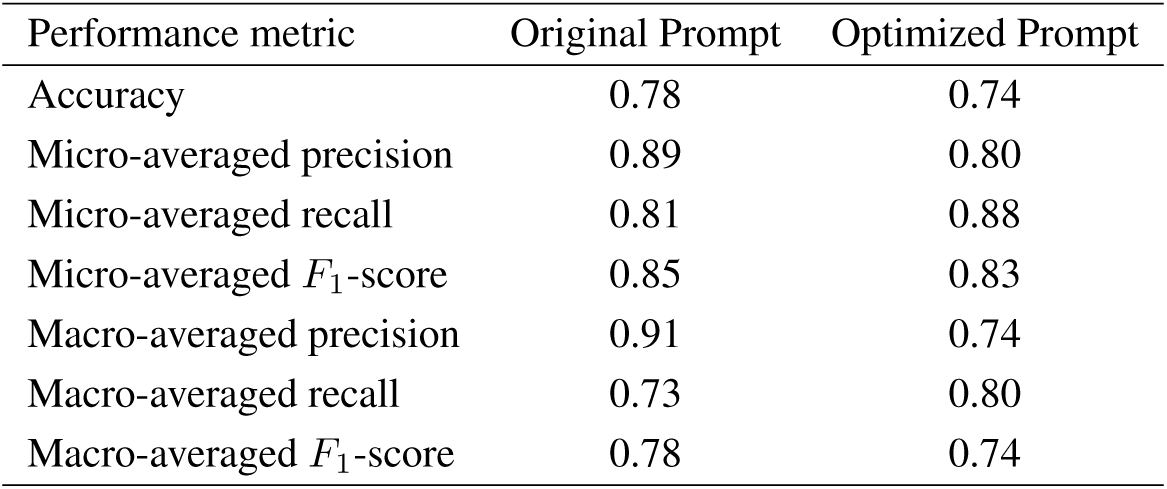
Overall classification performance on project descriptions including negation.

**Table S13:** Classification performance by disease category on project descriptions including negation using the original prompt.

| Disease category | Precision | Recall | True positives |
| --- | --- | --- | --- |
| Cardiovascular diseases | 0.92 | 0.92 | 12 |
| Chronic respiratory diseases | 1.00 | 0.38 | 3 |
| Diabetes and kidney diseases | 1.00 | 0.95 | 18 |
| Digestive diseases | 0.00 | 0.00 | 0 |
| Enteric infections | 0.64 | 0.58 | 14 |
| HIV/AIDS and STIs | 0.95 | 0.97 | 317 |
| Maternal and neonatal disorders | 0.85 | 0.50 | 17 |
| Mental disorders | 1.00 | 0.50 | 10 |
| Musculoskeletal disorders | 0.00 | 0.00 | 0 |
| Neglected tropical diseases and malaria | 0.95 | 0.98 | 125 |
| Neoplasms | 0.97 | 0.95 | 35 |
| Neurological disorders | 0.90 | 0.90 | 9 |
| Nutritional deficiencies | 0.88 | 0.87 | 84 |
| Other | 0.00 | 0.00 | 0 |
| Respiratory infections and tuberculosis | 0.68 | 0.86 | 77 |
| Sense organ diseases | 0.00 | 0.00 | 0 |
| Skin and subcutaneous diseases | 1.00 | 0.50 | 1 |
| Substance use disorders | 1.00 | 0.38 | 3 |

**Table S14:** Classification performance by disease category on project descriptions including negation using the negation-optimized prompt.

| Disease category | Precision | Recall | True positives |
| --- | --- | --- | --- |
| Cardiovascular diseases | 0.87 | 1.00 | 13 |
| Chronic respiratory diseases | 1.00 | 0.75 | 6 |
| Diabetes and kidney diseases | 0.86 | 0.95 | 18 |
| Digestive diseases | 0.00 | 0.00 | 0 |
| Enteric infections | 0.71 | 0.83 | 20 |
| HIV/AIDS and STIs | 0.91 | 1.00 | 326 |
| Maternal and neonatal disorders | 0.33 | 0.71 | 24 |
| Mental disorders | 0.56 | 0.75 | 15 |
| Musculoskeletal disorders | 0.00 | 0.00 | 0 |
| Neglected tropical diseases and malaria | 0.91 | 0.98 | 125 |
| Neoplasms | 0.88 | 1.00 | 37 |
| Neurological disorders | 0.75 | 0.90 | 9 |
| Nutritional deficiencies | 0.85 | 0.97 | 94 |
| Other | 0.33 | 0.03 | 2 |
| Respiratory infections and tuberculosis | 0.70 | 0.97 | 87 |
| Sense organ diseases | 1.00 | 0.33 | 1 |
| Skin and subcutaneous diseases | 0.67 | 1.00 | 2 |
| Substance use disorders | 0.45 | 0.62 | 5 |

### S3 Prompt design

The prompt was designed following best practices in prompt engineering and prior research [80–82]. It includes a short task description, a list of possible labels, and the description of the aid project to be classified. The prompt is designed to guide the LLM along the following reasoning process. (i) The LLM should first determine whether a project is health-related. If not (or if unclear), the LLM should classify the project as “Other”. (ii) If the project is health-related but broad and not related to a specific disease category, it should be classified as “General Health”. (iii) Only disease-specific projects are categorized into the 17 specific disease categories. To reduce false positives, we instructed the LLM to follow a conservative approach: when the classification was uncertain, it should default to “Other”. We found that this helps prevent the overestimation of funding for specific diseases by avoiding the misclassification of non-disease-related projects into disease categories. Similarly, we observed that introducing a category “General Health” helped improve classification performance. As a result, the LLM avoids forcing projects related to the broader health systems but that do not align with a specific disease into one of the narrow disease labels, which also minimized false positives. The full prompt is provided in Supplementary Figure S21.

**Fig. S21.**
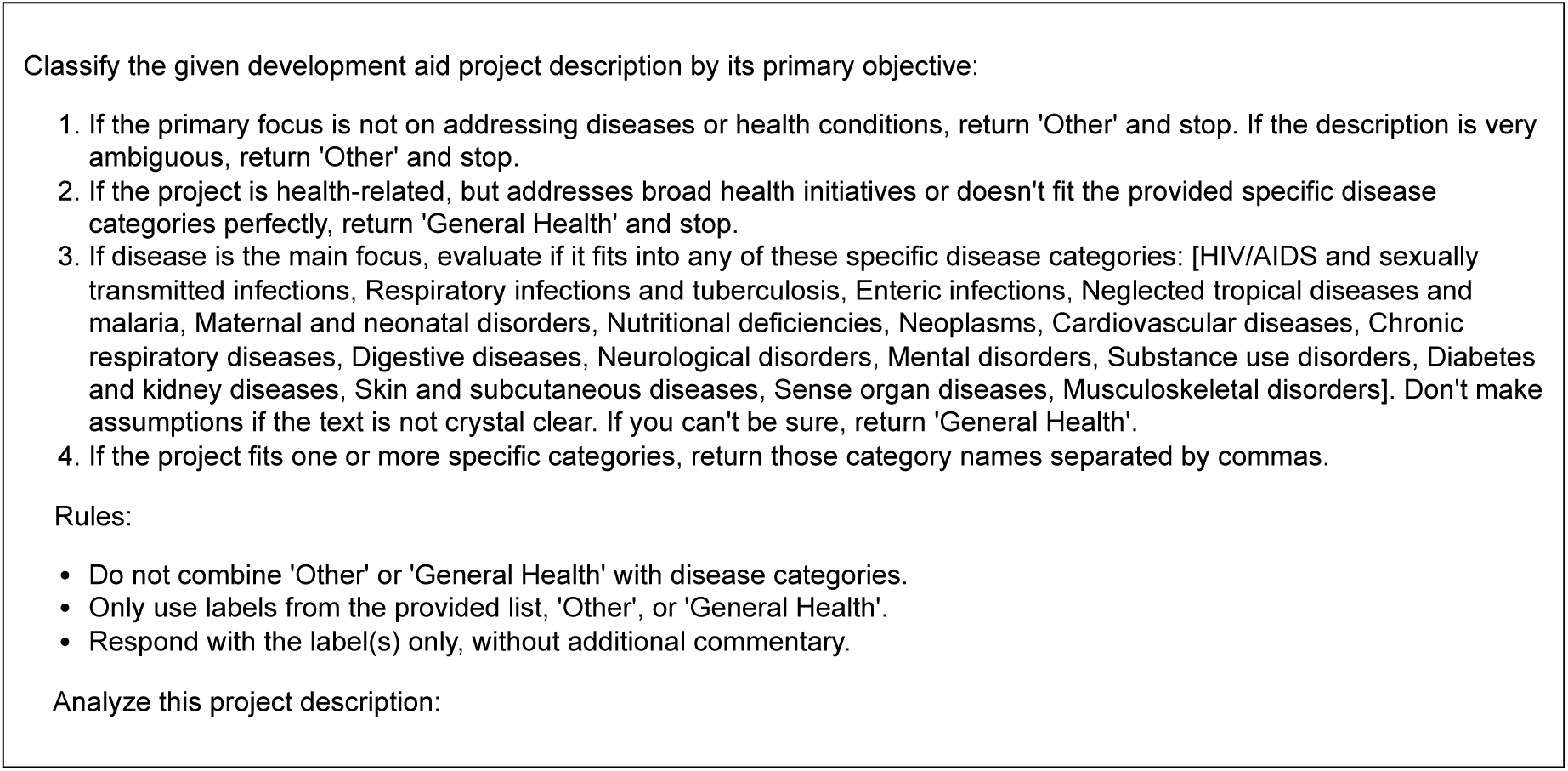
Prompt design. The prompt combines a short task description, a list of possible labels, and the description of the ODA project to be classified. It guides the LLM along several reasoning steps to prevent the misclassification of non-disease-related projects or projects that do not align with a specific disease, effectively reducing false positives.

### S4 Hyperparameters

The hyperparameters of our LLM approach were chosen as follows. We set the *temperature* and *top-p* values to 0.7 to balance deterministic output – balancing consistency in the disease classification with sufficient flexibility for edge cases. We specified *top-k* = 50 and a *repetition penalty* of 1.0 to effectively distinguish between closely related disease categories while preventing classification loops in complex multi-disease descriptions. The *maximum token output* was capped at 40, which provided sufficient length for our classification task while ensuring computational scalability.

**Table S15:**
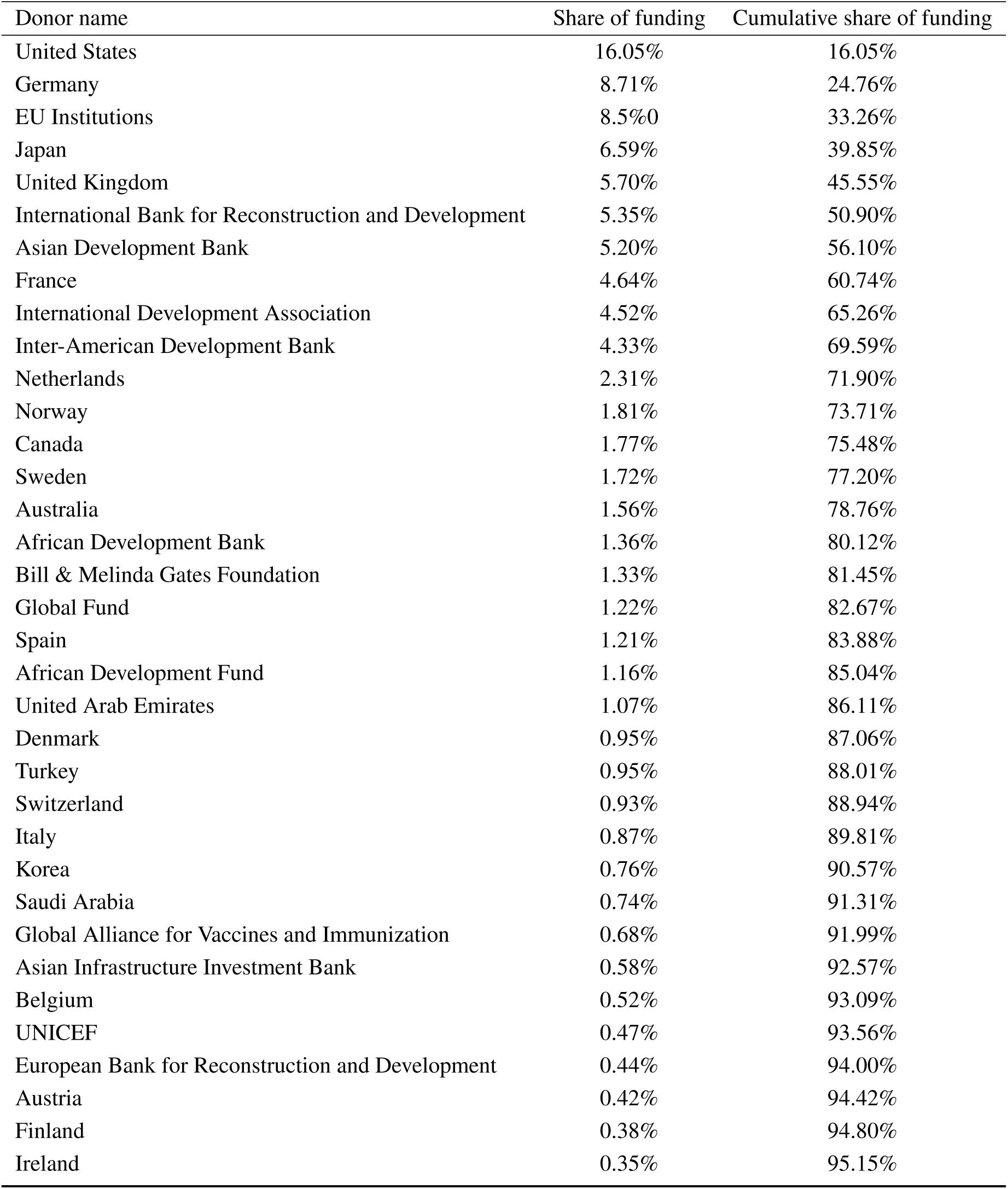
Share and cumulative share of global development funding by donor from 2000 to 2022. The top 35 donors of the 165 total donors contribute over 95% of the total funding.

### S5 Keyword list

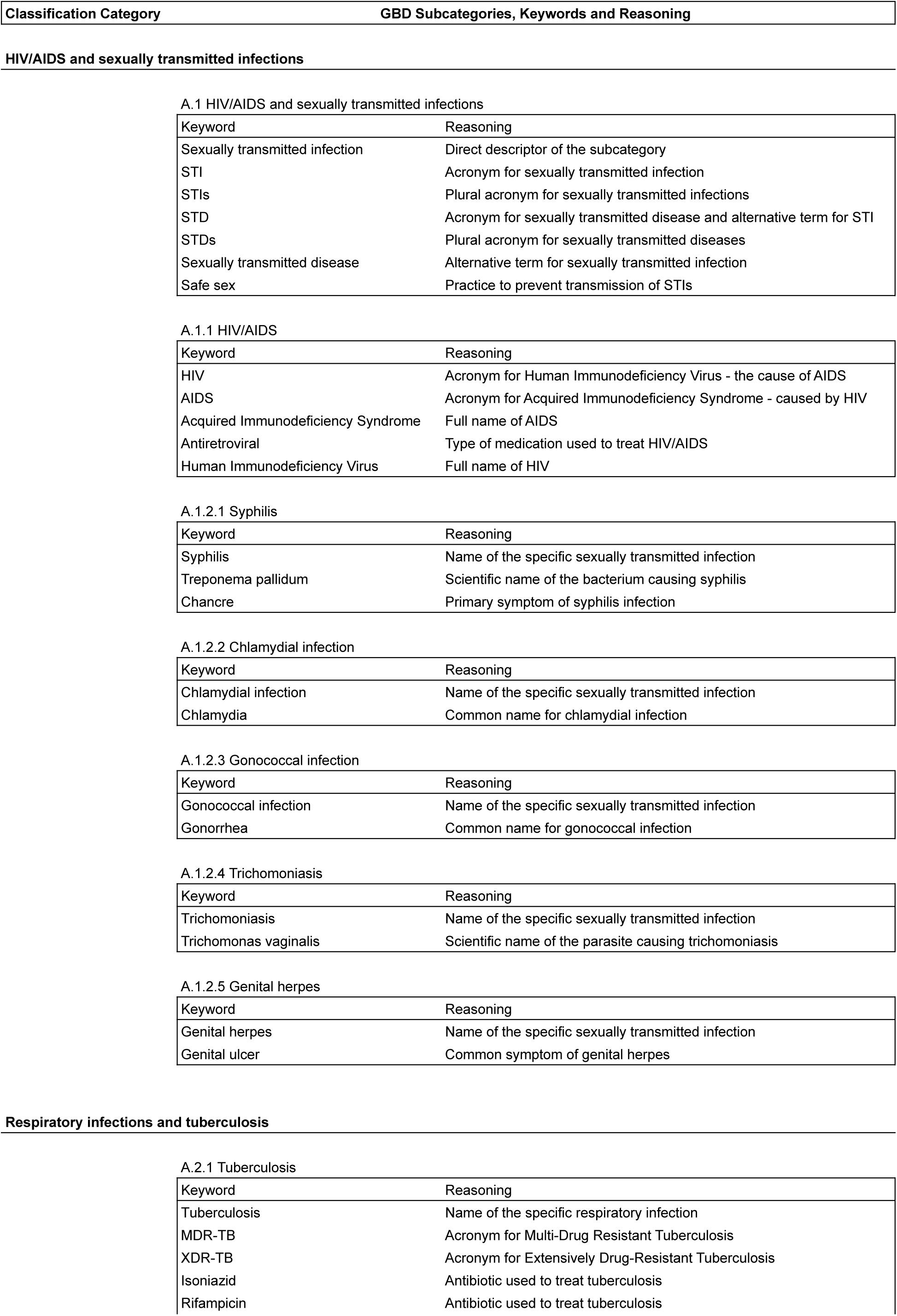
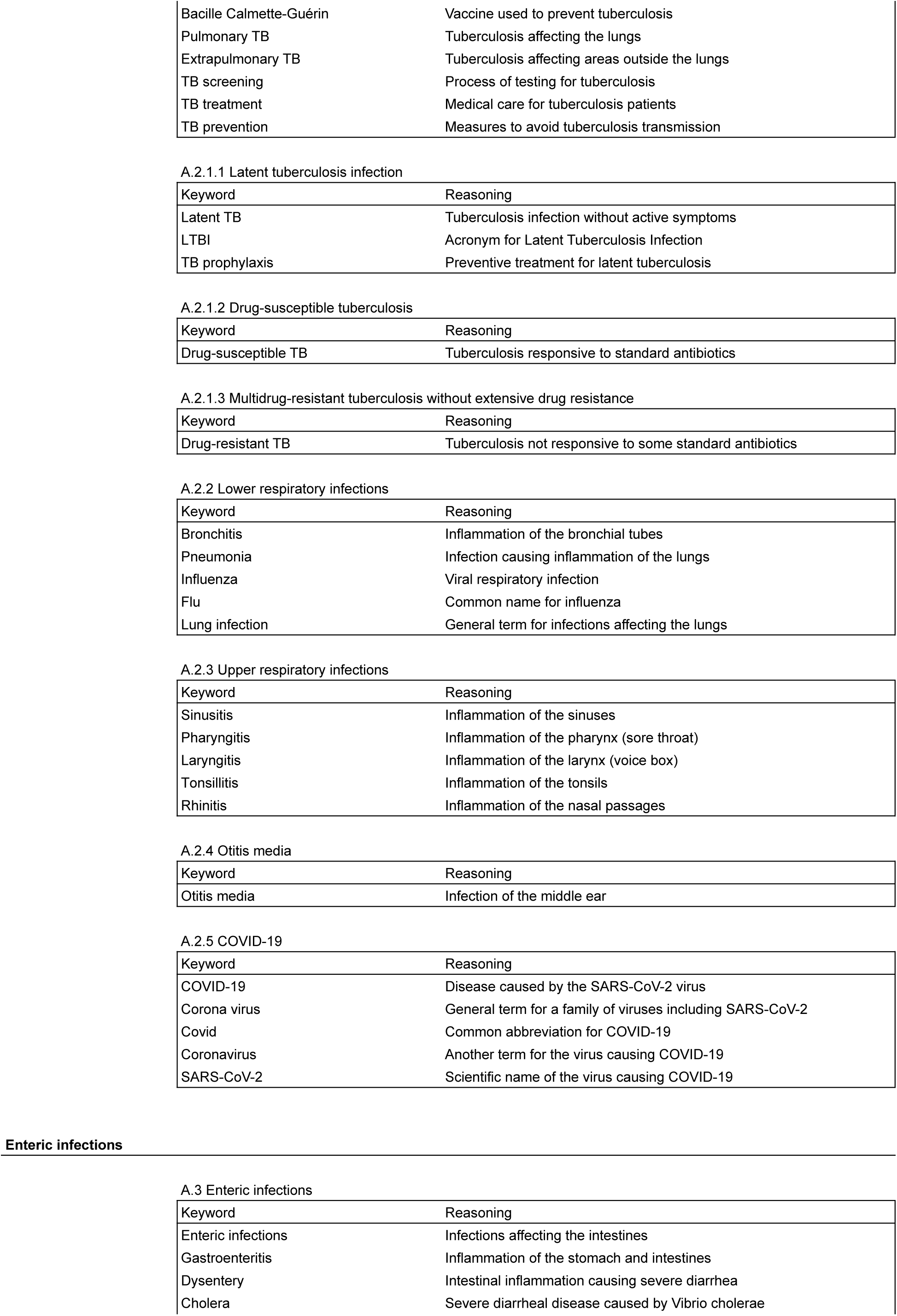
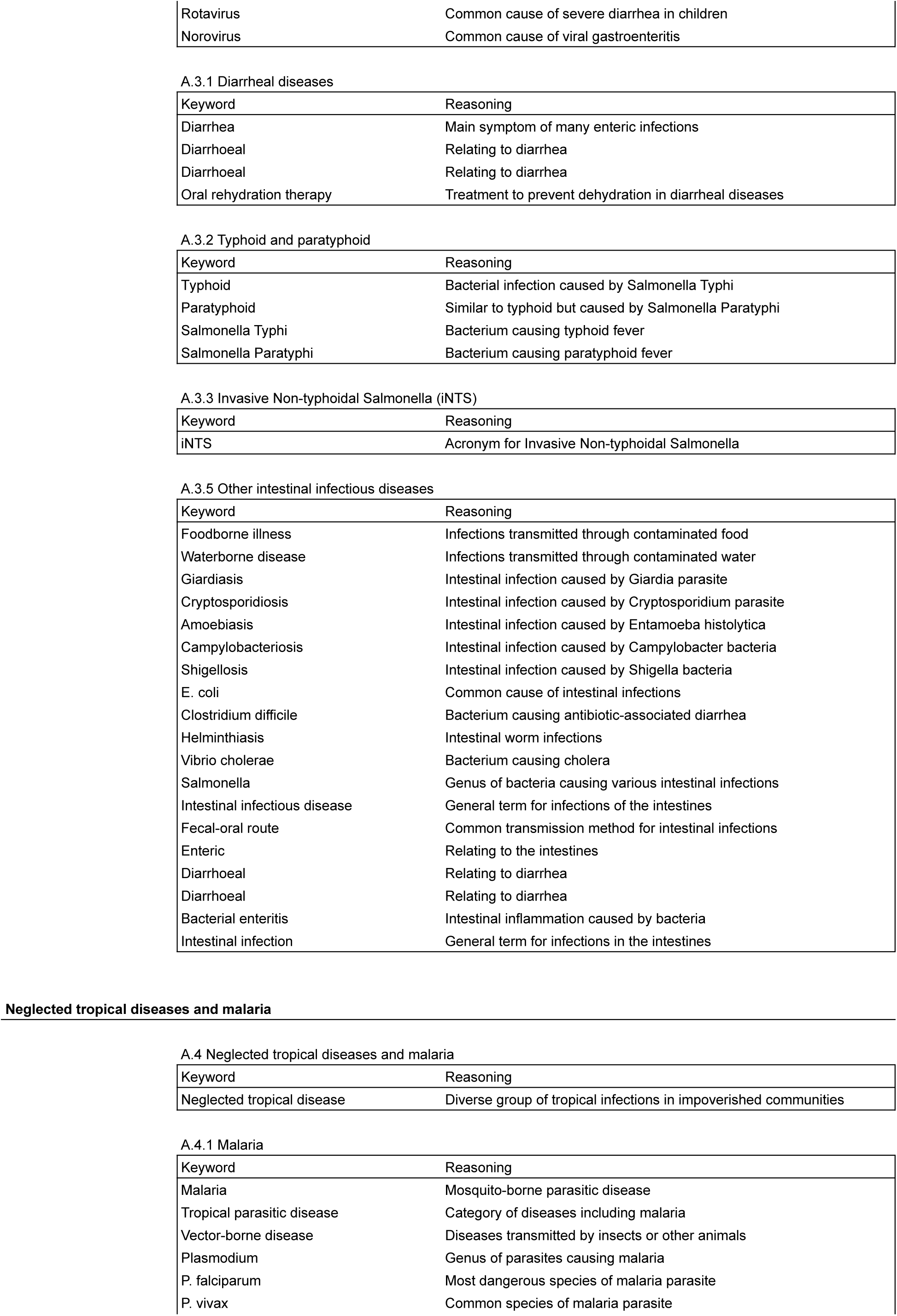
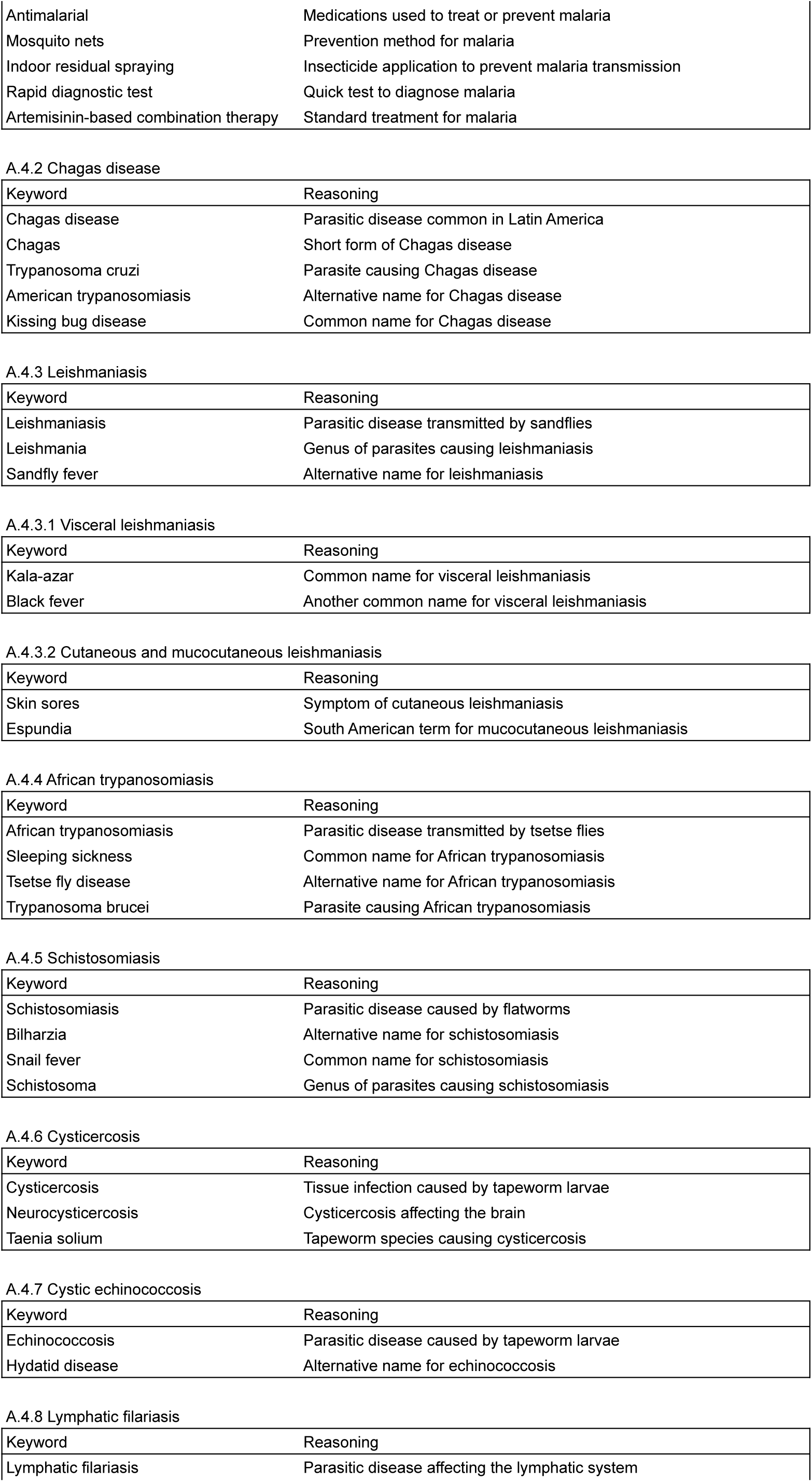
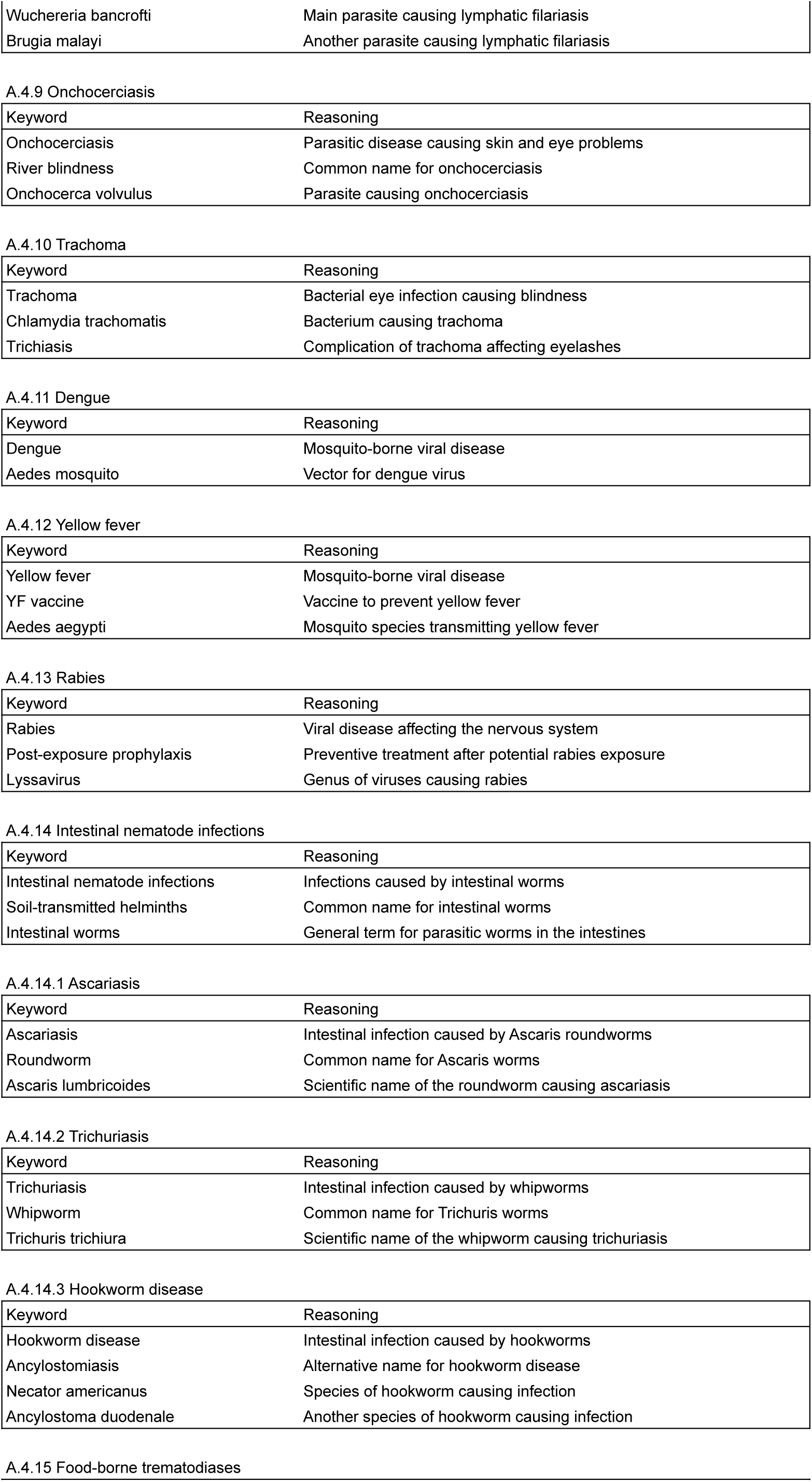
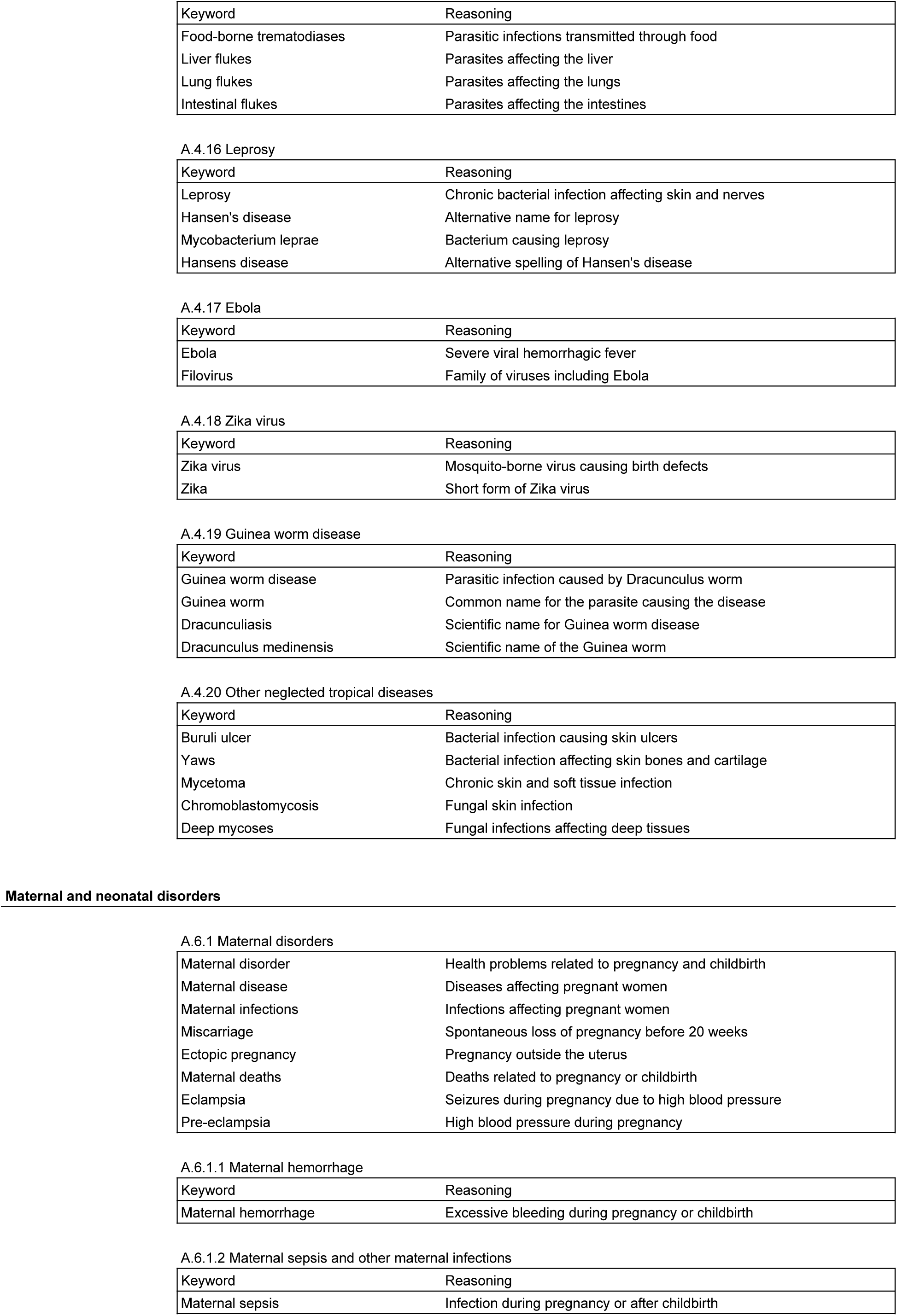
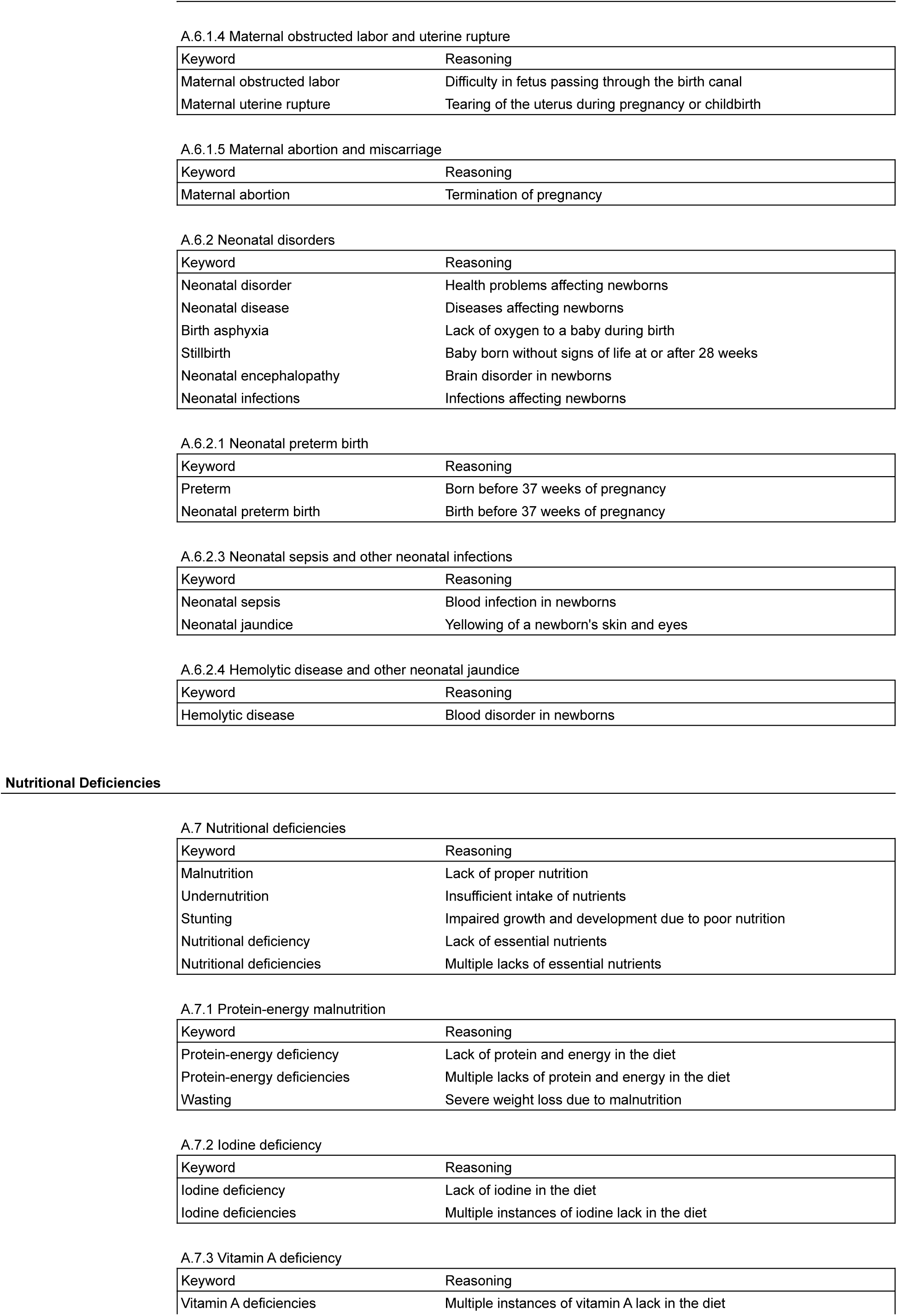
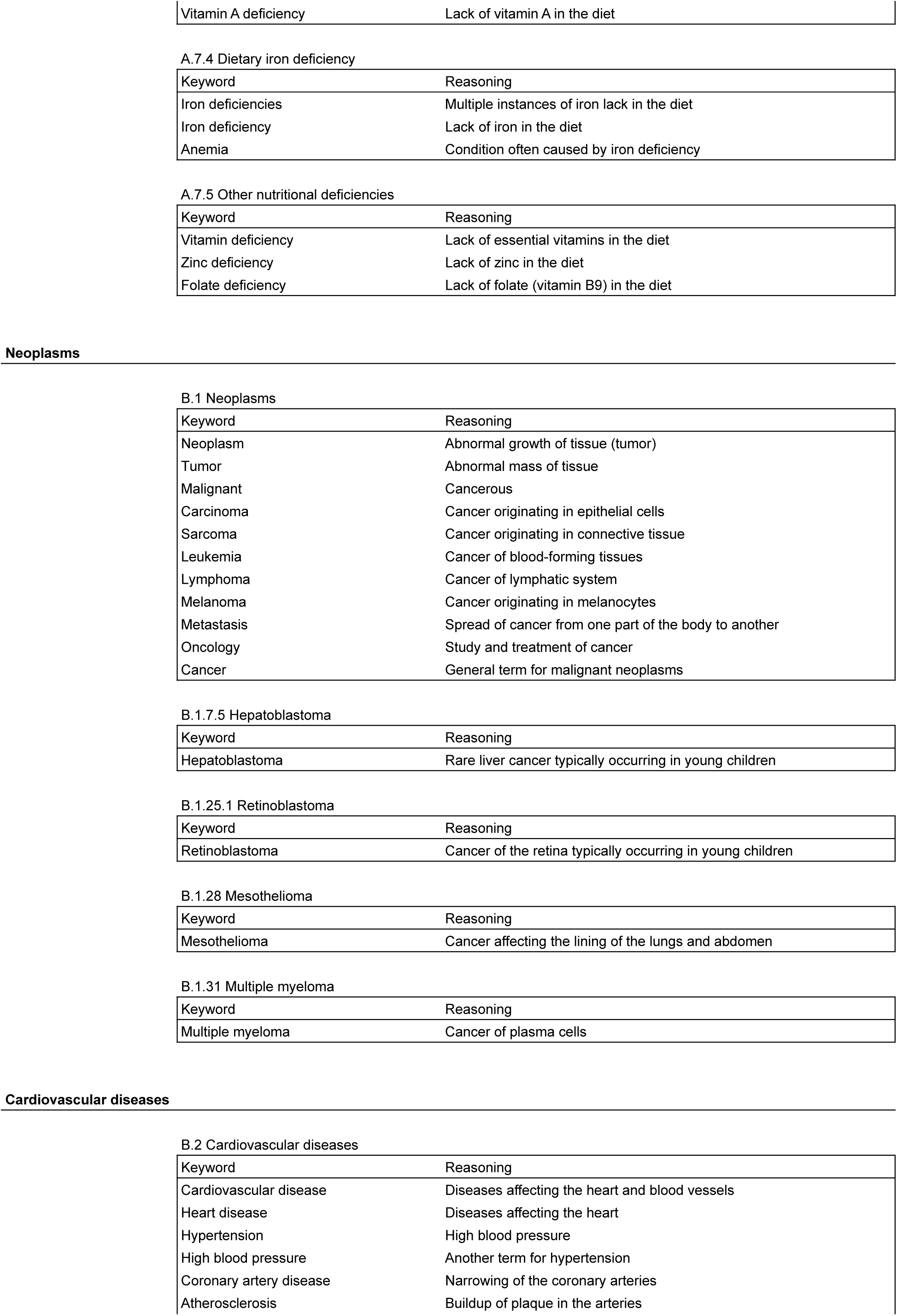
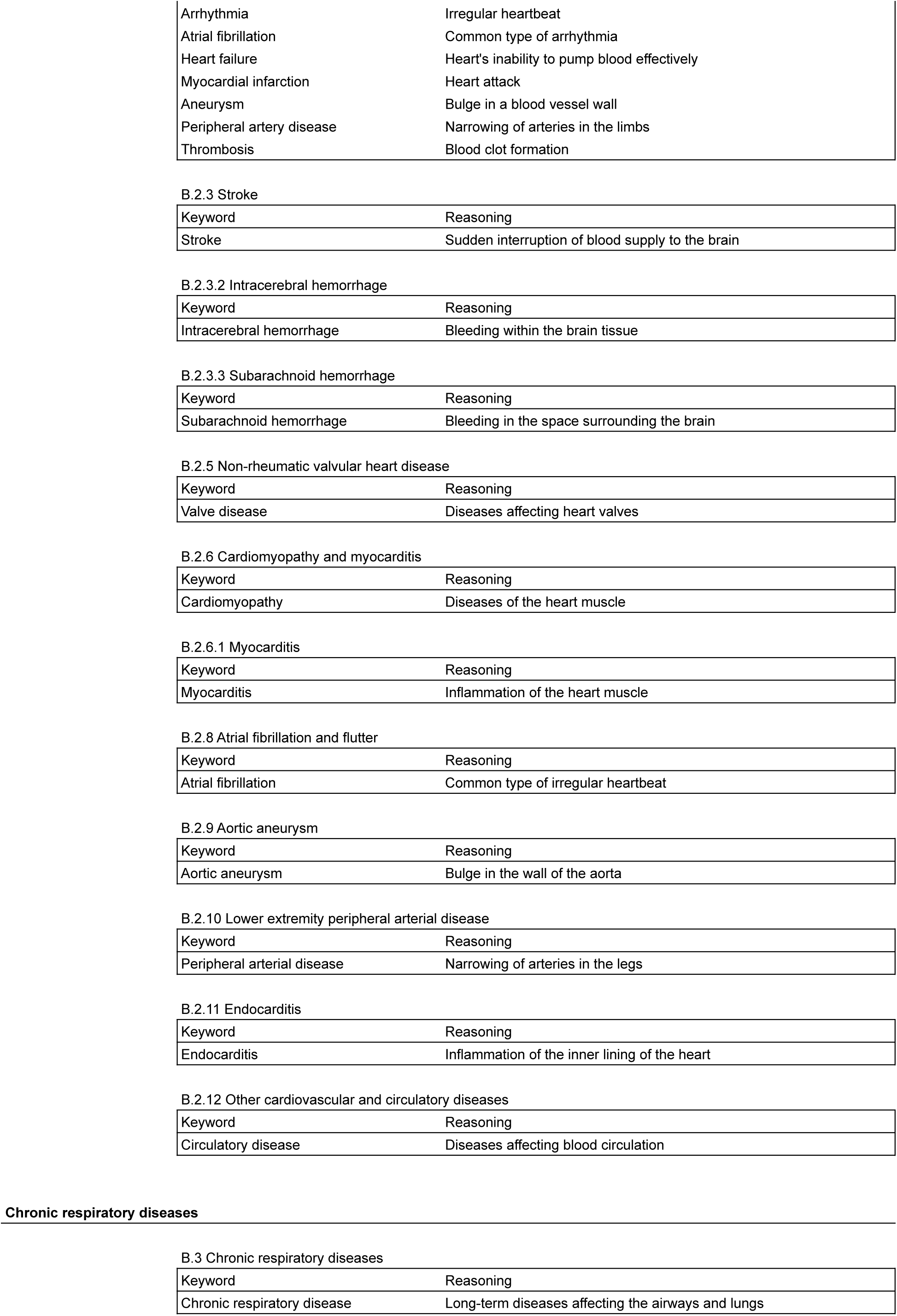
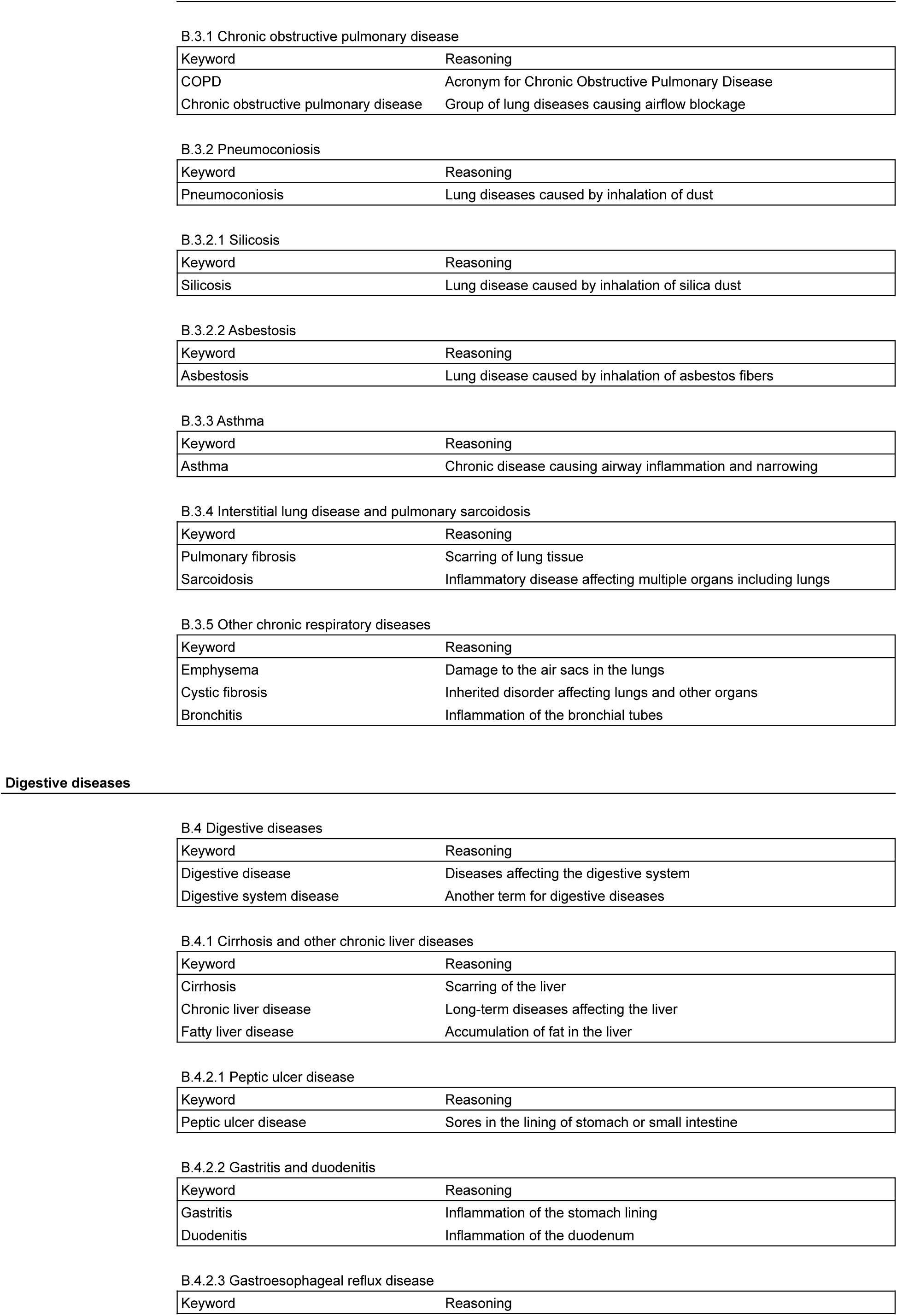
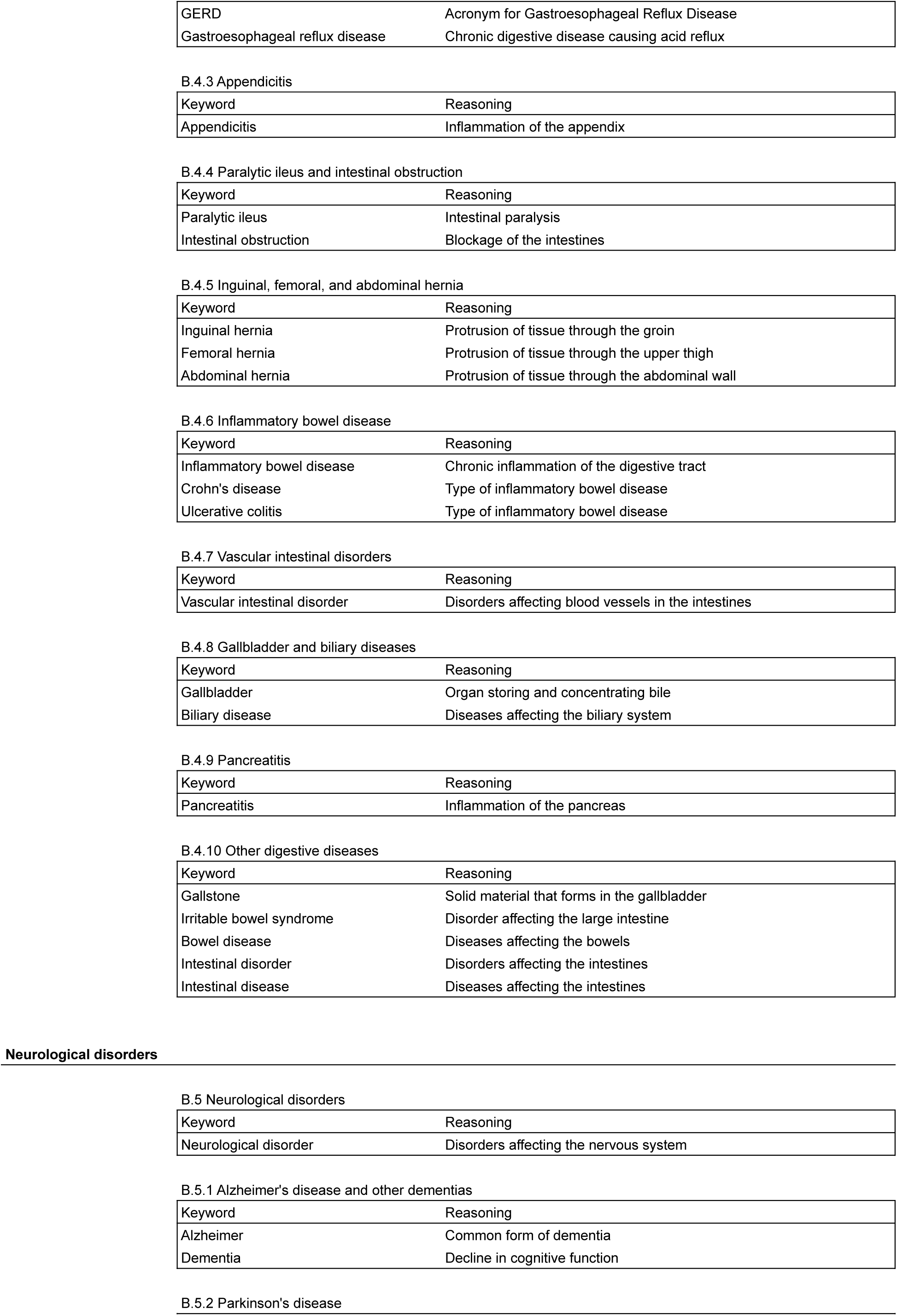
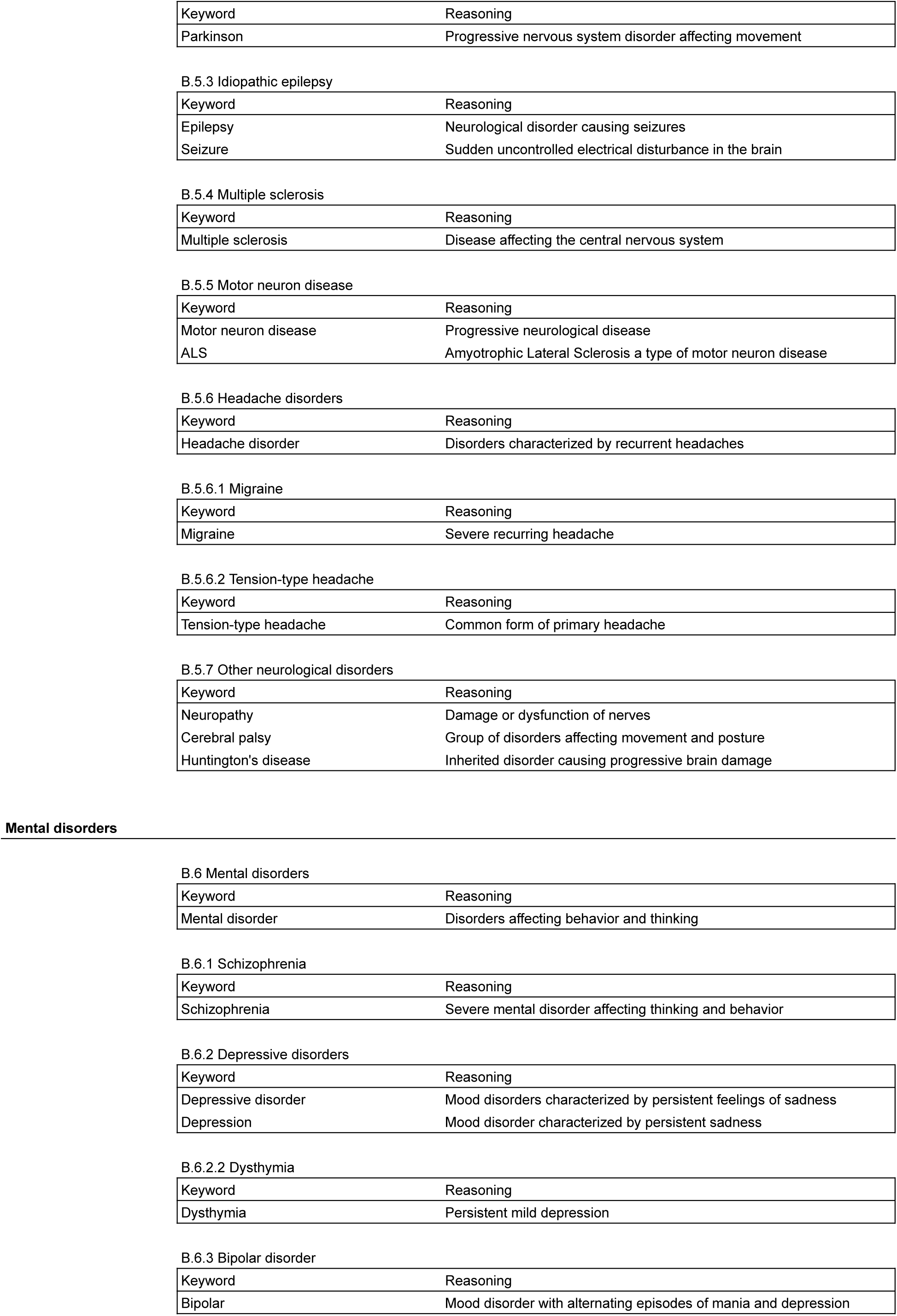
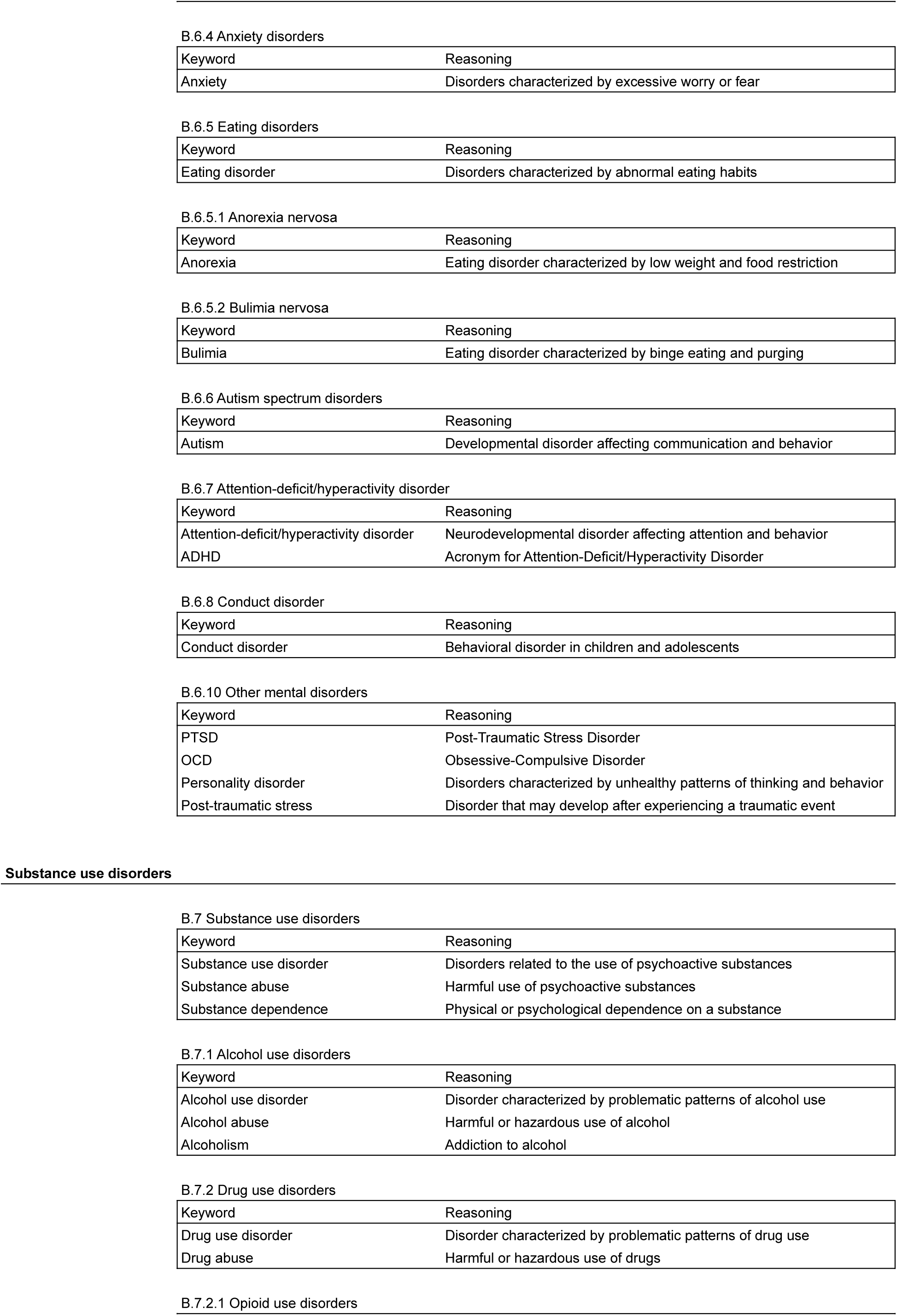
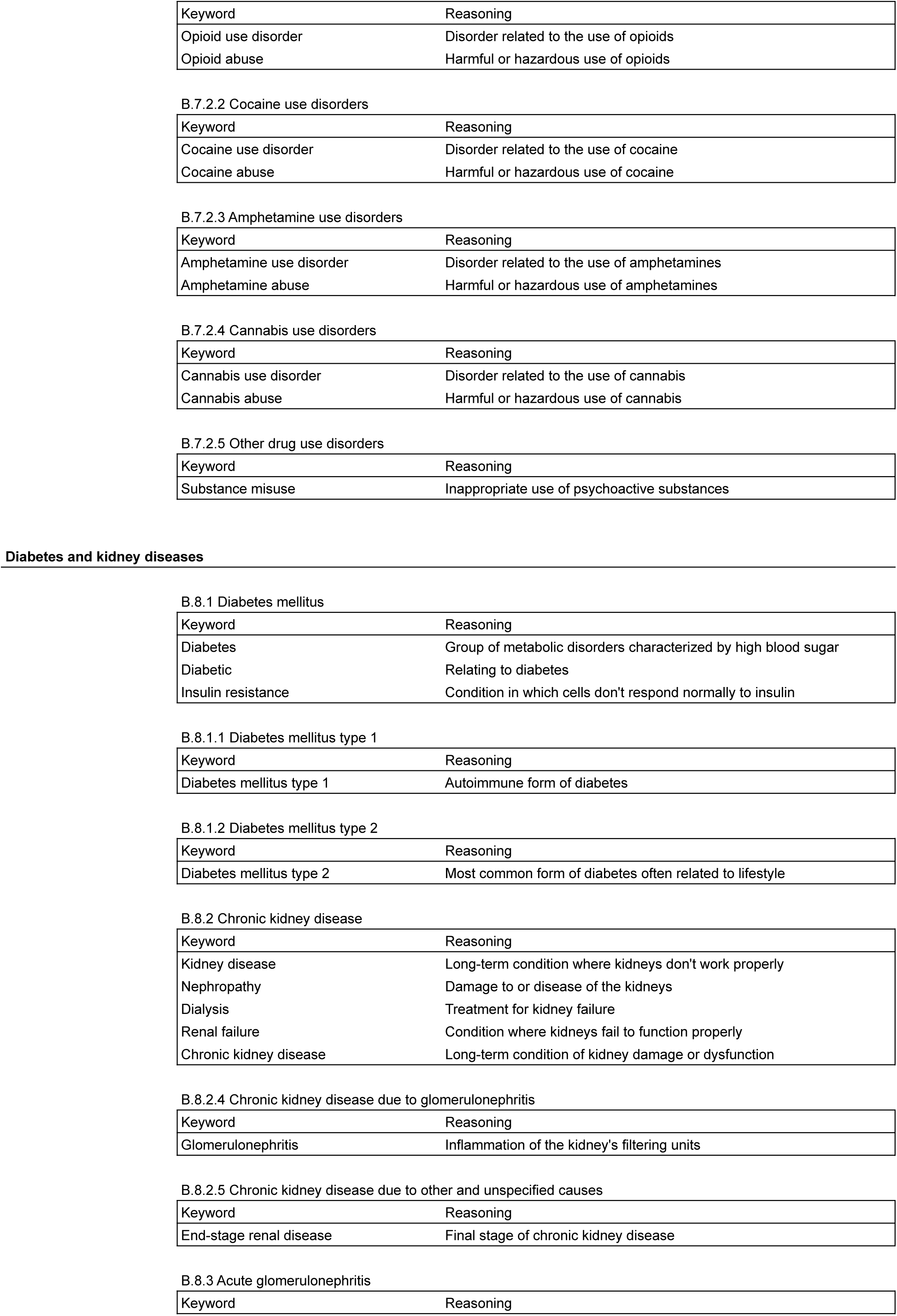
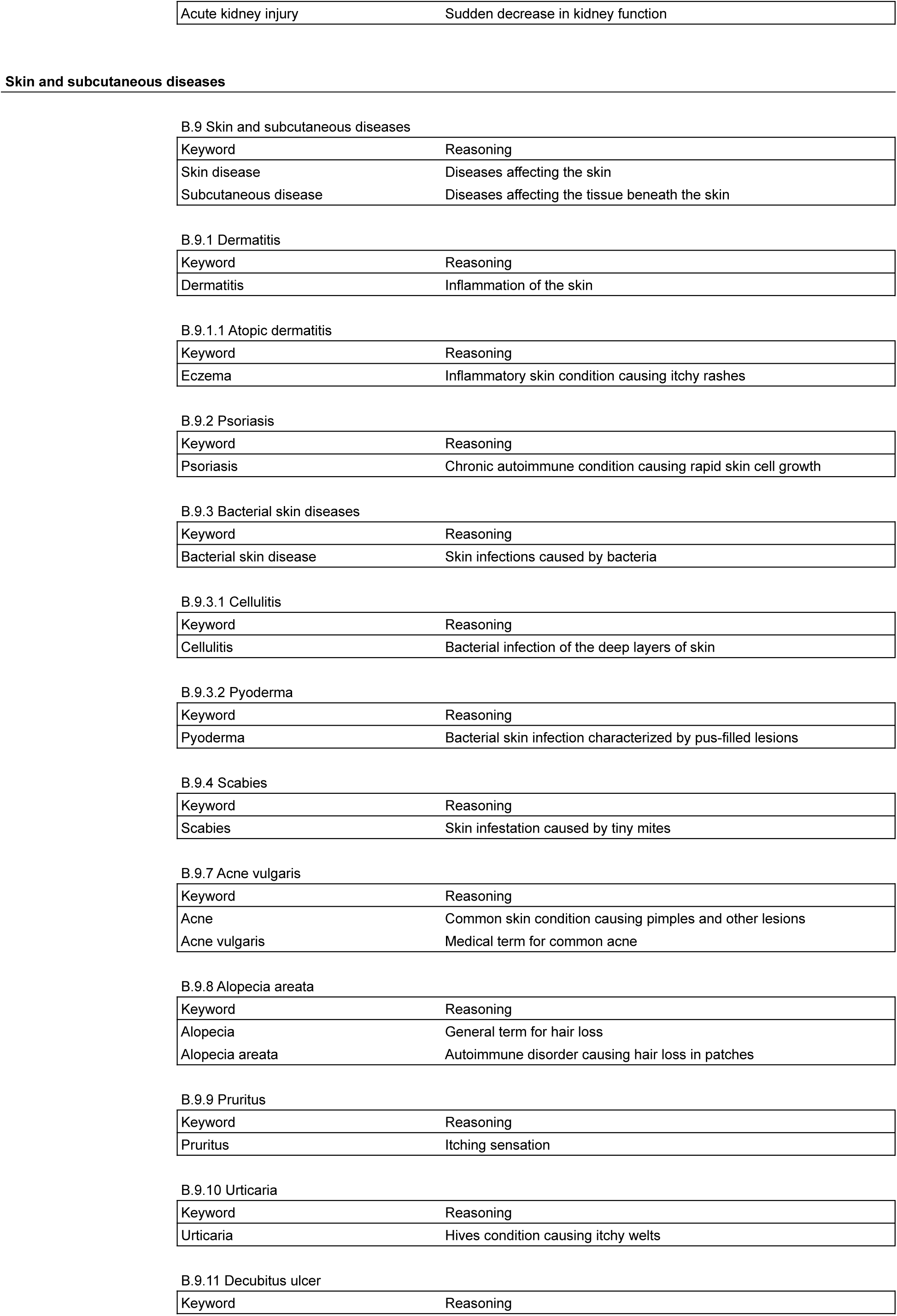
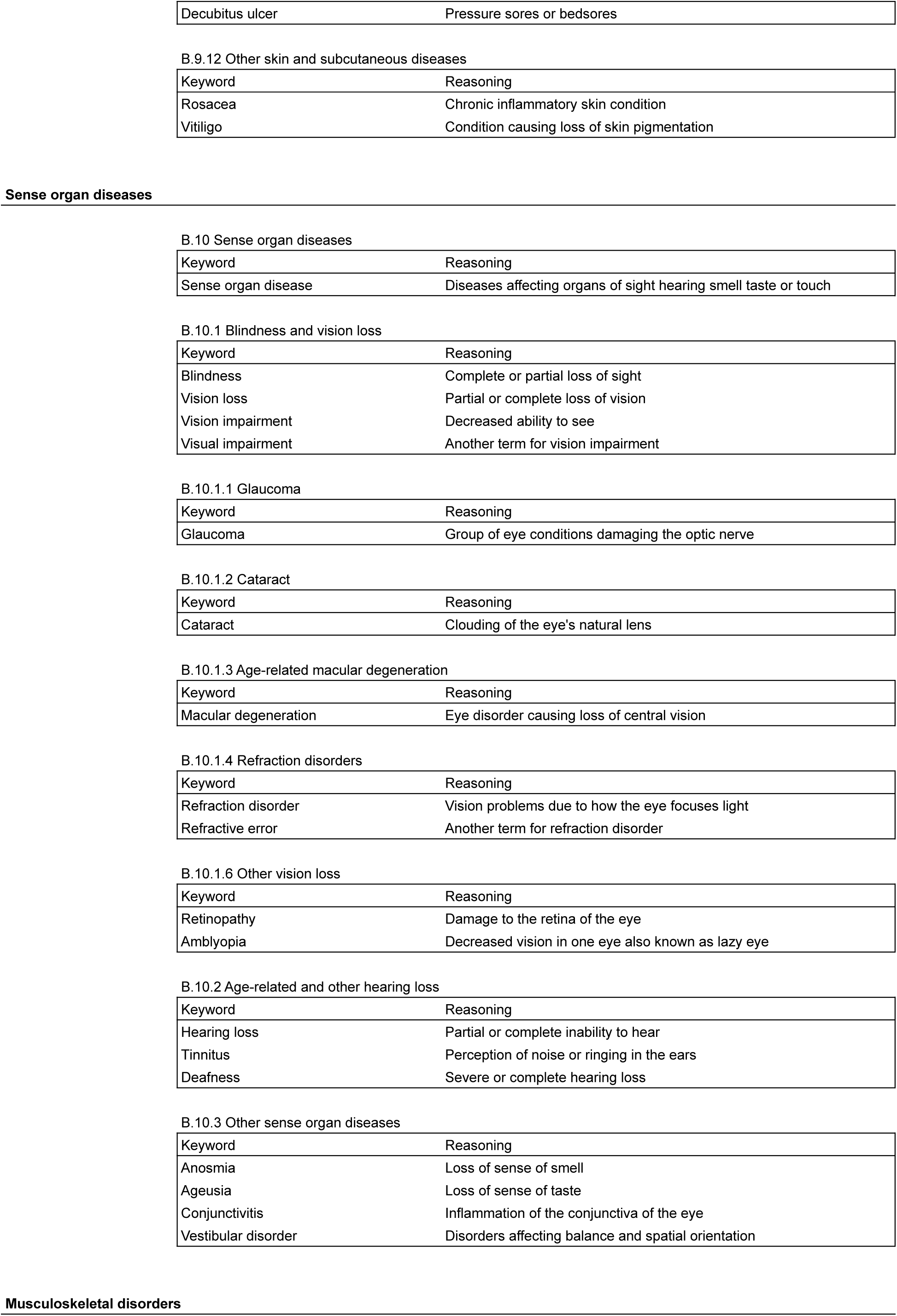
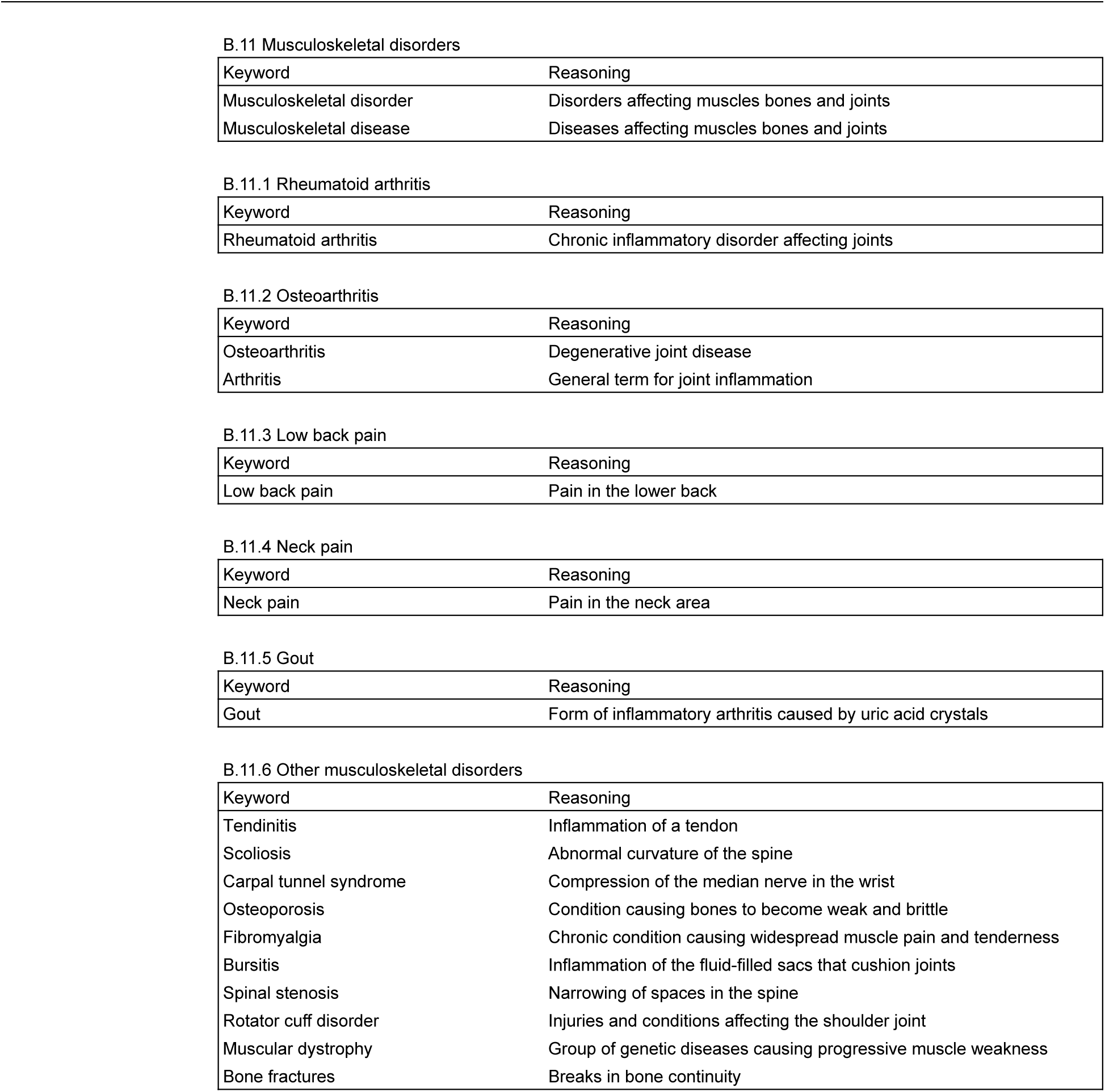

| Classification Category | GBD Subcategories, Keywords and Reasoning |
| --- | --- |

HIV/AIDS and sexually transmitted infections A.1 HIV/AIDS and sexually transmitted infections
| Keyword | Reasoning |
| --- | --- |
| Sexually transmitted infection | Direct descriptor of the subcategory |
| STI | Acronym for sexually transmitted infection |
| STIs | Plural acronym for sexually transmitted infections |
| STD | Acronym for sexually transmitted disease and alternative term for STI |
| STDs | Plural acronym for sexually transmitted diseases |
| Sexually transmitted disease | Alternative term for sexually transmitted infection |
| Safe sex | Practice to prevent transmission of STIs |

A.1.1 HIV/AIDS
| Keyword | Reasoning |
| --- | --- |
| HIV | Acronym for Human Immunodeficiency Virus - the cause of AIDS |
| AIDS | Acronym for Acquired Immunodeficiency Syndrome - caused by HIV |
| Acquired Immunodeficiency Syndrome | Full name of AIDS |
| Antiretroviral | Type of medication used to treat HIV/AIDS |
| Human Immunodeficiency Virus | Full name of HIV |

A.1.2.1 Syphilis
| Keyword | Reasoning |
| --- | --- |
| Syphilis | Name of the specific sexually transmitted infection |
| Treponema pallidum | Scientific name of the bacterium causing syphilis |
| Chancre | Primary symptom of syphilis infection |

A.1.2.2 Chlamydial infection
| Keyword | Reasoning |
| --- | --- |
| Chlamydial infection | Name of the specific sexually transmitted infection |
| Chlamydia | Common name for chlamydial infection |

A.1.2.3 Gonococcal infection
| Keyword | Reasoning |
| --- | --- |
| Gonococcal infection | Name of the specific sexually transmitted infection |
| Gonorrhea | Common name for gonococcal infection |

A.1.2.4 Trichomoniasis
| Keyword | Reasoning |
| --- | --- |
| Trichomoniasis | Name of the specific sexually transmitted infection |
| Trichomonas vaginalis | Scientific name of the parasite causing trichomoniasis |

A.1.2.5 Genital herpes
| Keyword | Reasoning |
| --- | --- |
| Genital herpes | Name of the specific sexually transmitted infection |
| Genital ulcer | Common symptom of genital herpes |

Respiratory infections and tuberculosis A.2.1 Tuberculosis
| Keyword | Reasoning |
| --- | --- |
| Tuberculosis | Name of the specific respiratory infection |
| MDR-TB | Acronym for Multi-Drug Resistant Tuberculosis |
| XDR-TB | Acronym for Extensively Drug-Resistant Tuberculosis |
| Isoniazid | Antibiotic used to treat tuberculosis |
| Rifampicin | Antibiotic used to treat tuberculosis |
| Bacille Calmette-Guérin | Vaccine used to prevent tuberculosis |
| Pulmonary TB | Tuberculosis affecting the lungs |
| Extrapulmonary TB | Tuberculosis affecting areas outside the lungs |
| TB screening | Process of testing for tuberculosis |
| TB treatment | Medical care for tuberculosis patients |
| TB prevention | Measures to avoid tuberculosis transmission |

A.2.1.1 Latent tuberculosis infection
| Keyword | Reasoning |
| --- | --- |
| Latent TB | Tuberculosis infection without active symptoms |
| LTBI | Acronym for Latent Tuberculosis Infection |
| TB prophylaxis | Preventive treatment for latent tuberculosis |

A.2.1.2 Drug-susceptible tuberculosis
| Keyword | Reasoning |
| --- | --- |
| Drug-susceptible TB | Tuberculosis responsive to standard antibiotics |

A.2.1.3 Multidrug-resistant tuberculosis without extensive drug resistance
| Keyword | Reasoning |
| --- | --- |
| Drug-resistant TB | Tuberculosis not responsive to some standard antibiotics |

A.2.2 Lower respiratory infections
| Keyword | Reasoning |
| --- | --- |
| Bronchitis | Inflammation of the bronchial tubes |
| Pneumonia | Infection causing inflammation of the lungs |
| Influenza | Viral respiratory infection |
| Flu | Common name for influenza |
| Lung infection | General term for infections affecting the lungs |

A.2.3 Upper respiratory infections
| Keyword | Reasoning |
| --- | --- |
| Sinusitis | Inflammation of the sinuses |
| Pharyngitis | Inflammation of the pharynx (sore throat) |
| Laryngitis | Inflammation of the larynx (voice box) |
| Tonsillitis | Inflammation of the tonsils |
| Rhinitis | Inflammation of the nasal passages |

A.2.4 Otitis media
| Keyword | Reasoning |
| --- | --- |
| Otitis media | Infection of the middle ear |

A.2.5 COVID-19
| Keyword | Reasoning |
| --- | --- |
| COVID-19 | Disease caused by the SARS-CoV-2 virus |
| Corona virus | General term for a family of viruses including SARS-CoV-2 |
| Covid | Common abbreviation for COVID-19 |
| Coronavirus | Another term for the virus causing COVID-19 |
| SARS-CoV-2 | Scientific name of the virus causing COVID-19 |

Enteric infections A.3 Enteric infections
| Keyword | Reasoning |
| --- | --- |
| Enteric infections | Infections affecting the intestines |
| Gastroenteritis | Inflammation of the stomach and intestines |
| Dysentery | Intestinal inflammation causing severe diarrhea |
| Cholera | Severe diarrheal disease caused by *Vibrio cholerae* |
| Rotavirus | Common cause of severe diarrhea in children |
| Norovirus | Common cause of viral gastroenteritis |

A.3.1 Diarrheal diseases
| Keyword | Reasoning |
| --- | --- |
| Diarrhea | Main symptom of many enteric infections |
| Diarrhoeal | Relating to diarrhea |
| Diarrhoeal | Relating to diarrhea |
| Oral rehydration therapy | Treatment to prevent dehydration in diarrheal diseases |

A.3.2 Typhoid and paratyphoid
| Keyword | Reasoning |
| --- | --- |
| Typhoid | Bacterial infection caused by Salmonella Typhi |
| Paratyphoid | Similar to typhoid but caused by Salmonella Paratyphi |
| Salmonella Typhi | Bacterium causing typhoid fever |
| Salmonella Paratyphi | Bacterium causing paratyphoid fever |

A.3.3 Invasive Non-typhoidal Salmonella (iNTS)
| Keyword | Reasoning |
| --- | --- |
| iNTS | Acronym for Invasive Non-typhoidal Salmonella |

A.3.5 Other intestinal infectious diseases
| Keyword | Reasoning |
| --- | --- |
| Foodborne illness | Infections transmitted through contaminated food |
| Waterborne disease | Infections transmitted through contaminated water |
| Giardiasis | Intestinal infection caused by Giardia parasite |
| Cryptosporidiosis | Intestinal infection caused by Cryptosporidium parasite |
| Amoebiasis | Intestinal infection caused by Entamoeba histolytica |
| Campylobacteriosis | Intestinal infection caused by Campylobacter bacteria |
| Shigellosis | Intestinal infection caused by Shigella bacteria |
| E. coli | Common cause of intestinal infections |
| Clostridium difficile | Bacterium causing antibiotic-associated diarrhea |
| Helminthiasis | Intestinal worm infections |
| Vibrio cholerae | Bacterium causing cholera |
| Salmonella | Genus of bacteria causing various intestinal infections |
| Intestinal infectious disease | General term for infections of the intestines |
| Fecal-oral route | Common transmission method for intestinal infections |
| Enteric | Relating to the intestines |
| Diarrhoeal | Relating to diarrhea |
| Diarrhoeal | Relating to diarrhea |
| Bacterial enteritis | Intestinal inflammation caused by bacteria |
| Intestinal infection | General term for infections in the intestines |

Neglected tropical diseases and malaria A.4 Neglected tropical diseases and malaria
| Keyword | Reasoning |
| --- | --- |
| Neglected tropical disease | Diverse group of tropical infections in impoverished communities |

A.4.1 Malaria
| Keyword | Reasoning |
| --- | --- |
| Malaria | Mosquito-borne parasitic disease |
| Tropical parasitic disease | Category of diseases including malaria |
| Vector-borne disease | Diseases transmitted by insects or other animals |
| Plasmodium | Genus of parasites causing malaria |
| P. falciparum | Most dangerous species of malaria parasite |
| P. vivax | Common species of malaria parasite |
| Antimalarial | Medications used to treat or prevent malaria |
| Mosquito nets | Prevention method for malaria |
| Indoor residual spraying | Insecticide application to prevent malaria transmission |
| Rapid diagnostic test | Quick test to diagnose malaria |
| Artemisinin-based combination therapy | Standard treatment for malaria |

A.4.2 Chagas disease
| Keyword | Reasoning |
| --- | --- |
| Chagas disease | Parasitic disease common in Latin America |
| Chagas | Short form of Chagas disease |
| Trypanosoma cruzi | Parasite causing Chagas disease |
| American trypanosomiasis | Alternative name for Chagas disease |
| Kissing bug disease | Common name for Chagas disease |

A.4.3 Leishmaniasis
| Keyword | Reasoning |
| --- | --- |
| Leishmaniasis | Parasitic disease transmitted by sandflies |
| Leishmania | Genus of parasites causing leishmaniasis |
| Sandfly fever | Alternative name for leishmaniasis |

A.4.3.1 Visceral leishmaniasis
| Keyword | Reasoning |
| --- | --- |
| Kala-azar | Common name for visceral leishmaniasis |
| Black fever | Another common name for visceral leishmaniasis |

A.4.3.2 Cutaneous and mucocutaneous leishmaniasis
| Keyword | Reasoning |
| --- | --- |
| Skin sores | Symptom of cutaneous leishmaniasis |
| Espundia | South American term for mucocutaneous leishmaniasis |

A.4.4 African trypanosomiasis
| Keyword | Reasoning |
| --- | --- |
| African trypanosomiasis | Parasitic disease transmitted by tsetse flies |
| Sleeping sickness | Common name for African trypanosomiasis |
| Tsetse fly disease | Alternative name for African trypanosomiasis |
| Trypanosoma brucei | Parasite causing African trypanosomiasis |

A.4.5 Schistosomiasis
| Keyword | Reasoning |
| --- | --- |
| Schistosomiasis | Parasitic disease caused by flatworms |
| Bilharzia | Alternative name for schistosomiasis |
| Snail fever | Common name for schistosomiasis |
| Schistosoma | Genus of parasites causing schistosomiasis |

A.4.6 Cysticercosis
| Keyword | Reasoning |
| --- | --- |
| Cysticercosis | Tissue infection caused by tapeworm larvae |
| Neurocysticercosis | Cysticercosis affecting the brain |
| Taenia solium | Tapeworm species causing cysticercosis |

A.4.7 Cystic echinococcosis
| Keyword | Reasoning |
| --- | --- |
| Echinococcosis | Parasitic disease caused by tapeworm larvae |
| Hydatid disease | Alternative name for echinococcosis |

A.4.8 Lymphatic filariasis
| Keyword | Reasoning |
| --- | --- |
| Lymphatic filariasis | Parasitic disease affecting the lymphatic system |
| Wuchereria bancrofti | Main parasite causing lymphatic filariasis |
| Brugia malayi | Another parasite causing lymphatic filariasis |

A.4.9 Onchocerciasis
| Keyword | Reasoning |
| --- | --- |
| Onchocerciasis | Parasitic disease causing skin and eye problems |
| River blindness | Common name for onchocerciasis |
| Onchocerca volvulus | Parasite causing onchocerciasis |

A.4.10 Trachoma
| Keyword | Reasoning |
| --- | --- |
| Trachoma | Bacterial eye infection causing blindness |
| Chlamydia trachomatis | Bacterium causing trachoma |
| Trichiasis | Complication of trachoma affecting eyelashes |

A.4.11 Dengue
| Keyword | Reasoning |
| --- | --- |
| Dengue | Mosquito-borne viral disease |
| Aedes mosquito | Vector for dengue virus |

A.4.12 Yellow fever
| Keyword | Reasoning |
| --- | --- |
| Yellow fever | Mosquito-borne viral disease |
| YF vaccine | Vaccine to prevent yellow fever |
| Aedes aegypti | Mosquito species transmitting yellow fever |

A.4.13 Rabies
| Keyword | Reasoning |
| --- | --- |
| Rabies | Viral disease affecting the nervous system |
| Post-exposure prophylaxis | Preventive treatment after potential rabies exposure |
| Lyssavirus | Genus of viruses causing rabies |

A.4.14 Intestinal nematode infections
| Keyword | Reasoning |
| --- | --- |
| Intestinal nematode infections | Infections caused by intestinal worms |
| Soil-transmitted helminths | Common name for intestinal worms |
| Intestinal worms | General term for parasitic worms in the intestines |

A.4.14.1 Ascariasis
| Keyword | Reasoning |
| --- | --- |
| Ascariasis | Intestinal infection caused by Ascaris roundworms |
| Roundworm | Common name for Ascaris worms |
| Ascaris lumbricoides | Scientific name of the roundworm causing ascariasis |

A.4.14.2 Trichuriasis
| Keyword | Reasoning |
| --- | --- |
| Trichuriasis | Intestinal infection caused by whipworms |
| Whipworm | Common name for Trichuris worms |
| Trichuris trichiura | Scientific name of the whipworm causing trichuriasis |

A.4.14.3 Hookworm disease
| Keyword | Reasoning |
| --- | --- |
| Hookworm disease | Intestinal infection caused by hookworms |
| Ancylostomiasis | Alternative name for hookworm disease |
| Necator americanus | Species of hookworm causing infection |
| Ancylostoma duodenale | Another species of hookworm causing infection |

A.4.15 Food-borne trematodiasis
| Keyword | Reasoning |
| --- | --- |
| Food-borne trematodiasis | Parasitic infections transmitted through food |
| Liver flukes | Parasites affecting the liver |
| Lung flukes | Parasites affecting the lungs |
| Intestinal flukes | Parasites affecting the intestines |

A.4.16 Leprosy
| Keyword | Reasoning |
| --- | --- |
| Leprosy | Chronic bacterial infection affecting skin and nerves |
| Hansen's disease | Alternative name for leprosy |
| Mycobacterium leprae | Bacterium causing leprosy |
| Hansens disease | Alternative spelling of Hansen's disease |

A.4.17 Ebola
| Keyword | Reasoning |
| --- | --- |
| Ebola | Severe viral hemorrhagic fever |
| Filovirus | Family of viruses including Ebola |

A.4.18 Zika virus
| Keyword | Reasoning |
| --- | --- |
| Zika virus | Mosquito-borne virus causing birth defects |
| Zika | Short form of Zika virus |

A.4.19 Guinea worm disease
| Keyword | Reasoning |
| --- | --- |
| Guinea worm disease | Parasitic infection caused by Dracunculus worm |
| Guinea worm | Common name for the parasite causing the disease |
| Dracunculiasis | Scientific name for Guinea worm disease |
| Dracunculus medinensis | Scientific name of the Guinea worm |

A.4.20 Other neglected tropical diseases
| Keyword | Reasoning |
| --- | --- |
| Buruli ulcer | Bacterial infection causing skin ulcers |
| Yaws | Bacterial infection affecting skin bones and cartilage |
| Mycetoma | Chronic skin and soft tissue infection |
| Chromoblastomycosis | Fungal skin infection |
| Deep mycoses | Fungal infections affecting deep tissues |
| A.6.1 Maternal disorders |  |
| Maternal disorder | Health problems related to pregnancy and childbirth |
| Maternal disease | Diseases affecting pregnant women |
| Maternal infections | Infections affecting pregnant women |
| Miscarriage | Spontaneous loss of pregnancy before 20 weeks |
| Ectopic pregnancy | Pregnancy outside the uterus |
| Maternal deaths | Deaths related to pregnancy or childbirth |
| Eclampsia | Seizures during pregnancy due to high blood pressure |
| Pre-eclampsia | High blood pressure during pregnancy |

A.6.1.1 Maternal hemorrhage
| Keyword | Reasoning |
| --- | --- |
| Maternal hemorrhage | Excessive bleeding during pregnancy or childbirth |

A.6.1.2 Maternal sepsis and other maternal infections
| Keyword | Reasoning |
| --- | --- |
| Maternal sepsis | Infection during pregnancy or after childbirth |

A.6.1.4 Maternal obstructed labor and uterine rupture
| Keyword | Reasoning |
| --- | --- |
| Maternal obstructed labor | Difficulty in fetus passing through the birth canal |
| Maternal uterine rupture | Tearing of the uterus during pregnancy or childbirth |

A.6.1.5 Maternal abortion and miscarriage
| Keyword | Reasoning |
| --- | --- |
| Maternal abortion | Termination of pregnancy |

A.6.2 Neonatal disorders
| Keyword | Reasoning |
| --- | --- |
| Neonatal disorder | Health problems affecting newborns |
| Neonatal disease | Diseases affecting newborns |
| Birth asphyxia | Lack of oxygen to a baby during birth |
| Stillbirth | Baby born without signs of life at or after 28 weeks |
| Neonatal encephalopathy | Brain disorder in newborns |
| Neonatal infections | Infections affecting newborns |

A.6.2.1 Neonatal preterm birth
| Keyword | Reasoning |
| --- | --- |
| Preterm | Born before 37 weeks of pregnancy |
| Neonatal preterm birth | Birth before 37 weeks of pregnancy |

A.6.2.3 Neonatal sepsis and other neonatal infections
| Keyword | Reasoning |
| --- | --- |
| Neonatal sepsis | Blood infection in newborns |
| Neonatal jaundice | Yellowing of a newborn's skin and eyes |

A.6.2.4 Hemolytic disease and other neonatal jaundice
| Keyword | Reasoning |
| --- | --- |
| Hemolytic disease | Blood disorder in newborns |

Nutritional Deficiencies A.7 Nutritional deficiencies
| Keyword | Reasoning |
| --- | --- |
| Malnutrition | Lack of proper nutrition |
| Undernutrition | Insufficient intake of nutrients |
| Stunting | Impaired growth and development due to poor nutrition |
| Nutritional deficiency | Lack of essential nutrients |
| Nutritional deficiencies | Multiple lacks of essential nutrients |

A.7.1 Protein-energy malnutrition
| Keyword | Reasoning |
| --- | --- |
| Protein-energy deficiency | Lack of protein and energy in the diet |
| Protein-energy deficiencies | Multiple lacks of protein and energy in the diet |
| Wasting | Severe weight loss due to malnutrition |

A.7.2 Iodine deficiency
| Keyword | Reasoning |
| --- | --- |
| Iodine deficiency | Lack of iodine in the diet |
| Iodine deficiencies | Multiple instances of iodine lack in the diet |

A.7.3 Vitamin A deficiency
| Keyword | Reasoning |
| --- | --- |
| Vitamin A deficiencies | Multiple instances of vitamin A lack in the diet |
| Vitamin A deficiency | Lack of vitamin A in the diet |

A.7.4 Dietary iron deficiency
| Keyword | Reasoning |
| --- | --- |
| Iron deficiencies | Multiple instances of iron lack in the diet |
| Iron deficiency | Lack of iron in the diet |
| Anemia | Condition often caused by iron deficiency |

A.7.5 Other nutritional deficiencies
| Keyword | Reasoning |
| --- | --- |
| Vitamin deficiency | Lack of essential vitamins in the diet |
| Zinc deficiency | Lack of zinc in the diet |
| Folate deficiency | Lack of folate (vitamin B9) in the diet |

Neoplasms B.1 Neoplasms
| Keyword | Reasoning |
| --- | --- |
| Neoplasm | Abnormal growth of tissue (tumor) |
| Tumor | Abnormal mass of tissue |
| Malignant | Cancerous |
| Carcinoma | Cancer originating in epithelial cells |
| Sarcoma | Cancer originating in connective tissue |
| Leukemia | Cancer of blood-forming tissues |
| Lymphoma | Cancer of lymphatic system |
| Melanoma | Cancer originating in melanocytes |
| Metastasis | Spread of cancer from one part of the body to another |
| Oncology | Study and treatment of cancer |
| Cancer | General term for malignant neoplasms |

B.1.7.5 Hepatoblastoma
| Keyword | Reasoning |
| --- | --- |
| Hepatoblastoma | Rare liver cancer typically occurring in young children |

B.1.25.1 Retinoblastoma
| Keyword | Reasoning |
| --- | --- |
| Retinoblastoma | Cancer of the retina typically occurring in young children |

B.1.28 Mesothelioma
| Keyword | Reasoning |
| --- | --- |
| Mesothelioma | Cancer affecting the lining of the lungs and abdomen |

B.1.31 Multiple myeloma
| Keyword | Reasoning |
| --- | --- |
| Multiple myeloma | Cancer of plasma cells |

Cardiovascular diseases B.2 Cardiovascular diseases
| Keyword | Reasoning |
| --- | --- |
| Cardiovascular disease | Diseases affecting the heart and blood vessels |
| Heart disease | Diseases affecting the heart |
| Hypertension | High blood pressure |
| High blood pressure | Another term for hypertension |
| Coronary artery disease | Narrowing of the coronary arteries |
| Atherosclerosis | Buildup of plaque in the arteries |
| Arrhythmia | Irregular heartbeat |
| Atrial fibrillation | Common type of arrhythmia |
| Heart failure | Heart's inability to pump blood effectively |
| Myocardial infarction | Heart attack |
| Aneurysm | Bulge in a blood vessel wall |
| Peripheral artery disease | Narrowing of arteries in the limbs |
| Thrombosis | Blood clot formation |
| B.2.3 Stroke |  |
| Keyword | Reasoning |
| Stroke | Sudden interruption of blood supply to the brain |
| B.2.3.2 Intracerebral hemorrhage |  |
| Keyword | Reasoning |
| Intracerebral hemorrhage | Bleeding within the brain tissue |
| B.2.3.3 Subarachnoid hemorrhage |  |
| Keyword | Reasoning |
| Subarachnoid hemorrhage | Bleeding in the space surrounding the brain |
| B.2.5 Non-rheumatic valvular heart disease |  |
| Keyword | Reasoning |
| Valve disease | Diseases affecting heart valves |
| B.2.6 Cardiomyopathy and myocarditis |  |
| Keyword | Reasoning |
| Cardiomyopathy | Diseases of the heart muscle |
| B.2.6.1 Myocarditis |  |
| Keyword | Reasoning |
| Myocarditis | Inflammation of the heart muscle |
| B.2.8 Atrial fibrillation and flutter |  |
| Keyword | Reasoning |
| Atrial fibrillation | Common type of irregular heartbeat |
| B.2.9 Aortic aneurysm |  |
| Keyword | Reasoning |
| Aortic aneurysm | Bulge in the wall of the aorta |
| B.2.10 Lower extremity peripheral arterial disease |  |
| Keyword | Reasoning |
| Peripheral arterial disease | Narrowing of arteries in the legs |
| B.2.11 Endocarditis |  |
| Keyword | Reasoning |
| Endocarditis | Inflammation of the inner lining of the heart |
| B.2.12 Other cardiovascular and circulatory diseases |  |
| Keyword | Reasoning |
| Circulatory disease | Diseases affecting blood circulation |
| B.3 Chronic respiratory diseases |  |
| Keyword | Reasoning |
| Chronic respiratory disease | Long-term diseases affecting the airways and lungs |

B.3.1 Chronic obstructive pulmonary disease
| Keyword | Reasoning |
| --- | --- |
| COPD | Acronym for Chronic Obstructive Pulmonary Disease |
| Chronic obstructive pulmonary disease | Group of lung diseases causing airflow blockage |

B.3.2 Pneumoconiosis
| Keyword | Reasoning |
| --- | --- |
| Pneumoconiosis | Lung diseases caused by inhalation of dust |

B.3.2.1 Silicosis
| Keyword | Reasoning |
| --- | --- |
| Silicosis | Lung disease caused by inhalation of silica dust |

B.3.2.2 Asbestosis
| Keyword | Reasoning |
| --- | --- |
| Asbestosis | Lung disease caused by inhalation of asbestos fibers |

B.3.3 Asthma
| Keyword | Reasoning |
| --- | --- |
| Asthma | Chronic disease causing airway inflammation and narrowing |

B.3.4 Interstitial lung disease and pulmonary sarcoidosis
| Keyword | Reasoning |
| --- | --- |
| Pulmonary fibrosis | Scarring of lung tissue |
| Sarcoidosis | Inflammatory disease affecting multiple organs including lungs |

B.3.5 Other chronic respiratory diseases
| Keyword | Reasoning |
| --- | --- |
| Emphysema | Damage to the air sacs in the lungs |
| Cystic fibrosis | Inherited disorder affecting lungs and other organs |
| Bronchitis | Inflammation of the bronchial tubes |

Digestive diseases B.4 Digestive diseases
| Keyword | Reasoning |
| --- | --- |
| Digestive disease | Diseases affecting the digestive system |
| Digestive system disease | Another term for digestive diseases |

B.4.1 Cirrhosis and other chronic liver diseases
| Keyword | Reasoning |
| --- | --- |
| Cirrhosis | Scarring of the liver |
| Chronic liver disease | Long-term diseases affecting the liver |
| Fatty liver disease | Accumulation of fat in the liver |

B.4.2.1 Peptic ulcer disease
| Keyword | Reasoning |
| --- | --- |
| Peptic ulcer disease | Sores in the lining of stomach or small intestine |

B.4.2.2 Gastritis and duodenitis
| Keyword | Reasoning |
| --- | --- |
| Gastritis | Inflammation of the stomach lining |
| Duodenitis | Inflammation of the duodenum |

B.4.2.3 Gastroesophageal reflux disease
| Keyword | Reasoning |
| --- | --- |
| GERD | Acronym for Gastroesophageal Reflux Disease |
| Gastroesophageal reflux disease | Chronic digestive disease causing acid reflux |
| B.4.3 Appendicitis |  |
| Keyword | Reasoning |
| Appendicitis | Inflammation of the appendix |
| B.4.4 Paralytic ileus and intestinal obstruction |  |
| Keyword | Reasoning |
| Paralytic ileus | Intestinal paralysis |
| Intestinal obstruction | Blockage of the intestines |
| B.4.5 Inguinal, femoral, and abdominal hernia |  |
| Keyword | Reasoning |
| Inguinal hernia | Protrusion of tissue through the groin |
| Femoral hernia | Protrusion of tissue through the upper thigh |
| Abdominal hernia | Protrusion of tissue through the abdominal wall |
| B.4.6 Inflammatory bowel disease |  |
| Keyword | Reasoning |
| Inflammatory bowel disease | Chronic inflammation of the digestive tract |
| Crohn's disease | Type of inflammatory bowel disease |
| Ulcerative colitis | Type of inflammatory bowel disease |
| B.4.7 Vascular intestinal disorders |  |
| Keyword | Reasoning |
| Vascular intestinal disorder | Disorders affecting blood vessels in the intestines |
| B.4.8 Gallbladder and biliary diseases |  |
| Keyword | Reasoning |
| Gallbladder | Organ storing and concentrating bile |
| Biliary disease | Diseases affecting the biliary system |
| B.4.9 Pancreatitis |  |
| Keyword | Reasoning |
| Pancreatitis | Inflammation of the pancreas |
| B.4.10 Other digestive diseases |  |
| Keyword | Reasoning |
| Gallstone | Solid material that forms in the gallbladder |
| Irritable bowel syndrome | Disorder affecting the large intestine |
| Bowel disease | Diseases affecting the bowels |
| Intestinal disorder | Disorders affecting the intestines |
| Intestinal disease | Diseases affecting the intestines |
| B.5 Neurological disorders |  |
| Keyword | Reasoning |
| Neurological disorder | Disorders affecting the nervous system |
| B.5.1 Alzheimer's disease and other dementias |  |
| Keyword | Reasoning |
| Alzheimer | Common form of dementia |
| Dementia | Decline in cognitive function |
| B.5.2 Parkinson's disease |  |
| Keyword | Reasoning |
| Parkinson | Progressive nervous system disorder affecting movement |
| B.5.3 Idiopathic epilepsy |  |
| Keyword | Reasoning |
| Epilepsy | Neurological disorder causing seizures |
| Seizure | Sudden uncontrolled electrical disturbance in the brain |
| B.5.4 Multiple sclerosis |  |
| Keyword | Reasoning |
| Multiple sclerosis | Disease affecting the central nervous system |
| B.5.5 Motor neuron disease |  |
| Keyword | Reasoning |
| Motor neuron disease | Progressive neurological disease |
| ALS | Amyotrophic Lateral Sclerosis a type of motor neuron disease |
| B.5.6 Headache disorders |  |
| Keyword | Reasoning |
| Headache disorder | Disorders characterized by recurrent headaches |
| B.5.6.1 Migraine |  |
| Keyword | Reasoning |
| Migraine | Severe recurring headache |
| B.5.6.2 Tension-type headache |  |
| Keyword | Reasoning |
| Tension-type headache | Common form of primary headache |
| B.5.7 Other neurological disorders |  |
| Keyword | Reasoning |
| Neuropathy | Damage or dysfunction of nerves |
| Cerebral palsy | Group of disorders affecting movement and posture |
| Huntington's disease | Inherited disorder causing progressive brain damage |
| B.6 Mental disorders |  |
| Keyword | Reasoning |
| Mental disorder | Disorders affecting behavior and thinking |
| B.6.1 Schizophrenia |  |
| Keyword | Reasoning |
| Schizophrenia | Severe mental disorder affecting thinking and behavior |
| B.6.2 Depressive disorders |  |
| Keyword | Reasoning |
| Depressive disorder | Mood disorders characterized by persistent feelings of sadness |
| Depression | Mood disorder characterized by persistent sadness |
| B.6.2.2 Dysthymia |  |
| Keyword | Reasoning |
| Dysthymia | Persistent mild depression |
| B.6.3 Bipolar disorder |  |
| Keyword | Reasoning |
| Bipolar | Mood disorder with alternating episodes of mania and depression |
| B.6.4 Anxiety disorders |  |
| Keyword | Reasoning |
| Anxiety | Disorders characterized by excessive worry or fear |
| B.6.5 Eating disorders |  |
| Keyword | Reasoning |
| Eating disorder | Disorders characterized by abnormal eating habits |
| B.6.5.1 Anorexia nervosa |  |
| Keyword | Reasoning |
| Anorexia | Eating disorder characterized by low weight and food restriction |
| B.6.5.2 Bulimia nervosa |  |
| Keyword | Reasoning |
| Bulimia | Eating disorder characterized by binge eating and purging |
| B.6.6 Autism spectrum disorders |  |
| Keyword | Reasoning |
| Autism | Developmental disorder affecting communication and behavior |
| B.6.7 Attention-deficit/hyperactivity disorder |  |
| Keyword | Reasoning |
| Attention-deficit/hyperactivity disorder | Neurodevelopmental disorder affecting attention and behavior |
| ADHD | Acronym for Attention-Deficit/Hyperactivity Disorder |
| B.6.8 Conduct disorder |  |
| Keyword | Reasoning |
| Conduct disorder | Behavioral disorder in children and adolescents |
| B.6.10 Other mental disorders |  |
| Keyword | Reasoning |
| PTSD | Post-Traumatic Stress Disorder |
| OCD | Obsessive-Compulsive Disorder |
| Personality disorder | Disorders characterized by unhealthy patterns of thinking and behavior |
| Post-traumatic stress | Disorder that may develop after experiencing a traumatic event |
| B.7 Substance use disorders |  |
| Keyword | Reasoning |
| Substance use disorder | Disorders related to the use of psychoactive substances |
| Substance abuse | Harmful use of psychoactive substances |
| Substance dependence | Physical or psychological dependence on a substance |
| B.7.1 Alcohol use disorders |  |
| Keyword | Reasoning |
| Alcohol use disorder | Disorder characterized by problematic patterns of alcohol use |
| Alcohol abuse | Harmful or hazardous use of alcohol |
| Alcoholism | Addiction to alcohol |
| B.7.2 Drug use disorders |  |
| Keyword | Reasoning |
| Drug use disorder | Disorder characterized by problematic patterns of drug use |
| Drug abuse | Harmful or hazardous use of drugs |

B.7.2.1 Opioid use disorders
| Keyword | Reasoning |
| --- | --- |
| Opioid use disorder | Disorder related to the use of opioids |
| Opioid abuse | Harmful or hazardous use of opioids |

B.7.2.2 Cocaine use disorders
| Keyword | Reasoning |
| --- | --- |
| Cocaine use disorder | Disorder related to the use of cocaine |
| Cocaine abuse | Harmful or hazardous use of cocaine |

B.7.2.3 Amphetamine use disorders
| Keyword | Reasoning |
| --- | --- |
| Amphetamine use disorder | Disorder related to the use of amphetamines |
| Amphetamine abuse | Harmful or hazardous use of amphetamines |

B.7.2.4 Cannabis use disorders
| Keyword | Reasoning |
| --- | --- |
| Cannabis use disorder | Disorder related to the use of cannabis |
| Cannabis abuse | Harmful or hazardous use of cannabis |

B.7.2.5 Other drug use disorders
| Keyword | Reasoning |
| --- | --- |
| Substance misuse | Inappropriate use of psychoactive substances |

Diabetes and kidney diseases B.8.1 Diabetes mellitus
| Keyword | Reasoning |
| --- | --- |
| Diabetes | Group of metabolic disorders characterized by high blood sugar |
| Diabetic | Relating to diabetes |
| Insulin resistance | Condition in which cells don't respond normally to insulin |

B.8.1.1 Diabetes mellitus type 1
| Keyword | Reasoning |
| --- | --- |
| Diabetes mellitus type 1 | Autoimmune form of diabetes |

B.8.1.2 Diabetes mellitus type 2
| Keyword | Reasoning |
| --- | --- |
| Diabetes mellitus type 2 | Most common form of diabetes often related to lifestyle |

B.8.2 Chronic kidney disease
| Keyword | Reasoning |
| --- | --- |
| Kidney disease | Long-term condition where kidneys don't work properly |
| Nephropathy | Damage to or disease of the kidneys |
| Dialysis | Treatment for kidney failure |
| Renal failure | Condition where kidneys fail to function properly |
| Chronic kidney disease | Long-term condition of kidney damage or dysfunction |

B.8.2.4 Chronic kidney disease due to glomerulonephritis
| Keyword | Reasoning |
| --- | --- |
| Glomerulonephritis | Inflammation of the kidney's filtering units |

B.8.2.5 Chronic kidney disease due to other and unspecified causes
| Keyword | Reasoning |
| --- | --- |
| End-stage renal disease | Final stage of chronic kidney disease |

B.8.3 Acute glomerulonephritis
| Keyword | Reasoning |
| --- | --- |
| Acute kidney injury | Sudden decrease in kidney function |
| B.9 Skin and subcutaneous diseases |  |
| Keyword | Reasoning |
| Skin disease | Diseases affecting the skin |
| Subcutaneous disease | Diseases affecting the tissue beneath the skin |
| B.9.1 Dermatitis |  |
| Keyword | Reasoning |
| Dermatitis | Inflammation of the skin |
| B.9.1.1 Atopic dermatitis |  |
| Keyword | Reasoning |
| Eczema | Inflammatory skin condition causing itchy rashes |
| B.9.2 Psoriasis |  |
| Keyword | Reasoning |
| Psoriasis | Chronic autoimmune condition causing rapid skin cell growth |
| B.9.3 Bacterial skin diseases |  |
| Keyword | Reasoning |
| Bacterial skin disease | Skin infections caused by bacteria |
| B.9.3.1 Cellulitis |  |
| Keyword | Reasoning |
| Cellulitis | Bacterial infection of the deep layers of skin |
| B.9.3.2 Pyoderma |  |
| Keyword | Reasoning |
| Pyoderma | Bacterial skin infection characterized by pus-filled lesions |
| B.9.4 Scabies |  |
| Keyword | Reasoning |
| Scabies | Skin infestation caused by tiny mites |
| B.9.7 Acne vulgaris |  |
| Keyword | Reasoning |
| Acne | Common skin condition causing pimples and other lesions |
| Acne vulgaris | Medical term for common acne |
| B.9.8 Alopecia areata |  |
| Keyword | Reasoning |
| Alopecia | General term for hair loss |
| Alopecia areata | Autoimmune disorder causing hair loss in patches |
| B.9.9 Pruritus |  |
| Keyword | Reasoning |
| Pruritus | Itching sensation |
| B.9.10 Urticaria |  |
| Keyword | Reasoning |
| Urticaria | Hives condition causing itchy welts |
| B.9.11 Decubitus ulcer |  |
| Keyword | Reasoning |
| Decubitus ulcer | Pressure sores or bedsores |
| B.9.12 Other skin and subcutaneous diseases |  |
| Keyword | Reasoning |
| Rosacea | Chronic inflammatory skin condition |
| Vitiligo | Condition causing loss of skin pigmentation |
| B.10 Sense organ diseases |  |
| Keyword | Reasoning |
| Sense organ disease | Diseases affecting organs of sight hearing smell taste or touch |
| B.10.1 Blindness and vision loss |  |
| Keyword | Reasoning |
| Blindness | Complete or partial loss of sight |
| Vision loss | Partial or complete loss of vision |
| Vision impairment | Decreased ability to see |
| Visual impairment | Another term for vision impairment |
| B.10.1.1 Glaucoma |  |
| Keyword | Reasoning |
| Glaucoma | Group of eye conditions damaging the optic nerve |
| B.10.1.2 Cataract |  |
| Keyword | Reasoning |
| Cataract | Clouding of the eye's natural lens |
| B.10.1.3 Age-related macular degeneration |  |
| Keyword | Reasoning |
| Macular degeneration | Eye disorder causing loss of central vision |
| B.10.1.4 Refraction disorders |  |
| Keyword | Reasoning |
| Refraction disorder | Vision problems due to how the eye focuses light |
| Refractive error | Another term for refraction disorder |
| B.10.1.6 Other vision loss |  |
| Keyword | Reasoning |
| Retinopathy | Damage to the retina of the eye |
| Amblyopia | Decreased vision in one eye also known as lazy eye |
| B.10.2 Age-related and other hearing loss |  |
| Keyword | Reasoning |
| Hearing loss | Partial or complete inability to hear |
| Tinnitus | Perception of noise or ringing in the ears |
| Deafness | Severe or complete hearing loss |
| B.10.3 Other sense organ diseases |  |
| Keyword | Reasoning |
| Anosmia | Loss of sense of smell |
| Ageusia | Loss of sense of taste |
| Conjunctivitis | Inflammation of the conjunctiva of the eye |
| Vestibular disorder | Disorders affecting balance and spatial orientation |

Musculoskeletal disorders B.11 Musculoskeletal disorders
| Keyword | Reasoning |
| --- | --- |
| Musculoskeletal disorder | Disorders affecting muscles bones and joints |
| Musculoskeletal disease | Diseases affecting muscles bones and joints |

B.11.1 Rheumatoid arthritis
| Keyword | Reasoning |
| --- | --- |
| Rheumatoid arthritis | Chronic inflammatory disorder affecting joints |

B.11.2 Osteoarthritis
| Keyword | Reasoning |
| --- | --- |
| Osteoarthritis | Degenerative joint disease |
| Arthritis | General term for joint inflammation |

B.11.3 Low back pain
| Keyword | Reasoning |
| --- | --- |
| Low back pain | Pain in the lower back |

B.11.4 Neck pain
| Keyword | Reasoning |
| --- | --- |
| Neck pain | Pain in the neck area |

B.11.5 Gout
| Keyword | Reasoning |
| --- | --- |
| Gout | Form of inflammatory arthritis caused by uric acid crystals |

B.11.6 Other musculoskeletal disorders
| Keyword | Reasoning |
| --- | --- |
| Tendinitis | Inflammation of a tendon |
| Scoliosis | Abnormal curvature of the spine |
| Carpal tunnel syndrome | Compression of the median nerve in the wrist |
| Osteoporosis | Condition causing bones to become weak and brittle |
| Fibromyalgia | Chronic condition causing widespread muscle pain and tenderness |
| Bursitis | Inflammation of the fluid-filled sacs that cushion joints |
| Spinal stenosis | Narrowing of spaces in the spine |
| Rotator cuff disorder | Injuries and conditions affecting the shoulder joint |
| Muscular dystrophy | Group of genetic diseases causing progressive muscle weakness |
| Bone fractures | Breaks in bone continuity |

## References

[1] World Health Organization. World health statistics 2023: Monitoring health for the SDGs. https://cdn.who.int/media/docs/default-source/gho-documents/world-health-statistic-reports/2023/world-health-statistics-2023_20230519_pdf (2023).

[2] United Nations. The Sustainable Development Goals Report 2024. United Nations Sustainable Development Goals (2024).

[3] Comfort, H. et al. Global, regional, and national stillbirths at 20 weeks’ gestation or longer in 204 countries and territories, 1990–2021: findings from the Global Burden of Disease Study 2021. The Lancet 404, 1955–1988 (2024).

[4] Carter, A. et al. Global, regional, and national burden of HIV/AIDS, 1990–2021, and forecasts to 2050, for 204 countries and territories: the Global Burden of Disease Study 2021. The Lancet HIV 11, E807–E822 (2024).

[5] Vollset, S. E. et al. Burden of disease scenarios for 204 countries and territories, 2022–2050: a forecasting analysis for the Global Burden of Disease Study 2021. The Lancet 403, 2204–2256 (2024).

[6] World Health Organization. Countdown to 2030: Stronger alignment for country impact. https://www.who.int/news/item/23-05-2024-countdown-to-2030--stronger-alignment-for-country-impact (2024).

[7] United Nations. Transforming our world: the 2030 Agenda for Sustainable Development. United Nations General Assembly (2015).

[8] Hafeez, A. et al. The state of health in Pakistan and its provinces and territories, 1990–2019: a systematic analysis for the Global Burden of Disease Study 2019. The Lancet Global Health 11, e229–e243 (2023).

[9] Li, Z. et al. Temporal trends in the burden of non-communicable diseases in countries with the highest malaria burden, 1990–2019: Evaluating the double burden of non-communicable and communicable diseases in epidemiological transition. Globalization and Health 18 (2022).

[10] Joint United Nations Programme on HIV/AIDS (UNAIDS). Global HIV & AIDS statistics - 2024 fact sheet. Web page (2024). URL https://www.unaids.org/en/resources/fact-sheet. Accessed June 1, 2025.

[11] World Health Organization. Global tuberculosis report 2024. Tech. Rep., World Health Organization, Geneva (2024).

[12] World Health Organization. World malaria report 2023. Tech. Rep., World Health Organization, Geneva (2023).

[13] Callaway, E. ‘It is chaos’: US funding freezes are endangering global health. Nature 638, 299–300 (2025).

[14] Sheldrick, M. Foreign aid is shrinking—What happens next? (2025). URL https://www.forbes.com/sites/globalcitizen/2025/02/25/foreign-aid-is-shrinking-what-happens-next/.

[15] Yamey, G. & Titanji, B. K. Withdrawal of the United States from the WHO — How President Trump is weakening public health. New England Journal of Medicine 392, 1457–1460 (2025).

[16] Bishen, S. Funding gaps threaten global health security, warns OECD – and other top health stories. World Economic Forum (2025). Accessed June 1, 2025.

[17] Weaver, M. R. et al. Health care spending effectiveness: Estimates suggest that spending improved US health from 1996 to 2016. Health Affairs 41, 994–1004 (2022).

[18] Shi, J., Jin, Y. & Zheng, Z. Addressing global health challenges requires harmonised and innovative approaches to the development assistance for health. BMJ Global Health 8, e012314 (2023).

[19] Bendavid, E. & Bhattacharya, J. The relationship of health aid to population health improvements. JAMA Internal Medicine 174, 881–887 (2014).

[20] Xie, S. et al. Evolution and effectiveness of bilateral and multilateral development assistance for health: a mixed-methods review of trends and strategic shifts (1990–2022). BMJ Global Health 10, e017818 (2025).

[21] Kumar, M. B., et al. Donor aid mentioning newborns and stillbirths, 2002–19: an analysis of levels, trends, and equity. The Lancet Global Health 11, e1785–e1793 (2023). Publisher: Elsevier.

[22] Fullman, N. et al. Measuring performance on the healthcare access and quality index for 195 countries and territories and selected subnational locations: a systematic analysis from the Global Burden of Disease Study 2016. The Lancet 391, 2236–2271 (2018).

[23] Dieleman, J. L. et al. Global health development assistance remained steady in 2013 but did not align with recipients’ disease burden. Health Affairs 33, 878–886 (2014).

[24] Micah, A. E. et al. Tracking development assistance for health and for COVID-19: a review of development assistance, government, out-of-pocket, and other private spending on health for 204 countries and territories, 1990–2050. The Lancet 398, 1317–1343 (2021).

[25] Chang, A. Y. et al. Past, present, and future of global health financing: a review of development assistance, government, out-of-pocket, and other private spending on health for 195 countries, 1995–2050. The Lancet 393, 2233–2260 (2019).

[26] Li, Z., Richter, L. & Lu, C. Tracking development assistance for reproductive, maternal, newborn, child and adolescent health in conflict-affected countries. BMJ Global Health 4, e001614 (2019).

[27] Dieleman, J. L. et al. Development assistance for health: past trends, associations, and the future of international financial flows for health. The Lancet 387, 2536–2544 (2016).

[28] Dieleman, J. L. et al. National spending on health by source for 184 countries between 2013 and 2040. The Lancet 387, 2521–2535 (2016).

[29] Boum, Y., II et al. Advancing equitable global health research partnerships in Africa. BMJ Global Health 3, e000868 (2018).

[30] Resch, S., Ryckman, T. & Hecht, R. Funding AIDS programmes in the era of shared responsibility: an analysis of domestic spending in 12 low-income and middle-income countries. The Lancet Global Health 3, e52–e61 (2014).

[31] Institute for Health Metrics and Evaluation. Financing global health (2025). URL https://www.healthdata.org/data-tools-practices/interactive-visuals/financing-global-health. Accessed November 5, 2025.

[32] Grollman, C. et al. 11 years of tracking aid to reproductive, maternal, newborn, and child health: estimates and analysis for 2003–13 from the Countdown to 2015. The Lancet Global Health 5 (2016).

[33] Mann, C. et al. Countdown to 2015 country case studies: what can analysis of national health financing contribute to understanding MDG 4 and 5 progress? BMC Public Health 16 (2016).

[34] Lu, C. et al. Assessing development assistance for child survival between 2000 and 2014: A multi-sectoral perspective. PLoS ONE 12, e0178887 (2017).

[35] Martinez-Alvarez, M. et al. Trends in the alignment and harmonization of reproductive, maternal, newborn, and child health funding, 2008–13. Health Affairs 36, 1876–1886 (2017).

[36] Grollman, C. et al. Developing a dataset to track aid for reproductive, maternal, newborn and child health, 2003–2013. Scientific Data 4 (2017).

[37] Pitt, C., Grollman, C., Martinez-Alvarez, M., Arregoces, L. & Borghi, J. Tracking aid for global health goals: a systematic comparison of four approaches applied to reproductive, maternal, newborn, and child health. The Lancet Global Health 6, e859–e874 (2018).

[38] Micah, A. E. et al. Health sector spending and spending on HIV/AIDS, tuberculosis, and malaria, and development assistance for health: progress towards Sustainable Development Goal 3. The Lancet 396, 693–724 (2020).

[39] Pitt, C. et al. Countdown to 2015: an analysis of donor funding for prenatal and neonatal health, 2003–2013. BMJ Global Health 2, e000205 (2017).

[40] James, S. L. et al. Global, regional, and national incidence, prevalence, and years lived with disability for 354 diseases and injuries for 195 countries and territories, 1990–2017: a systematic analysis for the Global Burden of Disease Study 2017. The Lancet 392, 1789–1858 (2018).

[41] Murray, C. J. L. The Global Burden of Disease Study at 30 years. Nature Medicine 28, 2019–2026 (2022).

[42] Voigt, K. & King, N. B. Out of Alignment? Limitations of the Global Burden of Disease in Assessing the Allocation of Global Health Aid. Public Health Ethics 10, 244–256 (2017).

[43] Anand, S. & Hanson, K. Disability-adjusted life years: a critical review. Journal of Health Economics 16, 685–702 (1997).

[44] OECD Statistics. Creditor reporting system. URL https://data-explorer.oecd.org/vis?tm=crs&pg=0&snb=25&df[ds]=dsDisseminateFinalDMZ&df[id]=DSD_CRS%40DF_CRS&df[ag]=OECD.DCD.FSD&df[vs]=1.4&dq=DAC..1000.100._T._T.D.Q._T..&lom=LASTNPERIODS&lo=5&to[TIME_PERIOD]=false. Accessed June 1, 2025.

[45] The Global Fund. Allocation methodology 2017–2019 (2016). URL https://archive.theglobalfund.org/media/4224/archive_bm35-05-allocationmethodology2017-2019_report_en.pdf.

[46] The Global Fund. Overview of the allocation methodology. URL https://archive.theglobalfund.org/media/1380/archive_allocations-2014-2016_methodology_en.pdf.

[47] Global Fund. Audit report the global fund’s methodology for the allocation of funds internal controls, risk management, and governance processes. URL https://www.theglobalfund.org/media/2627/oig_gf-oig-15-010_report_en.pdf.

[48] Shiffman, J. Donor funding priorities for communicable disease control in the developing world. Health Policy and Planning 21, 411–420 (2006).

[49] Carroll, M., Ruzgar, N., Fedatto, M., Schultz, K. & Cheung, M. ‘Show me the money’: An analysis of US global health funding from 1995 to 2019. Journal of Global Health 14, 04173 (2024).

[50] Shiffman, J. Has donor prioritization of HIV/AIDS displaced aid for other health issues? Health Policy and Planning 23, 95–100 (2008).

[51] Zakumumpa, H. et al. Understanding the persistence of vertical (stand-alone) HIV clinics in the health system in Uganda: a qualitative synthesis of patient and provider perspectives. BMC Health Services Research 18, 690 (2018).

[52] Palma, A. M. et al. Can the success of HIV scale-up advance the global chronic NCD agenda? Global Heart 11, 403 (2016).

[53] World Health Organization. Everybody’s business: strengthening health systems to improve health outcomes: WHO’s framework for action (2007). URL https://iris.who.int/handle/10665/43918. Place: Geneva Publisher: World Health Organization.

[54] Hafner, T. & Shiffman, J. The emergence of global attention to health systems strengthening. Health Policy and Planning 28, 41–50 (2013).

[55] Nimako, K. & Kruk, M. E. Seizing the moment to rethink health systems. The Lancet Global Health 9, e1758–e1762 (2021).

[56] Brink, D. T. et al. Impact of an international HIV funding crisis on HIV infections and mortality in low-income and middle-income countries: a modelling study. The Lancet HIV 12, e346–e354 (2025).

[57] Ballif, M. et al. The long-term impact of the COVID-19 pandemic on tuberculosis care and infection control measures in anti-retroviral therapy (ART) clinics in low-and middle-income countries: a multiregional site survey in Asia and Africa. BMJ Global Health 10 (2025).

[58] Arsenault, C. et al. COVID-19 and resilience of healthcare systems in ten countries. Nature Medicine 28, 1314–1324 (2022).

[59] Herrera, C. A. et al. COVID-19 disruption to routine health care services: How 8 Latin American and Caribbean countries responded. Health Affairs 42, 1667–1674 (2023).

[60] Marti, M. et al. Impact of the COVID-19 pandemic on TB services at ART programmes in low- and middle-income countries: a multi-cohort survey. Journal of the International AIDS Society 25, e26018 (2022).

[61] Brazier, E. et al. Service delivery challenges in HIV care during the first year of the COVID-19 pandemic: results from a site assessment survey across the global IeDEA consortium. Journal of the International AIDS Society 25, e26036 (2022).

[62] Dheda, K. et al. The intersecting pandemics of tuberculosis and COVID-19: population-level and patient-level impact, clinical presentation, and corrective interventions. The Lancet Respiratory Medicine 10, 603–622 (2022).

[63] WHO. More malaria cases and deaths in 2020 linked to COVID-19 disruptions. URL https://www.who.int/news/item/06-12-2021-more-malaria-cases-and-deaths-in-2020-linked-to-covid-19-disruptions.

[64] Nachega, J. B. et al. Minimizing the impact of the triple burden of COVID-19, tuberculosis and HIV on health services in sub-Saharan Africa. International Journal of Infectious Diseases 113, S16–S21 (2021).

[65] Global Tuberculosis Report 2024 (2024). URL https://www.who.int/teams/global-tuberculosis-programme/tb-reports/global-tuberculosis-report-2024.

[66] Reid, M., Yamey, G., Goosby, E., Jamison, D. & Schäferhoff, M. Seizing opportunities to end TB: a call for ambition and optimism on world TB day. The Lancet 401, 1153 (2023).

[67] World Health Organization. Invisible numbers: the true extent of noncommunicable diseases and what to do about them (2022).

[68] Agravat, P. et al. Research funding for newborn health and stillbirths, 2011–20: a systematic analysis of levels and trends. The Lancet Global Health 11, e1794–e1804 (2023).

[69] Toetzke, M., Banholzer, N. & Feuerriegel, S. Monitoring global development aid with machine learning. Nature Sustainability 5, 533–541 (2022).

[70] Kuzmanovic, M., Frauen, D., Hatt, T. & Feuerriegel, S. Causal machine learning for cost-effective allocation of development aid. Proceedings of the 28th ACM SIGKDD Conference on Knowledge Discovery and Data Mining 5283–5294 (2024).

[71] Bump, J. B. Global health aid allocation in the 21st century. Health Policy and Planning 33, i1–i3 (2018).

[72] Organisation for Economic Co-operation and Development (OECD). Comparative study of data reported to the OECD creditor reporting system (CRS) and to the aid management platform (AMP). Organisation for Economic Co-operation and Development (OECD) (2009).

[73] Lui, M. & Baldwin, T. langid.py: An off-the-shelf language identification tool. In Zhang, M. (ed.) Proceedings of the ACL 2012 System Demonstrations, 25–30 (Association for Computational Linguistics, Jeju Island, Korea, 2012).

[74] Danilak, M. langdetect: A Python port of Google’s language-detection library. https://pypi.org/project/langdetect/ (2014).

[75] Google. Google Cloud Translation API. https://cloud.google.com/translate (2025).

[76] World Bank. Population, total (2025). World Development Indicators. Available at https://data.worldbank.org/indicator/SP.POP.TOTL (Accessed June 1, 2025).

[77] Dubey, A., et al. The Llama 3 herd of models. arXiv:2407.21783 (2024).

[78] Together AI. URL https://www.together.ai/. Accessed June 1, 2025.

[79] Open LLM leaderboard. URL https://huggingface.co/spaces/open-llm-leaderboard/open_llm_leaderboard. Accessed November 7, 2024.

[80] Feuerriegel, S. et al. Using natural language processing to analyse text data in behavioural science. Nature Reviews Psychology 4, 96–111 (2025).

[81] Giray, L. Prompt engineering with ChatGPT: a guide for academic writers. Annals of Biomedical Engineering 51, 2629–2633 (2023).

[82] Lin, Z. How to write effective prompts for large language models. Nature Human Behaviour 8, 611–615 (2024).

[83] Murray, C. J. L. & Lopez, A. D. The Global Burden of Disease: A comprehensive assessment of mortality and disability from diseases, injuries, and risk factors in 1990 and projected to 2020 (Harvard School of Public Health, Cambridge, MA, 1996).

[84] Ward, Z. J. & Goldie, S. J. Global Burden of Disease Study 2021 estimates: implications for health policy and research. The Lancet 403, 1958–1959 (2024).

[85] Institute for Health Metrics and Evaluation. GBD results (2025). URL https://vizhub.healthdata.org/gbd-results/. Accessed June 1, 2025.

[86] Ferrari, A. J. et al. Global incidence, prevalence, years lived with disability (YLDs), disability-adjusted life-years (DALYs), and healthy life expectancy (HALE) for 371 diseases and injuries in 204 countries and territories and 811 subnational locations, 1990–2021: a systematic analysis for the Global Burden of Disease Study 2021. The Lancet 403, 2133–2161 (2024).

[87] Mukaka, M. M. Statistics corner: A guide to appropriate use of correlation coefficient in medical research. Malawi medical journal: the journal of Medical Association of Malawi 24, 69–71 (2012).

[88] Schober, P., Boer, C. & Schwarte, L. A. Correlation coefficients: Appropriate use and interpretation. Anesthesia and analgesia 126, 1763–1768 (2018).

[89] Dybul, M. Health financing seen from the global level: beyond the use of gross national income. *Health Economics*, Policy and Law 12, 121–137 (2017).

[90] The Global Fund. Description of the 2023–2025 allocation methodology (2023). URL https://www.theglobalfund.org/media/12675/fundingmodel_2023-2025-allocations_methodology_en.pdf.

[91] Gandhi, G. Charting the evolution of approaches employed by the Global Alliance for Vaccines and Immunizations (GAVI) to address inequities in access to immunization: a systematic qualitative review of GAVI policies, strategies and resource allocation mechanisms through an equity lens (1999–2014). BMC Public Health 15, 1198 (2015).

[92] Sterck, O., Roser, M., Ncube, M. & Thewissen, S. Allocation of development assistance for health: is the predominance of national income justified? Health Policy and Planning 33, i14–i23 (2018).

[93] Fan, V. Y., Glassman, A. & Silverman, R. L. How a new funding model will shift allocations from the Global Fund to fight AIDS, tuberculosis, and malaria. Health Affairs 33, 2238–2246 (2014).

[94] Norheim, O. F., Jha, P., Admasu, K. et al. Avoiding 40% of the premature deaths in each country, 2010–30: review of national mortality trends to help quantify the UN Sustainable Development Goal for health. Lancet 385, 239–252 (2015).

[95] Woods, B., Revill, P., Sculpher, M. & Claxton, K. Country-level cost-effectiveness thresholds: initial estimates and the need for further research. Value Health 19, 929–935 (2016).

[96] Ottersen, T. et al. New approaches to ranking countries for the allocation of development assistance for health: choices, indicators and implications. Health Policy and Planning 33, i31–i46 (2018).

[97] Jamison, D. T., Summers, L. H., Alleyne, G. et al. Global health 2035: a world converging within a generation. Lancet 382, 1898–1955 (2013).

[98] World Health Organization. Macroeconomics and Health: Investing in Health for Economic Development (2001). URL https://iris.who.int/bitstream/handle/10665/42435/924154550X.pdf.

[99] U.S. National Library of Medicine. Medical subject headings. URL https://www.nlm.nih.gov/mesh/meshhome.html. Accessed June 1, 2025.

